# Investigating causal relationships between loneliness, social isolation and health

**DOI:** 10.1101/2024.11.26.24317985

**Authors:** Darren D. Hilliard, Robyn E. Wootton, Hannah M. Sallis, Margot P. Van De Weijer, Jorien L. Treur, Pamela Qualter, Padraig Dixon, Eleanor C.M. Sanderson, David J. Carslake, Rebecca C. Richmond, Patricia Beloe, Lucy Turner-Harris, Lauren Bowes Byatt, Marcus R. Munafò, Zoe E. Reed

## Abstract

Loneliness and social isolation are important public health concerns due to their associations with a range of health outcomes. However, it is difficult to ascertain whether loneliness and social isolation cause those outcomes or whether the observed associations are biased by confounding and reverse causation. In this study we used a triangulation approach combining observational analysis, sibling control design, and Mendelian Randomisation (a genetically informed causal inference approach), to draw robust conclusions about these relationships. Using a combination of publicly available genome-wide association study (N= 17,526 to 2,083,151) and UK Biobank data (N= 8,075 to 414,432), we examined relationships between loneliness and social isolation and outcomes related to physical health, mental health and wellbeing and general health (reflecting both physical and mental health e.g., multimorbidity). Our results provide evidence for causal effects of loneliness and social isolation on poorer mental health and wellbeing and of loneliness on poorer general health. Evidence was generally stronger for loneliness compared to social isolation. We do not find evidence of effects on specific physical health outcomes; however, we cannot definitively rule out causal relationships. Interventions targeting loneliness and social isolation may be effective strategies for improving general health, mental health and wellbeing outcomes.

## Introduction

Loneliness and social isolation have become important public health issues in recent years, both being associated with poorer health outcomes, including specific physical and mental health outcomes (e.g., cardiovascular disease, depression, anxiety, and wellbeing), and general health outcomes (which capture both physical and mental health e.g., mortality) ^1–8^. Loneliness and social isolation are related concepts, but differ in how they are defined and measured. Loneliness is a subjective measure of how individuals feel about their relationships (i.e., perceived social connectedness) compared to what they may desire from relationships, whereas social isolation is an objective measure of the number and type of social connections an individual has ^9^.

There have been numerous studies examining the relationships between loneliness and/or social isolation, and health which have predominantly included observational analyses using cross-sectional data, demonstrating that these associations exist ^4,8,10–13^. There is also longitudinal evidence suggesting loneliness and social isolation may influence later health outcomes ^1,3,14^. Previous studies suggest that associations between loneliness and social isolation and health may be large, for example, loneliness was found to be associated with a 2.3 times higher likelihood of depression ^7^ and social isolation was found to be associated with a 1.5 times higher risk of coronary artery disease (CAD) ^15^. However, establishing whether these associations reflect causal relationships, and the direction of causation, can be difficult. Most previous studies have been unable to fully address this because they may be subject to residual confounding of the relationships between loneliness/social isolation and health outcomes, and reverse causation ^3^.

One approach that can overcome these issues of confounding and reverse causation, is Mendelian Randomisation (MR) ^16–19^, a genetically informed causal inference approach. MR uses genetic variants (single nucleotide polymorphisms; SNPs) as instrumental variables (IVs) for an exposure of interest (e.g., loneliness and social isolation) to estimate potentially causal relationships between these exposures and outcomes of interest (providing assumptions are satisfied ^19^). Whilst there have been many previous observational and longitudinal studies in this area of research, there have been few MR studies. Of those that have been conducted, some report evidence of causal effects of loneliness/social isolation on increased risk of various health outcomes, predominantly for mental health outcomes ^20–22^. Other MR studies have not found evidence for causal effects of loneliness/social isolation on both physical and mental health outcomes ^23–25^, reflecting mixed findings in this area. However, many of those studies have limitations in terms of varying definitions of the exposure ^20,22,25^, using less stringent p-value thresholds for instrument selection ^20,22^, and not including appropriate sensitivity analyses or not interpreting results appropriately in light of these ^20,22,25^. A more recent MR study, which also included observational analyses, found evidence of causal effects of loneliness on only 6 of the 26 physical and mental health outcomes examined (hypothyroidism, asthma, sleep apnoea, depression, psychoactive substance abuse and hearing loss) ^26^. In the current study we have built on previous MR analyses by using more specific measures of loneliness and social isolation (previous measures have captured multiple related concepts making it difficult to disentangle specific effects), including a wider number of relevant outcomes (such as rates of hospital admissions and quality-adjusted life years), including a greater range of sensitivity analyses, and conducting MR alongside other analyses to strengthen causal inference. These aspects are particularly important given that loneliness and social isolation are complex phenotypes and may violate key assumptions of MR.

We investigated specific health outcomes that are considered to have the greatest global ‘burden’ ^27,28^ and that have, in previous studies, been found to be associated with loneliness and/or social isolation. We categorised these into three broad categories of general health, physical health, and mental health and wellbeing. We employed a triangulation approach ^29,30^ that consisted of three parts: 1) observational analyses to establish whether associations exist between loneliness or social isolation and the health outcomes, adjusting for important potential confounders; 2) sibling control analyses to facilitate control for familial confounding (e.g., parental socioeconomic status) and shared genetic predispositions; and 3) MR analyses to estimate potential causal effects, reducing bias from confounding and reverse causation. These analyses, in combination, provide evidence as to whether the associations reflect causal effects. Consistent results across all three analyses, which each have different strengths and avoid different sources of bias, enables us to draw more robust conclusions about the relationships between loneliness/social isolation and health outcomes ^29^.

## Results

We combined observational analyses (cross-sectional and some with outcomes at a later timepoint, including time-to-event analyses), sibling control analyses, bidirectional two-sample MR (2SMR) ^16,31^ and one-sample MR (1SMR) ^32^ analyses in our triangulation approach. We used this to investigate the relationships between both loneliness and social isolation and a range of physical, general, and mental health outcomes, with the caveat that some outcomes were not available for either the 1SMR approach (as individual data was needed) or the 2SMR approach (as GWAS data was needed). The sibling control analyses included two estimates: between- and within-family effects. The within-family effect provided a less biased estimate as it accounts for confounding factors shared within families. We used a combination of publicly available genome-wide association study (GWAS) data for 2SMR and for the exposure in 1SMR, and data from UK Biobank for observational, sibling control and 1SMR analyses (see Methods for details). We also conducted multivariable MR (MVMR) to examine the independent effects of loneliness and social isolation on health.

The loneliness measure in UK Biobank captured whether the participant often felt lonely, and in the GWAS captured a combination of this same measure and other composite measures of loneliness (**Supplementary Table S2**). The social isolation measure in UK Biobank and the GWAS we conducted was a continuous variable combining frequency of friend/family visits and the number of people living in their household. The final measure had values between 0 and 2, where 2 indicated greater social isolation. We examined outcomes related to general health (hospital admissions ^33^, quality-adjusted life years [QALYs] ^34,35^, multimorbidity and mortality/death ^36,37^) ^12^, physical health (cardiovascular outcomes including coronary artery disease [CAD], heart failure ^38,39^, stroke, systolic blood pressure [SBP] ^40,41^ type 2 diabetes [T2D] ^42,43^) and mental health ^7^ (self-harm ^44^, suicidal attempts ^45^, depression diagnosis, a continuous depression trait ^46^, anxiety diagnosis and a continuous anxiety trait ^47^); wellbeing was captured by positive affect/happiness, meaning in life, a spectrum of wellbeing traits and life satisfaction ^1^) – see Methods for details.

### UK Biobank sample description

A description of the UK Biobank data used in our analyses is presented in **Table 1**.

**Table 1.** UK Biobank sample description.

| **Phenotype** | **N** | **Mean (SD) or percentage (%)** |
| --- | --- | --- |
| Age | 422,899 | 56.45 (8.10) years |
| Sex | 422,899 | Female: 54% |
| Townsend deprivation index | 422,356 | -1.28 (3.11) |
| Years of education | 401,953 | 14.99 (5.21) |
| Household income before tax | 357,567 | Less than £18,000: 22%<br>£18,000 to £30,999: 25%<br>£31,000 to £51,999: 26%<br>£52,000 to £100,000: 21%<br>Greater than £100,000: 6% |
| Ethnicity (self-reported responses) | 420,389 | White: 94%<br>Asian or Asian British (Indian, Pakistani, Bangladeshi): 2%<br>Black or Black British: 2%<br>Mixed or any other Asian background, or Chinese or other ethnic group: 2% |
| Disability | 411,595 | 33% |
| Number of ACEs | 140,757 | 0.76 (1.13) |
| Loneliness | 414,432 | 19% |
| Social isolation (on a scale of 0-2) | 412,650 | 1.01 (0.34) |
| Number of hospital admissions | 422,899 | 4.18 (13.66) |
| QALYs | 343,796 | 0.81 (0.20) |
| Multimorbidity greater than or equal to 2 | 422,790 | 55% |
| Death in follow-up | 422,898 | 7% |
| CAD in follow up | 400,050 | 7% |
| Heart failure in follow up | 420,640 | 3% |
| Stroke in follow up | 415,369 | 2% |
| Systolic blood pressure | 421,649 | 137.79 (18.68) mmHg |
| Type 2 diabetes | 422,899 | 5% |
| Self-harm | 133,146 | 4% |
| Suicide attempt | 132,956 | 2% |
| Depression | 422,899 | 14% |
| Depression trait (PHQ-9 score; on a scale of 0-27) | 131,040 | 2.75 (3.68) |
| Anxiety | 422,899 | 9% |
| Anxiety trait (GAD-7 score; on a scale of 0-21) | 131,638 | 2.15 (3.40) |
| Positive affect/happiness (on a scale of 1-6) | 229,531 | 2.52 (0.74) |
| Meaning in life (on a scale of 1-5) | 130,380 | 3.69 (0.83) |

*Note that the samples used in each analysis are a subset of this data depending on data availability.*
*SD=standard deviation, ACEs=adverse childhood experiences, QALYs= Quality-adjusted life years,*
*CAD=Coronary artery disease, PHQ-9=Patient Health Questionnaire 9-question version, GAD-7=Generalized Anxiety Disorder 7-question version.*

### Correlations

In UK Biobank, loneliness and social isolation had a phenotypic correlation of 0.13 (95% CI: 0.13 to 0.13). The genetic correlation (i.e., correlated genetic influences on both traits), based on GWAS data, was 0.31 (95% CI: 0.25 to 0.38). This demonstrates that the two phenotypes are capturing partially overlapping, but distinct traits.

### Triangulation approach

Results from our observational analyses, sibling control analyses, 1SMR and 2SMR analyses where loneliness and social isolation were the exposures are summarised in **Figures 1 to 6** and detailed in **Supplementary Materials Tables S6** (observational), **S9** (sibling control), **S10** (1SMR) and **S14** (2SMR). The effect sizes from these analyses are not always directly comparable because of the different approaches and measures used. We were mainly interested in how consistent the direction of effect was across the different analyses, with different assumptions, and how similar effect sizes were where approaches were similar (e.g., observational and sibling control). Details of each analysis can be found in the Methods. We have presented our main results for each of these outcome groups below, followed by sensitivity analyses.

**Figure 1.**
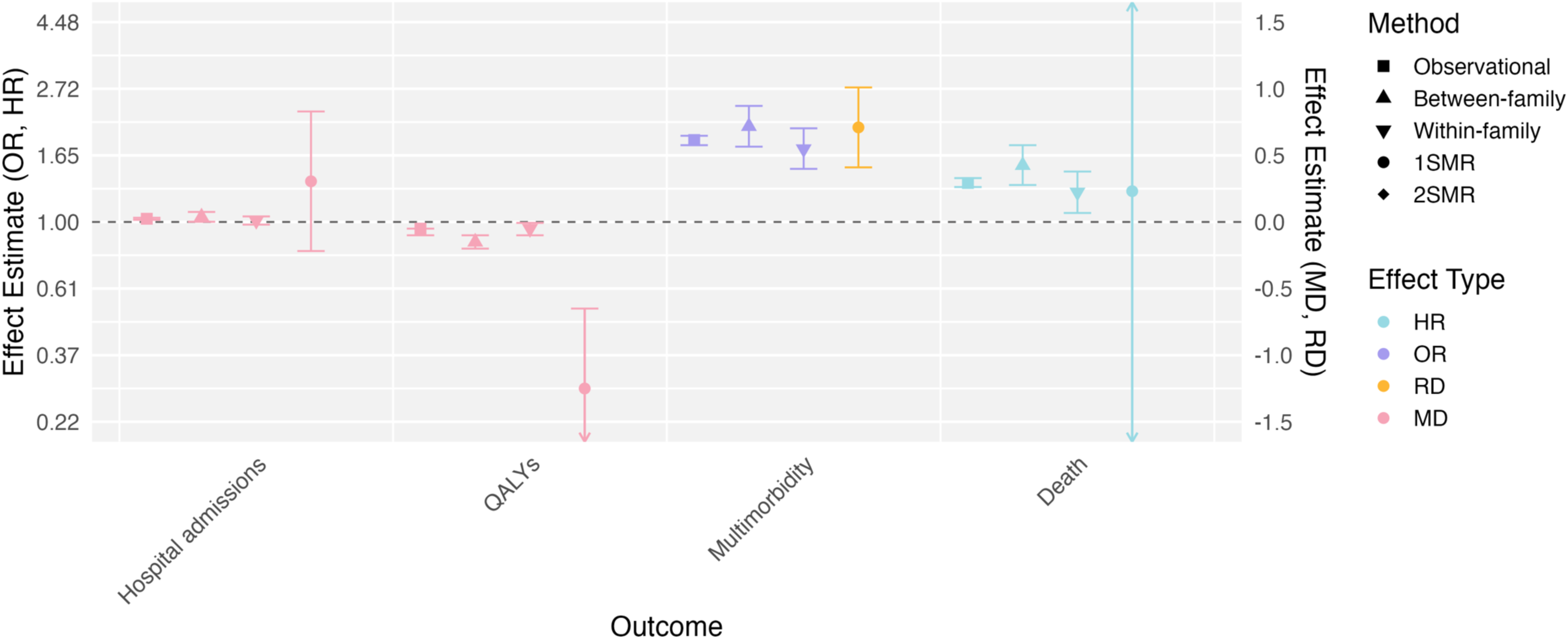
Main results from analyses examining relationships between loneliness and general health outcomes. 1SMR= One-sample Mendelian Randomisation, 2SMR= Two-sample Mendelian Randomisation, QALYs= Quality adjusted life years. There are no 2SMR results for these outcomes as these data were not available in genome-wide association studies. For continuous outcomes the effect estimate presented is the standardised mean difference (MD). For binary outcomes the effect estimate is either presented as the hazards ratio (HR) where Cox proportional hazards models were used, the odds ratio (OR, plotted as the log(OR)) where logistic regression models were used or for 2SMR with a binary outcome, or the risk difference (RD) for 1SMR analyses where logistic regression models were not appropriate to use. For one-sample MR binary outcomes this is the approximate RD of the outcome per unit increase in genetically predicted loneliness and for continuous outcomes this is the MD in the outcome per unit increase in genetically predicted loneliness. Effects for hospital admissions reflect the standardised count of times they have been admitted to hospital, for QALYs this is the standardised percentage change in QALYs per year of follow-up, for multimorbidity this is whether someone had two or more health conditions.

### Triangulation approach for general health outcomes

Overall, we found evidence across all approaches for causal effects of being lonely on decreased QALYs (*p*≤0.001 to 0.03) and increased risk of multimorbidity (*p*≤0.001) (**Figure 1**). For death and hospital admissions, there were associations between being lonely and increased hazards ratios for death (*p*≤0.001 to 0.83) as well as increased rates of hospital admissions (*p*≤0.001 to 0.50) across all analyses, other than MR analyses (**Figure 1**). We also found associations between increased social isolation and increased hazards ratios for death (*p*≤0.001 to 0.19) across all analyses, other than MR analyses (**Figure 2**). We did not find consistent evidence of causal effects where social isolation was the exposure for any of the other general health outcomes, as effects were only found in observational analyses (*p*≤0.001 to 0.02) (**Figure 2**). We note that the Cox proportional hazards models used for the 1SMR where death was the outcome were likely too imprecise to reliably assess evidence of a causal effect due to the complexity of the models. Additionally, where we do not see evidence of causal effects, confidence intervals were wide so should not be interpreted as evidence of absence of a causal effect.

**Figure 2.**
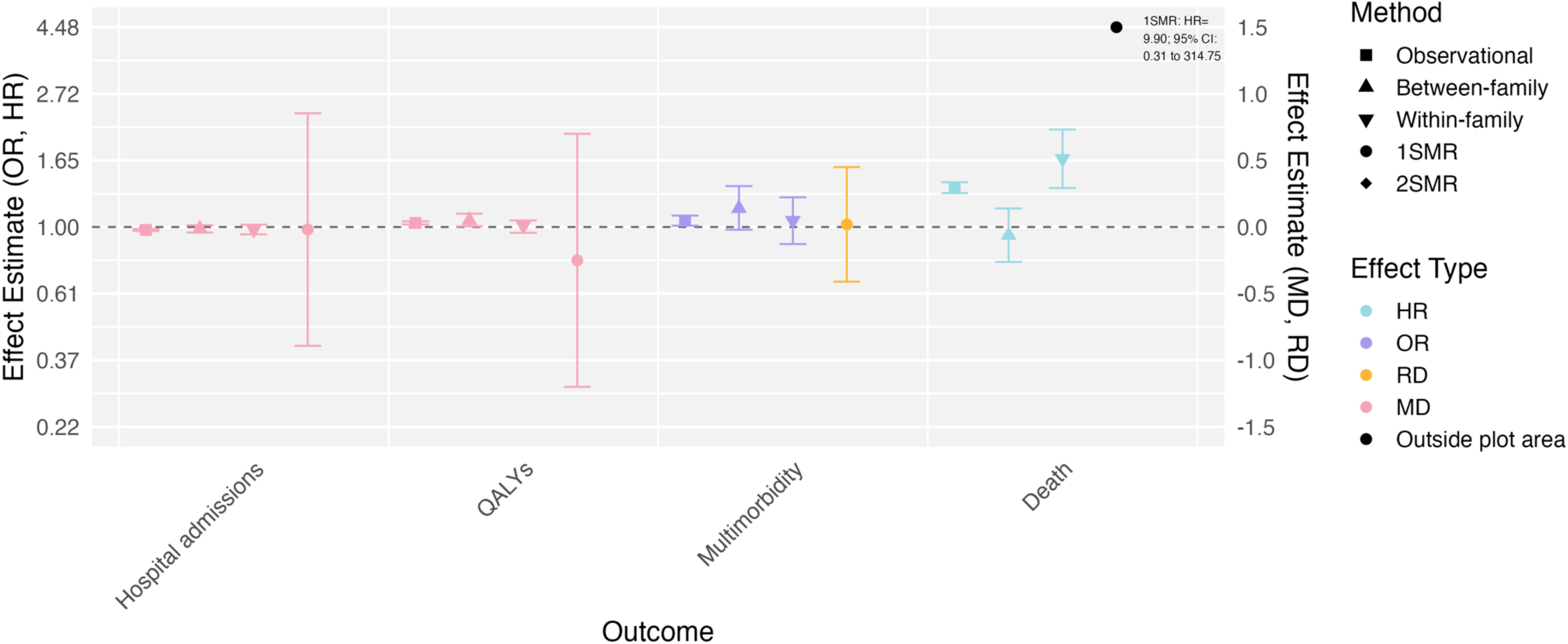
Main results from analyses examining relationships between social isolation and general health outcomes. 1SMR= One-sample Mendelian Randomisation, 2SMR= Two-sample Mendelian Randomisation, QALYs= Quality adjusted life years. There are no 2SMR results for these outcomes as these data were not available in genome-wide association studies. The point estimate for the 1SMR analysis for death was too large to plot alongside other points and therefore the results are shown in text on the plot. For continuous outcomes the effect estimate presented is the standardised mean difference (MD). For binary outcomes the effect estimate is either presented as the hazards ratio (HR) where Cox proportional hazards models were used, the odds ratio (OR, plotted as the log(OR)) where logistic regression models were used or for 2SMR with a binary outcome, or the risk difference (RD) for 1SMR analyses where logistic regression models were not appropriate to use. For one-sample MR binary outcomes this is the approximate RD of the outcome per unit increase in genetically predicted social isolation and for continuous outcomes this is the MD in the outcome per unit increase in genetically predicted social isolation. Effects for hospital admissions reflect the standardised count of times they have been admitted to hospital, for QALYs this is the standardised percentage change in QALYs per year of follow-up, for multimorbidity this is whether someone had two or more health conditions.

**Figure 3.**
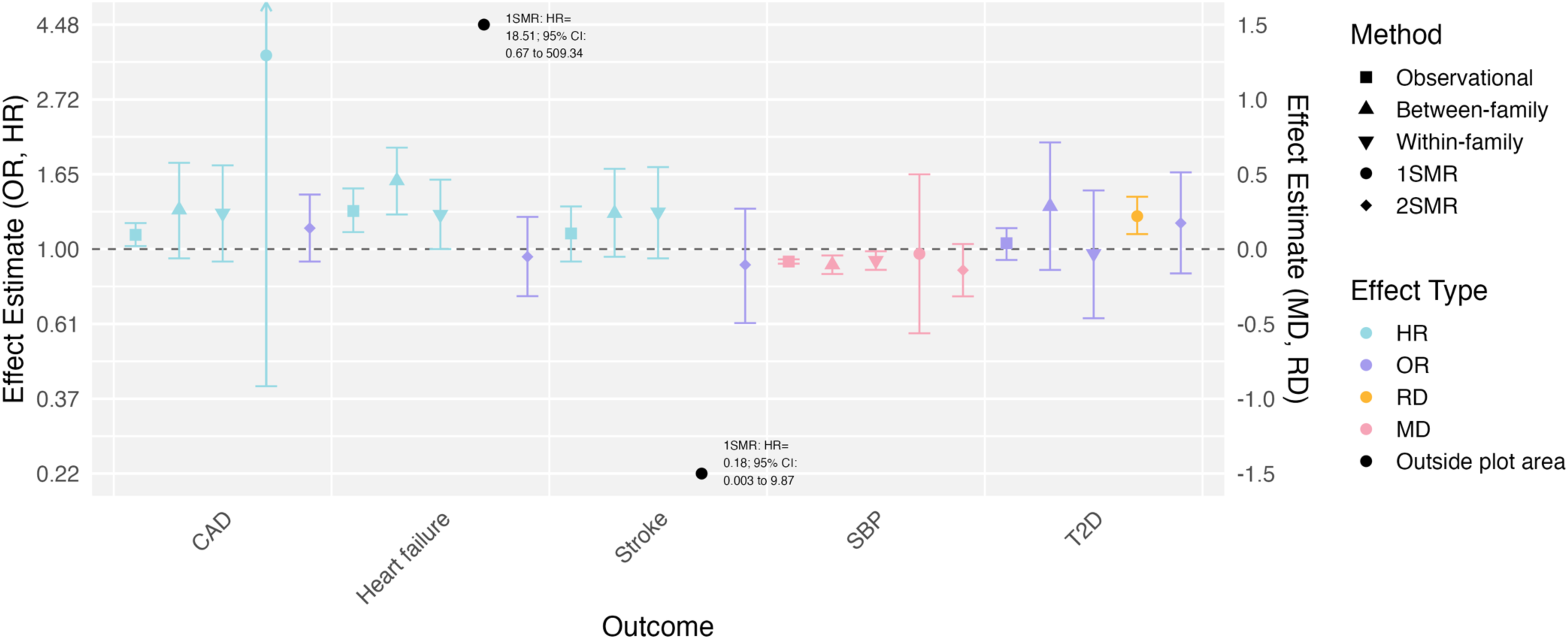
Main results from analyses examining relationships between loneliness and physical health outcomes. 1SMR= One-sample Mendelian Randomisation, 2SMR= Two-sample Mendelian Randomisation, CAD=Coronary artery disease, T2D=type 2 diabetes, SBP=systolic blood pressure. The point estimates for the 1SMR analysis for heart failure and stroke were too large to plot alongside other points and therefore the results are shown in text on the plot. For continuous outcomes the effect estimate presented is the standardised mean difference (MD). For binary outcomes the effect estimate is either presented as the hazards ratio (HR) where Cox proportional hazards models were used, the odds ratio (OR, plotted as the log(OR)) where logistic regression models were used or for 2SMR with a binary outcome, or the risk difference (RD) for 1SMR analyses where logistic regression models were not appropriate to use. For one-sample MR binary outcomes this is the approximate RD of the outcome per unit increase in genetically predicted loneliness and for continuous outcomes this is the MD in the outcome per unit increase in genetically predicted loneliness. Effects for systolic blood pressure reflect standardised millimetres of mercury (mmHg).

**Figure 4.**
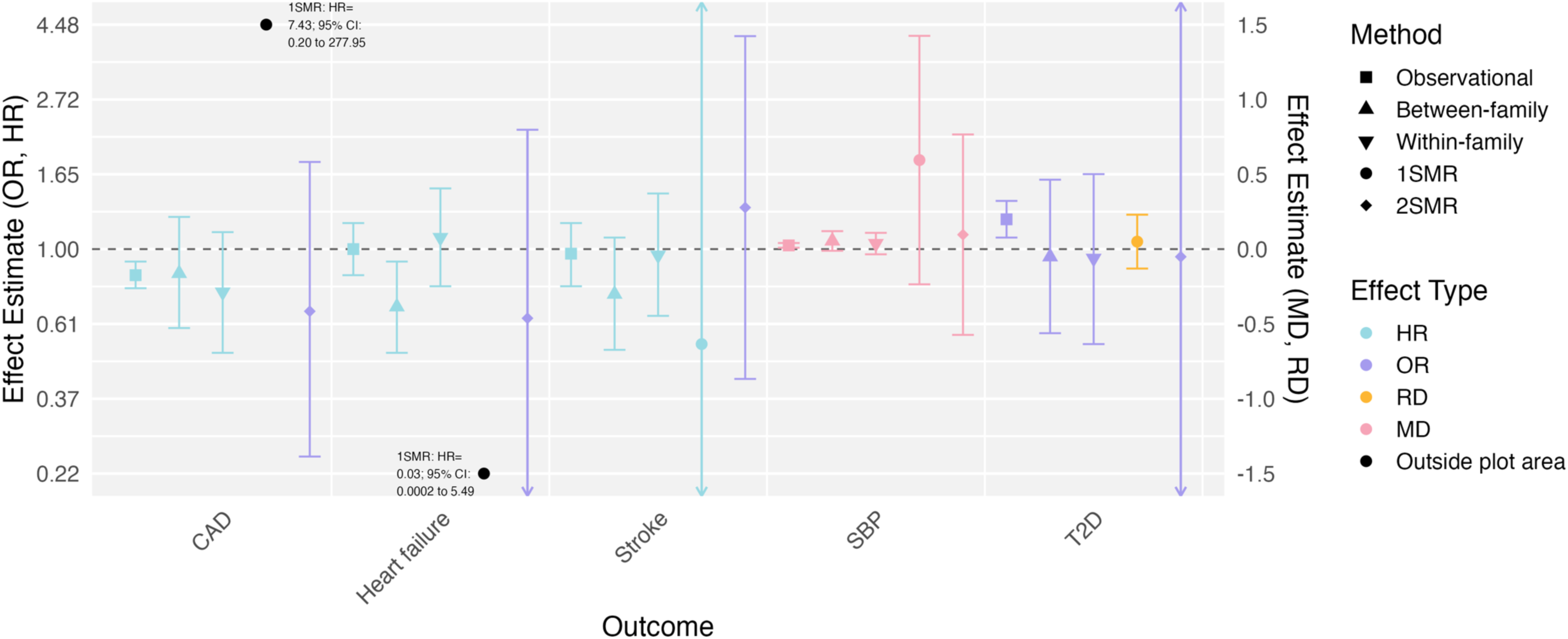
Main results from analyses examining relationships between social isolation and physical health outcomes. 1SMR= One-sample Mendelian Randomisation, 2SMR= Two-sample Mendelian Randomisation, CAD=Coronary artery disease, T2D=type 2 diabetes, SBP=systolic blood pressure. The point estimates for the 1SMR analysis for CAD and heart failure were too large to plot alongside other points and therefore the results are shown in text on the plot. For continuous outcomes the effect estimate presented is the standardised mean difference (MD). For binary outcomes the effect estimate is either presented as the hazards ratio (HR) where Cox proportional hazards models were used, the odds ratio (OR, plotted as the log(OR)) where logistic regression models were used or for 2SMR with a binary outcome, or the risk difference (RD) for 1SMR analyses where logistic regression models were not appropriate to use. For one-sample MR binary outcomes this is the approximate RD of the outcome per unit increase in genetically predicted social isolation and for continuous outcomes this is the MD in the outcome per unit increase in genetically predicted loneliness. Effects for systolic blood pressure reflect standardised millimetres of mercury (mmHg).

**Figure 5.**
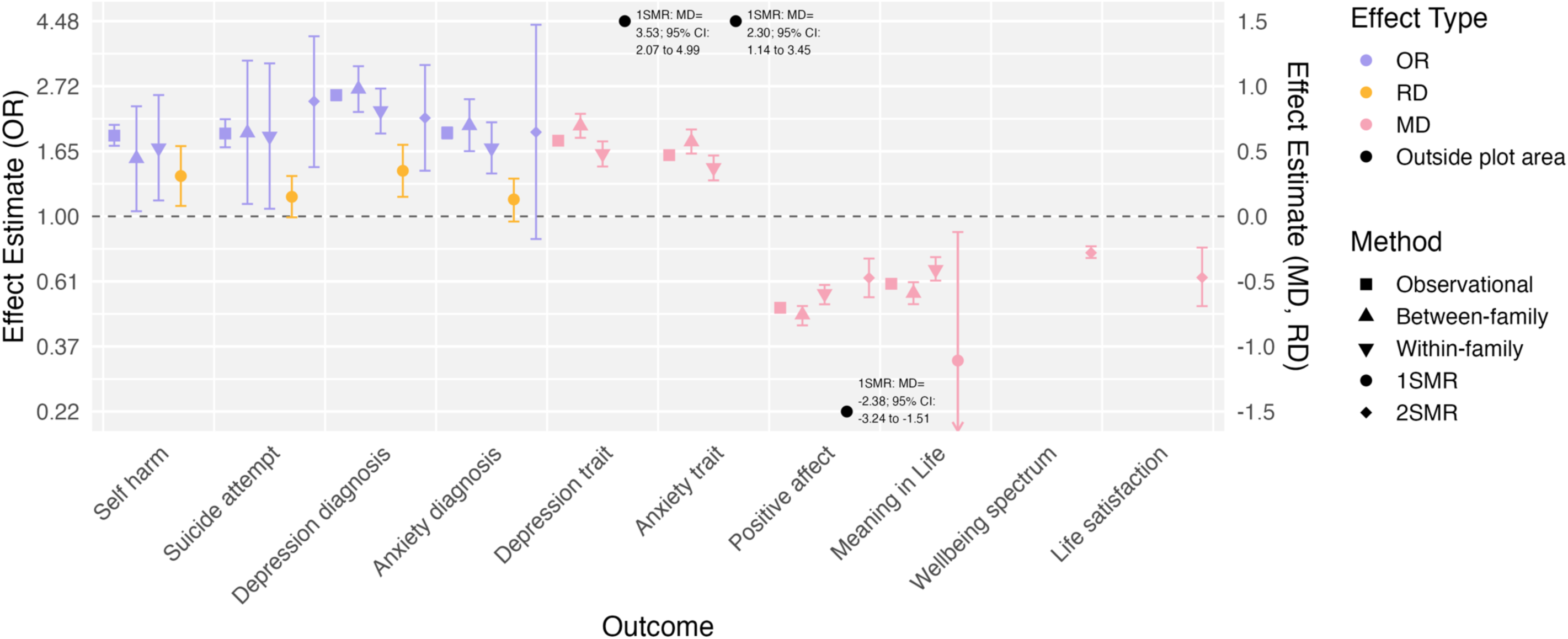
Main results from analyses examining relationships between loneliness and mental health and wellbeing outcomes. 1SMR= One-sample Mendelian Randomisation, 2SMR= Two-sample Mendelian Randomisation. There are no 2SMR results for self-harm, depression trait, anxiety trait or meaning in life as these data were not available in genome-wide association studies. There are only 2SMR results for wellbeing spectrum and life satisfaction because there were no equivalent measures in UK Biobank. The point estimates for the 1SMR analysis for depression trait, anxiety trait and positive affect were too large to plot alongside other points and therefore the results are shown in text on the plot. For continuous outcomes the effect estimate presented is the standardised mean difference (MD). For binary outcomes the effect estimate is either presented as the odds ratio (OR, plotted as the log(OR)) where logistic regression models were used or for 2SMR with a binary outcome, or the risk difference (RD) for 1SMR analyses where logistic regression models were not appropriate to use. For one-sample MR binary outcomes this is the approximate RD of the outcome per unit increase in genetically predicted loneliness and for continuous outcomes this is the MD in the outcome per unit increase in genetically predicted loneliness. Effects for the depression trait reflect the standardised score (ranging from 0 to 27), effects for the anxiety trait reflect the standardised score (ranging from 0 to 21), effects for happiness/positive affect reflect the standardised rating (ranging from 1 to 6), effects for meaning in life reflect the standardised rating (ranging from 1 to 5).

**Figure 6.**
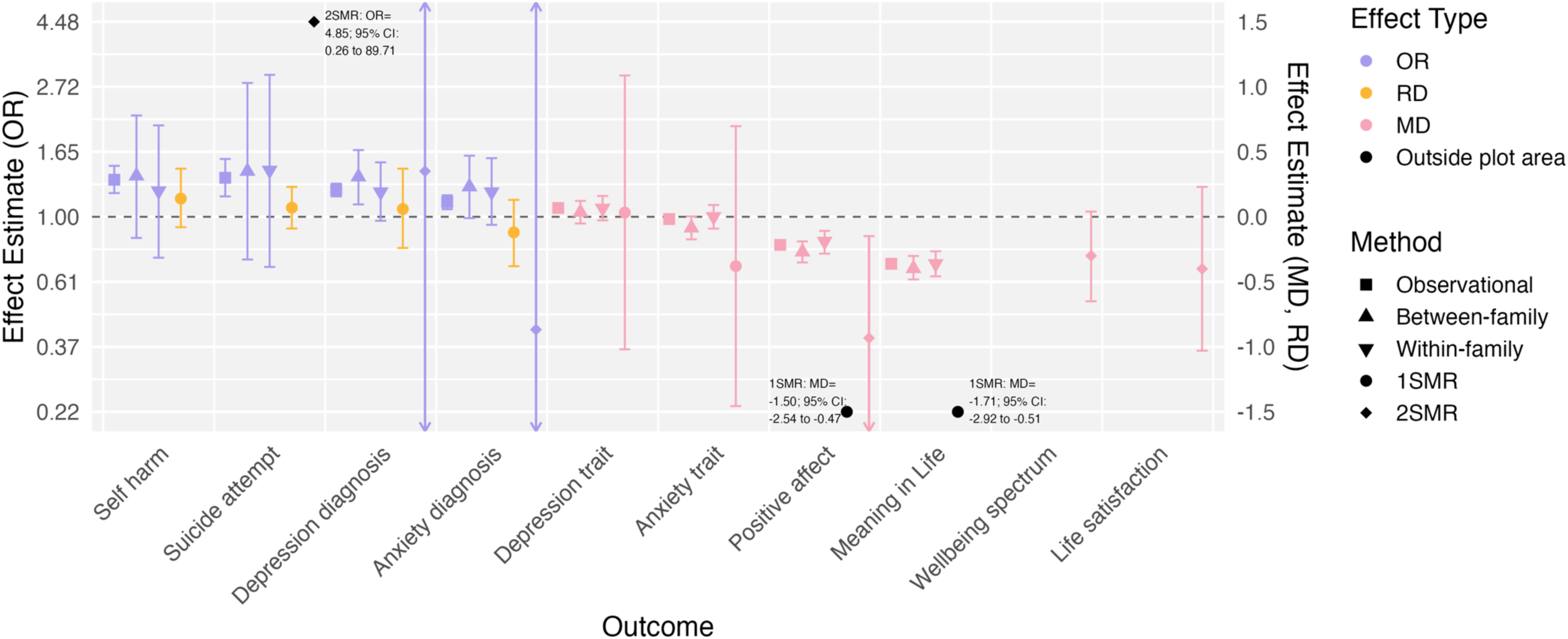
Main results from analyses examining relationships between social isolation and mental health and wellbeing outcomes. 1SMR= One-sample Mendelian Randomisation, 2SMR= Two-sample Mendelian Randomisation. There are no 2SMR results for self-harm, depression trait, anxiety trait or meaning in life as these data were not available in genome-wide association studies. There are only 2SMR results for wellbeing spectrum and life satisfaction because there were no equivalent measures in UK Biobank. The point estimates for the 2SMR analysis for suicide attempt and the 1SMR analysis for positive affect and meaning in life were too large to plot alongside other points and therefore the results are shown in text on the plot. For continuous outcomes the effect estimate presented is the standardised mean difference (MD). For binary outcomes the effect estimate is either presented as the odds ratio (OR, plotted as the log(OR)) where logistic regression models were used or for 2SMR with a binary outcome, or the risk difference (RD) for 1SMR analyses where logistic regression models were not appropriate to use. For one-sample MR binary outcomes this is the approximate RD of the outcome per unit increase in genetically predicted social isolation and for continuous outcomes this is the MD in the outcome per unit increase in genetically predicted social isolation. Effects for the depression trait reflect the standardised score (ranging from 0 to 27), effects for the anxiety trait reflect the standardised score (ranging from 0 to 21), effects for happiness/positive affect reflect the standardised rating (ranging from 1 to 6), effects for meaning in life reflect the standardised rating (ranging from 1 to 5).

### Triangulation approach for physical health outcomes

There was some evidence of effects of being lonely on decreased SBP (*p*≤0.001 to 0.91) and increased hazard ratios for heart failure (*p*≤0.001 to 0.72) observed across all analyses, other than the MR analyses. There was, however, a lack of evidence generally for being lonely on other physical health outcomes (CAD, stroke, T2D), other than observationally for increased hazards ratios of CAD (*p=*0.02) and in 1SMR analyses for increased T2D risk (*p*≤0.001). There was evidence for observational effects on increased hazards ratios of stroke and increased T2D risk, but after adjusting for ACEs and/or disability these effects attenuated **(Supplementary Table S6**). We did not find evidence of causal effects where social isolation was the exposure for any physical health outcomes, other than observationally for increased SBP (*p=*0.002), decreased hazards ratios of CAD (*p*≤0.001, although the effect in the unadjusted model was for increased hazards ratios of CAD) and increased T2D risk (*p=*0.002). With heart failure and stroke, effects were observed in unadjusted models but not in the adjusted models (**Supplementary Table S6**). We note that the Cox proportional hazards models used for the 1SMR where heart failure, CAD and stroke were outcomes, were also likely too imprecise to reliably assess evidence of a causal effect, due to the complexity of the models. Where we do not see evidence of causal effects, confidence intervals were wide so should not be interpreted as evidence of absence of a causal effect.

### Triangulation approach for mental health and wellbeing outcomes

Overall, we found evidence across all analyses for causal effects of being lonely on increased risk of self-harm (*p*≤0.001 to 0.01), suicide attempt (*p*≤0.001 to 0.06), depression diagnosis (*p*≤0.001), increased levels of the depression trait (*p*≤0.001) and anxiety trait (*p*≤0.001) and decreased levels of happiness/positive affect (*p*≤0.001), meaning in life (*p*≤0.001 to 0.03), wellbeing (*p*≤0.001) and life satisfaction (*p*≤0.001). There was some evidence of effects of being lonely on anxiety diagnosis (*p*≤0.001 to 0.13) in all analyses other than 1SMR and 2SMR, although the 2SMR analysis had lower power due to the smaller sample size compared to other outcome GWAS. We also found consistent evidence across analyses of potential causal effects of increased levels of social isolation on lower levels of happiness/positive affect (*p*≤0.001 to 0.02) and meaning in life (*p*≤0.001 to 0.008). However, this was not the case for the other mental health and life satisfaction outcomes, other than observationally for increased risk of self-harm (*p*≤0.001), suicide (*p*≤0.001), depression diagnosis (*p*≤0.001), increased depression trait (*p*≤0.001) and increased risk of anxiety diagnosis (*p*≤0.001). Where we do not see evidence of causal effects, confidence intervals were wide so should not be interpreted as evidence of absence of a causal effect.

### Sensitivity analyses for all outcomes

For the observational analyses using alternative models due to data being positively skewed (i.e., Gamma and negative binomial models) for hospital admissions, depression trait and anxiety trait, results were similar (**Supplementary Materials Table S7**). Similarly, when using a slightly different measure for social isolation (excluding children in household) results were similar (**Supplementary Materials Table S8**). We also conducted 1SMR analyses using unweighted genetic scores for loneliness from the published GWAS (we used the split sample approach in main analuses due to sample overlap and this additional analysis to compare results). Effects from this analysis were similar to or attenuated compared to the split sample main approach for loneliness (**Supplementary Materials Table S11**). We also used a 2SMR framework to conduct sensitivity analyses for the 1SMR analyses (using weighted median and weighted mode approaches). In these sensitivity analyses, effects were mostly attenuated and there was generally a lack of evidence of an effect with the weighted mode approach, but evidence of an effect (where effects were observed in the main analyses) with the weighted median approach (**Supplementary Materials Table S12**). This may suggest that some of the genetic variants used were weak, which could be attributed to horizontal pleiotropy (where the genetic variant influences the outcome through a pathway not via the exposure).

For our 2SMR analyses, effects tended to be consistent across 2SMR sensitivity analyses, with the exception of the MR-Egger ^48^ and simulation extrapolation (SIMEX)-adjusted MR-Egger. This may indicate the presence of horizontal pleiotropy, although given the wide confidence intervals and the low I-squared values (**Supplementary Materials Table S13**) it is possible that these tests were too imprecise to assess this reliably. Other tests **(Supplementary Materials Table S14)** did not show strong evidence for the presence of horizontal pleiotropy. Our MR-Causal Analysis Using Summary Effect estimates (CAUSE) ^49^ results indicated that there was evidence supporting a causal model over a sharing model for loneliness with depression, wellbeing spectrum and positive affect, and social isolation with wellbeing spectrum and positive affect. We also used Steiger filtering to estimate the percentage of genetic variants in the instrument that explained more variance in the exposure than the outcome, which can help to infer the most likely causal direction ^50^.

These results indicated that, for the most part, a high percentage (> 80%) of genetic variants explained more variance in the exposure than the outcomes, suggesting the exposures are more likely to cause the respective outcomes than vice versa. This was lower for loneliness and life satisfaction (58%), suggesting the causal direction is less clear in this case.

### Analyses with loneliness and social isolation as outcomes

We were interested in assessing whether relationships might be bidirectional using 2SMR with loneliness and social isolation as the outcomes, (see **Supplementary Materials Table S15**). Similar to analyses with loneliness and social isolation as exposures, we found evidence across the main 2SMR and sensitivity analyses for causal effects of depression diagnosis on increased loneliness and wellbeing spectrum, positive affect and life satisfaction on decreased loneliness.

For social isolation there was some suggestive evidence for depression diagnosis influencing increased social isolation, but this was inconsistent across sensitivity analyses. The most consistent results across sensitivity analyses were observed for decreased wellbeing spectrum, positive affect and life satisfaction on increased social isolation.

### Analyses examining independent effects of loneliness and social isolation

We conducted multivariable MR with both loneliness and social isolation as exposures to assess whether their influences on outcomes were independent (**Supplementary Materials Table S16**). Results supported 2SMR findings for increased loneliness on increased risk of suicide attempt, depression and decreased levels of wellbeing spectrum, positive affect and life satisfaction. These results were consistent across sensitivity analyses, with the exception of MR-Egger. This suggests that social isolation did not impact the influence of loneliness on these outcomes.

For social isolation the 2SMR effect observed on decreased positive affect seemed to attenuate with no evidence of a causal effect remaining when loneliness was also accounted for, suggesting loneliness may influence this effect (e.g., as a mediator).

## Discussion

Using a triangulation approach, our results provide consistent evidence across analyses that loneliness causally negatively impacts mental health and wellbeing outcomes, and that social isolation causally negatively impacts wellbeing outcomes. This is in line with previous studies demonstrating observational associations between loneliness, social isolation and poorer mental health ^1,7,8^ . Our findings suggest that effects on mental health and wellbeing may be stronger for loneliness and that loneliness could also be influencing the effect of social isolation on positive affect. We also found consistent evidence across analyses that being lonely causally negatively impacts some aspects of general health (i.e., increasing the risk of multimorbidity and decreasing QALYs), in line with previous studies ^1^. However, in contrast to previous observational studies ^2,14^ we did not find consistent evidence across analyses of causal effects of social isolation on general health, or of either loneliness or social isolation on specific physical health outcomes.

We found comparable effect sizes with previously published observational effects. Our results therefore suggest potentially large effects of loneliness on poorer mental health. We also found evidence of causal effects in the other direction (i.e., poorer mental health and wellbeing causing loneliness or greater social isolation). This suggests that the relationships between loneliness and social isolation and mental health and wellbeing may exacerbate each other.

The lack of evidence for causal effects of loneliness and social isolation on physical health outcomes and some general health outcomes does not mean that causal relationships do not exist. Our findings may, in fact, reflect an absence of evidence as opposed to evidence of absence. Our MR analyses, and some of our sibling control analyses, were not precise enough (i.e., confidence intervals were wide) to provide evidence for, or against, a causal interpretation in many cases. Often, we found evidence of effects observationally, where we adjusted for important potential confounders, but not in the sibling control or MR analyses. This may suggest that there are important familial confounders or shared genetic predispositions that influence these associations. Furthermore, the fact we find more consistent evidence of causal effects for loneliness on general health, but not specific physical health outcomes (which likely influence general health) is interesting and could be due to several reasons. First, it may suggest that the effects of individual physical health outcomes were too small to detect in our study, but the combined effects of these influence general health. Second, it may be that because the general health measures also capture mental health, that mental health outcomes are driving these effects. Third, we examined only a select number of physical health outcomes, and it may be that there are effects on other physical health outcomes not examined here (i.e., non-cardiovascular outcomes).

However, those we examined where chosen for being particularly burdensome health outcomes with previously observed associations with loneliness and/or social isolation.

Our results are in line with a recent study that included observational analyses in UK Biobank and MR analyses ^26^. That study differed from the current study in the specific exposures and GWAS used. They also included fewer analyses (i.e., no sibling control analyses or 1SMR analyses) and fewer sensitivity analyses, which are important given that loneliness and social isolation are complex phenotypes and MR assumptions e.g., around pleiotropy, are more likely to be violated. However, they found similar results to those in this study. Our findings draw on a robust pre-registered triangulation approach strengthening the conclusions drawn in both our study and this recent study.

A final aspect to discuss is that the social isolation measure we used is different to some previous observational studies in this area, which may partially explain the differential findings. Our measure of social isolation did not include group activities and was focused on household size and frequency of seeing friends/family. Unlike some previous studies ^51,52^, we opted not to include measures related to group activities or going to venues such as a gym or pub, because those may be capturing other behaviours that influence health outcomes through routes other than social interaction.

### Strengths and limitations

The main strength of our study is the use of a triangulation approach. The different methods we used have their own strengths and biases, but by interpreting the results together we have greater confidence in our findings. Another strength is the range of outcomes we explored which incorporate both self-reported and linked NHS electronic care record data, covering different aspects of general, physical and mental health and wellbeing in an in-depth manner. We also included an extensive number of sensitivity analyses across our approaches which address different biases and assumptions, resulting in more robust conclusions. Finally, we used data from large and recent GWAS and UK Biobank, a large cohort study with rich phenotypic data.

There are several limitations to the current study that should be considered when interpreting our findings. First, there were several analyses, particularly where social isolation was the exposure, where our MR estimates had wide confidence intervals meaning that power may have been lacking to detect effects. Second, our measure of loneliness in UK Biobank was a binary response to a single question asking whether the respondent was lonely, whereas other studies have used composite measures of loneliness. Similarly, our social isolation measure did not capture social interactions outside of the household, friends and family, potentially missing key sources of social interaction. Third, whilst we accounted for several important potential confounders in our observational and sibling control analyses, there may be factors that we could not account for. Fourth, results from our sibling analyses may not be generalisable because it required there to be at least two siblings from the same family participating in UK Biobank. In terms of representation, UK Biobank participants are generally healthier and less socioeconomically deprived than the general population ^53^ and our MR analyses were restricted to individuals of European ancestry. Therefore, our results may not be generalisable outside of those populations.

Fifth, loneliness and social isolation are unlikely to be directly caused by the genetic variants used as instruments in this study (i.e., they are biologically distal and the pathways from genetic variants to these exposures are likely to be complex). MR analyses are likely to be more reliable when exposures are more directly related to the genetic variants included as instruments ^16^. Additionally, this may increase the chances of there being pleiotropy (i.e., the instrument influencing the outcomes through pathways not via the exposure). Although we conducted a range of sensitivity analyses to address this, it may be that pleiotropic pathways do still exist.

### Future directions

Given the findings in this study there are some useful directions that future research could take. It is important to better understand the pathways from loneliness/social isolation to mental health and wellbeing outcomes to target effective interventions to improve these outcomes. Knowing specific timepoints and the impact of chronic loneliness (i.e., being lonely for a long period of time – we only assessed this at a single timepoint) and social isolation would also be beneficial, and this is something we were unable to examine in this study due to the data and approaches used. Finally, examining relationships with other physical outcomes would also be helpful.

### Conclusions

Overall, our triangulation study provided robust evidence suggesting potential causal effects of loneliness and social isolation on poorer mental health and wellbeing and of loneliness on poorer general health. Taken together, these findings suggest that loneliness, and potentially social isolation, are still important public health concerns, especially with respect to mental health and general health. Interventions targeting loneliness and social isolation may prove to be effective strategies for improving a range of mental health, wellbeing and general health outcomes.

## Methods

### Pre-registration

The analysis plan is pre-registered on the Open Science Framework (https://doi.org/10.17605/OSF.IO/GWPNY). Deviations from this analysis plan are outlined in the **Supplementary Materials (Section 1)**.

### Data sources and measures

*UK Biobank:* For conventional multivariable observational, sibling control, and 1SMR analyses we used phenotypic and genetic data from the UK Biobank, a large population-based prospective cohort of around 500,000 participants aged between 38 and 73 years and living in the UK, recruited between 2006 and 2010 ^54^. UK Biobank received ethics approval from the UK National Health Service Research Ethics Committee (REC reference for UK Biobank is 11/NW/0382) and all participants gave informed consent to the use of their anonymised data and samples for any health-related research and for UK Biobank to access their health-related records. We excluded participants who withdrew their consent using the latest withdrawal lists for this project (project number: 81499).

The phenotypic data we used are described in detail in **Supplementary Materials Table S1**. To summarise, the UK Biobank loneliness measure was a binary variable, where participants were asked whether they often felt lonely. The social isolation measure was a continuous variable we created by combining measures of frequency of friend/family visits and the number of people living in their household. We assigned values between 0 and 1 for each measure and then summed these to create a final measure with values between 0 and 2 where 2 indicated greater social isolation. We included the following general health outcomes; hospital admissions, quality-adjusted life years (QALYs), multimorbidity and mortality, the following physical health outcomes; coronary artery disease (CAD), heart failure, stroke, systolic blood pressure, and type 2 diabetes (T2D) and the following mental health/wellbeing outcomes; self-harm, suicide attempts, depression (binary/diagnosis and a continuous trait from the Patient Health Questionnaire 9-question version; PHQ-9, scale from 0-27), anxiety (binary/diagnosis and a continuous trait from the Generalized Anxiety Disorder 7-question version; GAD-7, scale from 0-21), positive affect (scale from 1-6) and meaning in life (scale from 1-5).

In UK Biobank there were 488,377 participants with genetic data available. Details on the quality control and other processing steps prior to using these data are described in **Supplementary Materials Section 2**. The number of UK Biobank participants for our main analyses ranged from 77,577 to 414,432 for observational analyses, 8,075 to 40,440 for sibling control analyses and 106,828 to 333,358 for 1SMR. The difference in sample sizes was due to the data available for each measure.

### Genome-wide association studies (GWAS)

We used publicly available GWAS summary statistics (N= 17,526 to 2,083,151) for our 2SMR analyses and for our exposures of loneliness and social isolation in 1SMR analyses (see **Supplementary Materials Table S2**). However, we conducted our own GWAS of social isolation in UK Biobank (see **Supplementary Materials Section 3, Supplementary Materials Figure S1 and Supplementary Materials Table S3**).

### Statistical analyses

In our triangulation approach we first conducted observational analyses to establish whether there was an association between loneliness or social isolation and a given health outcome. We then used sibling control and Mendelian Randomisation (MR) analyses to provide evidence of whether an association may reflect a causal relationship. Triangulation is particularly useful when different methods have different biases and assumptions. If results are consistent across the different methods, there is greater confidence in the results ^55^. Our triangulation approach was qualitative i.e., assessing whether there was consistent evidence of potential causal effect.

In the pre-registration we included power calculations for our MR and sibling control analyses. These did not guide how we conducted our analyses (i.e., we conducted them all regardless of power), but they did guide interpretation of our findings. The power calculations indicated that some analyses would likely be underpowered. For example, where anxiety and suicide were the outcomes for loneliness and social isolation exposures, and where social isolation was the exposure, many of these analyses may only be powered to detect larger effect sizes, and therefore results should be interpreted taking this into consideration.

All analyses were conducted in R version 4.1.0 ^56^.

### Correlation between loneliness and social isolation

We calculated the phenotypic Pearson correlations between the loneliness and social isolation measures in UK Biobank. We also calculated the genetic correlation between these using GWAS summary statistics for loneliness and social isolation using LD score regression^57^.

### Observational and sibling control analyses in UK Biobank

We conducted linear regressions (for continuous outcomes), logistic regressions (for binary outcomes) or Cox proportional hazard models (time-to-event outcomes) (i.e., all-cause mortality and incident CAD, heart failure and stroke). **Supplementary Materials Section 4** describes how we applied the Cox proportional hazards models in this study.

In all observational and sibling control analyses we adjusted for potential confounders with two levels of adjustment. In our first adjusted models we included potential demographic confounders (age when attended assessment centre, sex, assessment centre, socioeconomic position as measured by the Townsend deprivation index, education years based on qualifications ^58^, household income before tax and ethnicity). In the second adjusted model we included all previous potential confounders as well as long-standing illness/disability/infirmity and a score for adverse childhood experiences (ACEs) ^59^. We additionally conducted analyses using our first level of adjustment as well as 1) long-standing illness/disability/infirmity or 2) ACEs to examine the impact of these factors further. We note that disability, in particular, could be considered an outcome as well as a factor influencing loneliness, which could introduce potential collider bias and therefore results should be considered across all levels of adjustment. Details of potential confounders are presented in **Supplementary Materials Table S4**. We also conducted a number of additional sensitivity analyses to assess whether findings were robust under different models or with a different social isolation exposure accounting for number of children in the household (see **Supplementary Materials Section 5**).

In the sibling control analyses we only included (biological) siblings thus controlling for shared environmental confounding (e.g., childhood household SES) and partly shared genetic predispositions ^60^. The approach we used to identify siblings in UK Biobank to include in analyses is described in **Supplementary Materials Section 6**. In the sibling-control analyses, we calculated the sibship loneliness/social isolation average (the between-family effect) and each sibling’s deviation from the sibship average (the within-family effect, unbiased by shared familial confounders). Each of the outcomes was assessed using a model that included the between- and within-family estimates, and adjusting for the same factors as included in the observational models with the full sample.

### Mendelian Randomisation (MR) analyses

MR is a causal inference method that uses genetic variants as instrumental variables for an exposure of interest. MR is less prone to confounding by socioeconomic, environmental and behavioural characteristics, and by reverse causation, than conventional observational studies ^17,18^.

There are three core assumptions underlying MR:

1. The genetic instruments are robustly associated with the exposure of interest;
2. There is no confounding of the genetic instrument-outcome relationship;
3. The genetic instruments are only associated with the outcome via the exposure, and there are no direct effects between the genetic IV and the outcome

2SMR analyses used data obtained from GWAS of both the exposure and outcome of interest, whereas 1SMR analyses used GWAS data for the exposure of interest but individual level outcome data. Where data were available, we conducted both 2SMR analyses and 1SMR analyses. However, for some outcomes (hospital admissions, QALYs, multimorbidity, mortality and meaning in life) there were no GWAS available, or the data were only available in UK Biobank (resulting in full sample overlap with the exposure) and in this case, we only conducted 1SMR.

### One-sample MR

For 1SMR analyses we used the Applied Econometrics with R (AER) package in R ^61^ to conduct two-stage least squares regressions, except for where we used Cox proportional hazards models. For our continuous and binary outcomes we created genetic scores for our exposures of interest in UK Biobank and then used these in the two-stage least squares regression approach to estimate our causal effects. For the Cox proportional hazards models, we manually conducted the 1SMR using the ratio method ^62^. 1SMR analyses were not bidirectional.

In our 1SMR analyses we adjusted for age, sex, the first 20 principal components (PCs) from PC analysis of genotype data, assessment centre and genotyping chip (**Supplementary Materials Table S4**). Due to the exposure GWAS completely (social isolation) or partially (loneliness) being conducted in UK Biobank, and to thus avoid sample overlap which can bias estimates due to overfitting ^63^, we used a split sample approach ^64^. For this approach we split the sample into two random halves and in each half, we conducted a GWAS on social isolation and loneliness in UK Biobank (see **Supplementary Materials Section 3**). We used this GWAS data to create genetic scores in the other half of the sample, by using genetic variants and their weights with a p-value threshold of 1×10^−05^. We then used these to conduct our 1SMR analyses in each half of the sample. The results from the analyses in each half were then combined in a meta-analysis. Further details of this approach and sensitivity analyses conducted can be found in **Supplementary Materials Section 7**.

### Two-sample MR

We conducted bidirectional 2SMR analyses (i.e., with loneliness/social isolation as exposures and outcomes) using the ‘TwoSampleMR’ R package ^31^. We used summary data (including weights) of genetic variant-exposure genome-wide associations from published GWAS as described in **Supplementary Table S2** as genetic instruments for the exposures in our analyses. These were selected based on a p-value threshold of 5×10^−08^ for all analyses, except where suicide attempt (1×10^−06^) and anxiety (1×10^−05^) were the exposures. The steps taken to prepare these data for analyses are described in **Supplementary Materials Section 8**. For the main analyses we used the inverse-variance weighted (IVW) method ^65^. The sensitivity analyses assess various assumptions and potential biases in the MR approach. Inconsistent results may suggest bias from MR assumptions being violated. The sensitivity analyses included were MR-Egger ^48^, weighted median ^66^, MR using robust adjusted profile score (MR-RAPS) ^67^, MR Pleiotropy RESidual Sum and Outlier (MR-PRESSO) ^68^, MR lap ^69^ and MR-Causal Analysis Using Summary Effect estimates (CAUSE) ^49^. We also conducted Steiger filtering ^50^ to further infer the most likely causal direction. **Supplementary Materials Section 8** further describes these sensitivity analyses.

### Multivariable MR analyses

We additionally conducted multivariable MR (MVMR) ^70^ analyses to estimate independent effects of loneliness and social isolation on health outcomes (see **Supplementary Materials Section 9**).

## Data availability

Access details for the GWAS data used in this study are outlined in **Supplementary Table S5**. UK Biobank data are available through a procedure described at http://www.ukbiobank.ac.uk/using-the-resource/.

## Code availability

Analysis code is available from the University of Bristol’s Research Data Repository (http://data.bris.ac.uk/data/), at: To be added on acceptance.

## Acknowledgements

This research has been conducted using data from UK Biobank (project ID: 81499), a major biomedical database (www.ukbiobank.ac.uk). Part of the UK Biobank analyses used data provided by patients and collected by the NHS as part of their care and support. We would like to thank the research participants and employees of 23andMe, inc. for making this work possible, where GWAS data included 23andMe data. The MEGASTROKE project used for the stroke GWAS received funding from sources specified at www.megastroke.org/acknowledgments.html. A list of MEGASTROKE authors is provided here: www.megastroke.org/authors.html. The type 2 diabetes GWAS included data from the Million Veteran Program (MVP), Office of Research and Development, Veterans Health Administration, and was supported by the Veterans Administration (VA). The authors thank MVP staff, researchers, and volunteers, who have contributed to MVP, and especially participants who previously served their country in the military and now generously agreed to enrol in the study. (See www.research.va.gov/mvp/ for more details). The MVP GWAS data were provided through dbGaP under accession number phs001672. For the purpose of open access, the author(s) has applied a Creative Commons Attribution (CC BY) licence to any Author Accepted Manuscript version arising from this submission. Finally, we want to thank the following at the University of Bristol for their work on facilitating this project: Julie Lane (Pre-Award Finance), Darren Down (Post-Award Finance), Kim Cox (Post-Award Finance), Rajive Sharma (Contracts), John Birney (Contracts), Weili Qiu (MRC IEU Data Manager), Caroline Clancy-Cottle (Press Office) and Joanne Fryer (Press Office).

## Author contributions

Conceptualisation: DDH, ZER, REW, HMS, MPVDW, JLT, PQ, MRM. Methodology: ZER, REW, HMS, MPVDW, JLT, PD, ECMS, DJC. Formal analysis: ZER. Data Curation: ZER, RCR Writing - Original draft: DDH, ZER. Writing – Review and Editing: DDH, REW, HMS, MPVDW, JLT, PQ, PD, ECMS, DJC, RCR, PB, LTH, LBB, MRM, ZER. Supervision: DDH, ZER. Project administration: DDH, ZER. Funding acquisition: DDH, ZER.

