## Supplementary Materials for "Investigating causal relationships between loneliness, social isolation and health"

### 1. Deviations from pre-registered analysis plan

The analyses conducted deviated slightly from our pre-registered analysis plan (<https://doi.org/10.17605/OSF.IO/GWPNY>). These are listed below with the reason for the deviation.

- We did not conduct the Cox proportional hazards models using a piecewise regression or similar to examine the relationships between loneliness/social isolation and outcomes of coronary artery disease (CAD), heart failure, stroke and death over time. The main Cox proportional hazards models we conducted did not provide evidence of effects on these outcomes and therefore there was no clear value in conducting further analyses.
- We did not conduct multivariable Mendelian Randomisation (MVMR) within a one-sample Mendelian Randomisation (1SMR) context. This was due to a lack of strong evidence for effects with social isolation and the two-sample Mendelian Randomisation (2SMR) MVMR not revealing any strong results for social isolation.
- We did not conduct mediation MR. In our pre-registration we stated that we would prioritise physical health outcomes with mediation analyses, but given our lack of findings for physical health outcomes we did not conduct the mediation MR analyses.

### 2. Use of genetic data in UK Biobank

In UK Biobank there are 488,377 participants genotyped samples (49,979 genotyped using the UK BiLEVE array and 438,398 using the UK Biobank Axiom® Array). Pre-imputation quality control, phasing and imputation have been described elsewhere ^1^ and were already conducted on the data. To summarise multiallelic genetic variants and those with a minor allele frequency (MAF) ≤1% were removed. Phasing of genotype data was performed using a modified version of the SHAPEIT2 algorithm. Genetic variants were imputed to the Haplotype Reference Consortium (HRC) reference panel, using IMPUTE2 algorithms. A graded filtering with different imputation qualities for different MAF ranges was used (Info>0.3 for MAF>3%, Info>0.6 for MAF 1-3%, Info>0.8 for MAF 0.5-1% and Info>0.9 for MAF 0.1-0.5%), where MAF and info scores were recalculated on an in-house derived ‘European’ subset. Individuals with sex-mismatch or sex-chromosome aneuploidy were excluded. In-house quality control filtering of the UK Biobank data has been described in a published protocol ^2^. In addition, we restricted our sample for 1SMR analyses to include only individuals who self-reported as ‘White’ and ‘British’ and who had very similar genetic ancestry based on a principal components analysis of genotypes. Self-reported responses were from touchscreen questionnaire questions asking, ‘What is your ethnic group?’ (response options: White; Mixed; Asian or Asian British; Black or Black British, Chinese, Other ethnic group, Do not know; Prefer not to answer). If they selected ‘White’ then they were asked ‘What is your ethnic background?’ (response options: British; Irish; Any other white background; Prefer not to answer). We acknowledge that ethnicity is a complex social construct with different meanings across different contexts, and is distinct to, although often overlapping with, genetic ancestry ^3^. This was done to ensure that the UK Biobank sample was similar in ancestry to the GWAS samples to avoid any bias in the results due to potential differences in associations by ancestry. Previously estimated kinship coefficients using the KING toolset ^4^ identified 107,162 pairs of related individuals (third degree or closer). We used this as a basis to remove related individuals in all UK Biobank observational, sibling control and 1SMR analyses as described in the published protocol ^2^.

### 3. Genome wide association study (GWAS) of social isolation

We conducted our social isolation GWAS using the [MRC-IEU UK Biobank GWAS pipeline](https://github.com/MRCIEU/UKBiobankGWAS/tree/main) for 457,086 participants. We used BOLT-LMM software within the pipeline to conduct our GWAS, which used a linear mixed model accounting for relatedness and population stratification and restricted to individuals of ‘European’ ancestry. We adjusted for sex, genotyping chip, age and assessment centre. Filtering and quality control steps applied to the genetic data are described in **Supplementary Materials Section 2** i.e., multiallelic genetic variants or those with MAF ≤1% were removed and rarer genetic variants were required to have a higher imputation INFO score (Info>0.3 for MAF >3%; Info>0.6 for MAF 1-3%; Info>0.8 for MAF 0.5-1%; Info>0.9 for MAF 0.1-0.5%). Therefore, no additional filtering was applied after the GWAS was conducted. To obtain a set of genome-wide significant genetic variants to use in our analyses we restricted the data to those with a p-value <5x10^-08^, resulting in 1,534 genetic variants. We then clumped the data using the function in the ‘TwoSampleMR’ R package using an r2 of 0.001, window of 10,000 kb and the European 1000 genomes reference panel, resulting in 15 genetic variants to be used in subsequent analyses. A Manhattan plot and a Quantile-Quantile (QQ) plot of our GWAS is shown in **Supplementary Materials Figure S1**. The QQ plot and lambda value of 1.2 suggests there may be some population structure. However, we examined the linkage disequilibrium score intercept and ratio, where a value close to 1 for the intercept and a value close to 0 for the ratio indicates that any inflation is unlikely to be due to causes other than polygenic heritability (e.g., population structure. Our intercept value was 1.04 and our ratio value was 0.11, both reasonably close to 1 and 0, respectively. Details of the 15 genetic variants used in analyses are provided in **Supplementary Materials Table S3.**

### 4. Cox proportional hazards models

Cox proportional hazards models were used with the event defined as all-cause mortality or as incident CAD, heart failure or stroke. Follow-up began at the date of attending the assessment centre (between 13^th^ March 2006 and 1^st^ October 2010) and individuals who had the event in question before this date were excluded. Follow-up ended when a person had the event, when they died (if the event was not death) or on 30/09/2021; whichever came first. The latter date is the end of availability of hospital episode statistics data in our data download from UK Biobank. Time since joining the study was used as the time axis. We tested the proportional hazards assumption for each model using the cox.zph() function in the Survival R package.

### 5. Additional sensitivity analyses

Results for continuous outcomes were reported using robust standard errors, calculated using the ‘coeftest’ function from the ‘lmtest’ package in R. We also conducted sensitivity analyses for the outcome hospital admissions, depression trait and anxiety trait, where data were positively skewed, to assess whether alternative modelling approaches impacted the results. To assess this, we conducted sensitivity analyses using both Gamma and negative binomial models for the fully adjusted models. Finally, we also conducted additional analyses using a slightly different measure for social isolation. In this measure we attempted to account for number of children living in the household in case having children living in the home impacted the relationship between social isolation and health outcomes. To do this we used data on the number of children the person had given birth to (UK Biobank field ID: 2734) or fathered (UK Biobank field ID: 2405) and whether that person indicated that a child lived in their household (UK Biobank field ID: 6141). We subtracted the number of children the person had given birth to or fathered from the total household count if they indicated that a child lived at home. However, we ensured that this did not reduce the number of household members below the count of other people living in the household as reported (UK Biobank field ID: 6141).

We also report robust standard errors, calculated using the ‘coeftest’ function from the ‘lmtest’ package in R.

### 6. Details for identifying UK Biobank siblings

Siblings were defined as participants with a kinship coefficient (the probability that two alleles sampled at random from two individuals are identical by descent) between the probability of zero identical-by-state multiplied by –21 + 0.5 and 0.7, and a probability of zero identical-by-state (IBS0) (meaning there is a probability that the siblings have zero alleles in common at a particular locus) between 0.001 and 0.008. These parameters were previously determined from visual clustering ^5^ and are similar to those obtained elsewhere ^1,4^. We also removed adoptees by excluding individuals who responded yes to the question “Were you adopted as a child?”.

### 7. One sample Mendelian Randomisation details and sensitivity analyses

For the Cox Proportional hazards models in 1SMR we used the ratio method. In this approach we first used linear regression to estimate a mean difference in the exposure per unit of the genetic instrument, adjusted for covariates, providing the denominator for the ratio method. Then we used a Cox proportional hazards model to estimate the log(HR) of the outcome per unit of the genetic instrument, adjusted for covariates, providing the numerator for the ratio method. To get the MR estimate of the log(HR) we divided the numerator by the denominator. To obtain 95% confidence intervals we used the Taylor series formula ^6^.

When creating the genetic scores in the split sample approach, due to the low number of genome-wide significant genetic variants in each half sample for both loneliness and social isolation we used genetic variants and weights with a p-value threshold of 1x10^-05^ to create genetic scores in the other half of the sample. Using these genetic scores, we conducted our 1SMR analyses in each half and meta-analysed the results (the estimates from the 1SMR in each half of the data) using the ’meta’ package in R (with the metagen() function for a common/fixed effects model) ^7^. We also conducted sensitivity analyses for the 1SMR approach using weighted median and weighted mode analyses. To do this, in each half of the sample, we generated summary statistics for each of the outcomes by regressing the exposure genetic variant genotypes onto the MR outcome phenotypes, adjusting for age, sex, the first 20 PCs, assessment centre and genotyping chip. We then used the effect estimates from these regressions for the outcome along with the split sample GWAS output for the exposure as summary statistics for the weighted median and weighted mode sensitivity MR analyses using the ‘TwoSampleMR’ R package ^8,9^. Where loneliness was our exposure, as this was partially overlapping with UK Biobank data, we also conducted 1SMR analyses using genetic variants from the published GWAS data but in unweighted genetic scores (i.e., all alleles assigned a weight of one) in an attempt to remove any bias from the weights being created in a largely overlapping sample. Using unweighted scores however assumes all genetic variants have equal influence and therefore we conducted these alongside the split sample analyses as an additional sensitivity analysis with the loneliness exposure. This additional analysis was not conducted for social isolation due to full sample overlap.

### 8. Two sample Mendelian Randomisation details and sensitivity analyses

Prior to analyses, as per guidance for the TwoSampleMR R package, we excluded genetic variants in linkage disequilibrium (LD) (using an r^2^ of 0.001, window of 10,000 kb and the European 1000 genomes reference panel). If the resulting exposure genetic variants were unavailable in the outcome GWAS data we attempted to identify proxy genetic variants using LDlink ^10^ at an LD r^2^ threshold of >0.8. Once the final genetic variant set was established we harmonised the exposure and outcome data so that the genetic variant effects for exposure and outcome were all in the same direction using the TwoSampleMR harmonise_data() function, which attempted to infer the positive strand based on allele frequencies and aligned palindromic genetic variants using allele frequencies where possible (or excluded them if it was unable to do this).

For the main analyses we used an approach called the inverse-variance weighted (IVW) method, which assumes that there is no bias due to horizontal pleiotropy (i.e., where genetic variants influence the outcome via pathways other than through the exposure) by fitting a regression for the genetic variant-exposure and genetic variant-outcome associations and constraining the intercept to pass through zero. We also conducted sensitivity analyses to assess the assumptions of 2SMR. These were MR-Egger ^11^, weighted median ^12^, MR using robust adjusted profile score (MR-RAPS) ^13^, MR Pleiotropy RESidual Sum and Outlier (MR-PRESSO) ^14^, MR lap ^15^ and MR-Causal Analysis Using Summary Effect estimates (CAUSE) ^16^. The MR-Egger estimate is valid even if horizontal pleiotropy is present, under the assumption that the pleiotropic effect of each genetic variant is independent of the strength of that genetic variant as an instrument, by allowing the intercept to not be constrained and allows the extent of a pleiotropic effect to be estimated if the intercept differs from zero. The weighted median method allows for up to 50% of the weight of the genetic variants used in the genetic instrument to be invalid. The MR-RAPS method uses a profile-likelihood function to downweigh outliers and assumes pleotropic effects are normally distributed and therefore provides an unbiased estimate even if there are weak instruments. The MR-PRESSO method can be used to detect and correct for horizontal pleiotropy by estimating the extent to which any outliers impact the effect estimate. MR lap accounts for any sample overlap and reduces bias which may occur when using overlapping samples in the exposure and outcome. MR-CAUSE can model both uncorrelated and correlated horizontal pleiotropy, avoiding false positives that may result from correlated pleiotropy. Correlated pleiotropy can occur where genetic variants influence both the exposure and outcome, for example via a heritable confounder.

To estimate the strength of our instruments we calculated first stage F-statistics, where a larger value indicated our instrument was strong enough for analyses and smaller values indicated likely weak instruments ^12^. All MR methods (other than MR RAPS) assume that all genetic variants are strong instruments (i.e., there is no measurement error in the genetic variant-exposure estimates) ^12^, known as the ‘NO Measurement Error’ (NOME) assumption. The extent to which the NOME assumption may be violated can be estimated using the F-statistic and the regression dilution I-squared statistics (in relation to MR-Egger), where lower values indicate greater violation. If violated, the IVW estimate may be prone to weak instrument bias and the MR-Egger estimate may be biased towards the null. We assessed the extent of any violation of the NOME assumption using the I-squared values, where an I-squared of less than 0.9 indicated that the MR-Egger estimates should be interpreted with caution due to regression dilution. Where this was the case, we additionally conducted simulation extrapolation (SIMEX) corrections as a sensitivity analysis. The SIMEX approach is a bias adjustment method that can be used when the NOME assumption may be violated, providing an estimate for the case where NOME has been satisfied. When the I-squared is less than 0.6 the SIMEX analysis may also be unreliable. Where loneliness and social isolation were the outcomes, for the exposures we used the genetic variants reported as genome-wide significant in the original publication, even when the full sample was not used (e.g., due to restrictions on the data available for depression and the three wellbeing GWAS, see **Supplementary Materials Table S2**), but weights of these genetic variants in the sub-sample were used. In cases where there were less than 5 genetic variants we used a less strict p-value threshold (1x10^-06^ where suicide attempt was the exposure and 1x10^-05^ where anxiety was the exposure). Finally, we conducted Steiger filtering ^17^ to further infer the most likely causal direction. This is achieved by calculating the variance explained in both the exposure and outcome by the genetic variants included in the instrument and assessing whether more variance is explained in the exposure than the outcome. Here we calculate the percentage of genetic variants that explain more variance in the exposure than the outcome. However, Steiger filtering is limited, in that measurement error, if present, can influence the inferred causal direction and provide inaccurate results, thus caution should be taken when interpreting these results.

### 9. Multivariable MR analyses

MVMR is an extension of MR that can be used to estimate the potential causal effects of multiple exposures (here loneliness and social isolation) on an outcome and assess whether effects of each exposure (conditioning on the other) are independent. Analyses were conducted using the ‘MVMR’ package in R ^18^ using a 2SMR approach. Conditional F-statistics were also calculated with this R package.

### Table S1. Phenotypic data in UK Biobank for observational/sibling control analyses and one sample Mendelian Randomisation analyses

| **Phenotype** | **Description** | **UK Biobank codes** | **Timepoint** |
| --- | --- | --- | --- |
| *Exposures* | | |  |
| Loneliness | Asked “Do you often feel lonely?” (Yes/No). Could also respond with do not know or prefer not to answer. | 2020 | Baseline between 2006 and 2010 |
| Social isolation | We used data from two measures:  1) Frequency of friend/family visits (“How often do you visit friends or family or have them visit you?”). Participants could respond with the following options: almost daily; 2-4 times a week; about once a week; about once a month; once every few months; never or almost never; no friends/family outside household. We combined the last two into the same category and applied the following values to each (with a higher value reflecting greater social isolation): 0, 0.2, 0.4, 0.6, 0.8 and 1, respectively.  2) Number of people in household (“Including yourself, how many people are living together in your household? (Include those who usually live in the house such as students living away from home during term, partners in the armed forces or professions such as pilots)”. Participants responded by providing a number. Answers less than one or more than 100 were rejected. Participants responding with an answer over 12 were asked to confirm. We coded responses as follows in a similar manner to the measure above: 1 in household (i.e., lived alone) = 1, 2 in household = 0.75, 3 in household = 0.5, 4 in household = 0.25, 5+ in household = 0.  We summed values for each measure to create a combined measure of social isolation. | 1031  709 | Baseline between 2006 and 2010 |
| *General health outcomes* | | |  |
| Hospital admissions | We also used HES for the number of hospital admissions for each participant in the same follow-up period as the QALYs measure. This was similar to the outcome used by Hazewinkel and colleagues ^19^. | HES admissions data | Follow-up until 30/09/2021 |
| Quality-adjusted life years (QALYs) | QALYs were estimated based on data from hospital episode statistics (HES) which have been linked to all participants up to 30/09/2021, although we only included data from date of assessment for UK Biobank. This is similar data to that used previously by Harrison and colleagues ^20^. Briefly, 240 health conditions (based on a study by Sullivan and colleagues ^21^ using ICD 9/10 codes) were used to predict QALYs for all participants from recruitment to the aforementioned date or in case of death. A value of 1 is indicative of a full year of perfect health whilst a value of 0 indicates no quality of life or death, although values can also be negative. To ensure consistency with the permitted value range of UK EQ-5D-3L value set, we coded it so that values over 1 were forced to have a value of 1 and values below -0.594 were forced to have a value of -0.594 ^22^. | HES data using ICD 9 and ICD 10 diagnosis codes for the 240 health conditions | Follow-up until 30/09/2021 |
| Multimorbidity | The presence of two or more self-reported chronic conditions (out of 35 possible conditions). See previous work by North and colleagues for further information about the phenotypes included ^23^. | Self-reported conditions as per North and colleagues work: alcohol dependency (20002 – 1408); anorexia/bulimia/other eating disorder diagnosis (20002 – 1470); atrial fibrillation (20002 – 1471, 1483); bronchiectasis (20002 – 1114); liver disease and hepatitis (20002 – 1155, 1156, 1157, 1158, 1579, 1580, 1604, 1506); chronic sinusitis (20002 – 1416, 1413, 1417); chronic obstructive pulmonary disease (20002 – 1112, 1113, 1472; and 6152); coronary heart disease (20002 – 1074, 1075, 1079, 1588, 1492, 1591, 1586, 1587; and 6150); dementia/Alzheimer's/cognitive impairment(20002 – 1263); diabetes (20002 – 1220, 1222, 1223, 1276, 1468, 1608; and 2443); diverticulitis (20002 – 1458); hearing loss (20002 – 1420, 1421; 2247; 3393; 2257); heart failure (20002 – 1076); hypertension (20002 – 1065, 1072; and 6150); inflammatory bowel disease (20002 – 1459, 1461, 1462, 1463); multiple sclerosis (20002 – 1261); Parkinson’s disease (20002 – 1262); peptic ulcer disease (20002 – 1400, 1457, 1142); peripheral vascular disease (20002 – 1067, 1087); prostate disorders (20002 – 1207, 1396, 1516, 1517); psychoactive substance misuse (20002 – 1409, 1410); rheumatoid arthritis, other inflammatory polyarthropathies and systemic connective tissue disorders (20002 – 1373, 1464, 1381, 1477, 1313, 1377, 1322, 1383, 1480, 1481, 1384, 1372); stroke and transient ischaemic attack (20002 – 1081, 1082, 1583, 1491; and 6150); thyroid disorders (20002 – 1225, 1226, 1522, 1610); constipation (20002 – 1599; and 6154); migraine (20002 – 1265); epilepsy (20002 – 1264); asthma (20002 – 1111; and 6152); irritable bowel syndrome (20002 – 1154); psoriasis or eczema (20002 – 1452, 1453, 1669); anxiety and other neurotic, stress related and somatoform disorders or depression (20002 – 1286, 1287, 1288, 1469, 1615; and 2090; 2100); cancer diagnosis in last 5 years (20001); chronic kidney disease (20002 – 1192, 1193, 1194, 1519, 1607); painful condition (6154); schizophrenia (and other psychosis) or bipolar disorder (20002 – 1289, 1291) | Baseline between 2006 and 2010 |
| Mortality | We looked at death as a binary outcome in the same follow-up period as the QALYs measure. Death was coded as occurring for this outcome if there was a date of death recorded. Where there was no date of death recorded, we assumed that this event had not occurred and the participant was coded as not having this outcome. | 40000 | Follow-up until 30/09/2021 |
| *Physical health outcomes* | | |  |
| Coronary artery disease (CAD) | CAD was defined similarly to the measures used in Patel and colleagues ^24^, but using linked data where we knew the dates of the event after their assessment visit. CAD was defined based on a combination of ICD 10, ICD 9, Office of Population Censuses and Surveys Classification of Interventions and Procedures, version 4 (OPCS4) and mortality data. Those with this event occurring prior to assessment were excluded from analyses (based on the same codes and also self-report data). | HES data using ICD 9 and ICD 10 diagnosis codes and OPCS4 procedure codes and mortality data. | Follow-up until 30/09/2021 |
| Heart failure | Heart failure was defined in a similar manner to that for CAD. Heart failure was defined based on a combination of ICD 10 and ICD 9 codes and mortality data. Those with this event occurring prior to assessment were excluded from analyses (based on the same codes and also self-report data). | HES data using ICD 9 and ICD 10 diagnosis codes and mortality data. | Follow-up until 30/09/2021 |
| Stroke | Stroke was defined in a similar manner to that for CAD. Stroke was defined based on a combination of ICD 10 and ICD 9 codes and mortality data. Those with this event occurring prior to assessment were excluded from analyses (based on the same codes and also self-report data). | HES data using ICD 9 and ICD 10 diagnosis codes and mortality data. | Follow-up until 30/09/2021 |
| Systolic blood pressure | Mean systolic blood pressure (mmHg) was calculated from two readings a few moments apart. These were either automated, manual or a combination of both. If only a single measure was obtained, we used this instead. Automated readings were taken using an Omron 705 IT electronic blood pressure monitor with a range of 0-255. A manual monitor was used in cases where the automated device could not be used. | 4080; 93 | Baseline between 2006 and 2010 |
| Type 2 Diabetes | A possible or probable diagnosis of type 2 diabetes was used. This was defined as per the Eastwood algorithm. Details on the Eastwood algorithm can be found in the paper describing it ^25^. However, to summarise, the algorithm is used to identify diabetes status using a combination of self-report, primary and secondary care data measures within UK Biobank. We used this to identify ‘possible’ and ‘probable’ cases of type 2 diabetes, where probable cases have stronger evidence of this being the case from primary care data and possible cases have weaker evidence. We also excluded individuals who had a possible or probable type 1 diabetes as per the Eastwood algorithm. | A combination of variables were used to derive this measure using the Eastwood algorithm: insulin, metformin, sulfonylureas, meglitinides, glitazones, non-metformin oral anti-diabetic, and other oral anti-diabetic medication from the UK Biobank self-reported medications list (20003); gestational diabetes, diabetes not otherwise specified, type 1 diabetes and type 2 diabetes from the self-reported non-cancer illness codes list (20002) and corresponding ages (20009); other diabetes related self-reported measures (2443, 4041; 2976; 6177; 6153; 2986) | Baseline between 2006 and 2010 |
| *Mental health outcomes* | | |  |
| Self-harm | Self-harm: Asked “Have you deliberately harmed yourself, whether or not you meant to end your life?”. Could respond with no, yes or prefer not to answer. We used yes/no data. | 20480 | Online questionnaire between 2016 and 2017 |
| Suicide attempts | Suicide attempt: Those you responded yes to the above question were then asked “Have you harmed yourself with the intention to end your life?". Similarly, they could respond with no, yes or prefer not to answer. We used yes/no data. | 20483 | Online questionnaire between 2016 and 2017 |
| Depression | Binary: Diagnosis of depression obtained from a combination of self-reported data and ICD10 codes.  Continuous trait: The Patient Health Questionnaire 9-question version (PHQ-9) | Binary outcome was obtained from a number of diagnosis variables: depression diagnosis from self-reported non-cancer illness code list (20002 - 1286 variable); ICD 10 codes from primary and secondary diagnoses lists (41202 - F32, F320, F321, F322, F323, F328, F329, F33, F330, F331, F332, F333, F334, F338, F339 and 41204 – same F codes) and a question in the online mental health questionnaire asking “Have you been diagnosed with one or more of the following mental health problems by a professional, even if you don't have it currently? (tick all that apply)” (20544).  The continuous outcome was obtained from the PHQ-9 (20514; 20510; 20517; 20519; 20511; 20507; 20508; 20518; 20513) | Baseline between 2006 and 2010 and online questionnaire between 2016 and 2017  Online questionnaire between 2016 and 2017 |
| Anxiety | Binary: Diagnosis of anxiety (all types) obtained from a combination of self-reported data and ICD10 codes.  Continuous trait: The Generalized Anxiety Disorder 7-question version (GAD-7-7) | Binary outcome was obtained from a number of diagnosis variables: anxiety diagnosis from self-reported non-cancer illness code list (20002 - 1287 variable); ICD 10 codes from primary and secondary diagnoses lists (41202 – F40, F400, F401, F402, F408, F409, F41, F410, F411, F412, F413, F418, F419 and 41204 – same F codes) and a question in the online mental health questionnaire asking “Have you been diagnosed with one or more of the following mental health problems by a professional, even if you don't have it currently? (tick all that apply)” (20544).  The continuous outcome was obtained from the GAD-7-7 (20506; 20509; 20520; 20515; 20516; 20505; 20512) | Baseline between 2006 and 2010 and online questionnaire between 2016 and 2017  Online questionnaire between 2016 and 2017 |
| Wellbeing | Two measures of wellbeing:  1) Positive affect* – Asked “In general how happy are you?”. Could respond with extremely happy, very happy, moderately happy, moderately unhappy, very unhappy, extremely unhappy, or they could respond with do not know or prefer not to answer. This was asked at both the initial assessment visit and in an online follow-up. Therefore, we have used data where this is available for a participant from the initial measure and if this is missing then we have used data from the follow-up where data is available.  2) Meaning in life – Asked “To what extent do you feel your life to be meaningful?”. Could respond with not at all, a little, a moderate amount, very much, an extreme amount or they could respond with do not know or prefer not to answer. | 4526; 20458  20460 | Baseline between 2006 and 2010 and online questionnaire between 2016 and 2017  Online questionnaire between 2016 and 2017 |

**Here we have labelled this measure as positive affect in line with the variables included in the positive affect GWAS, although note that happiness is not the same as positive affect more broadly*

### Table S2. Genome-wide association study (GWAS) data for Mendelian Randomisation (MR) analyses

| **Phenotype** | **Description** | **Author** | **Number of individuals** | **Number of independent genome-wide significant genetic variants** |
| --- | --- | --- | --- | --- |
| *Exposures* | | | | |
| Loneliness | A binary measure of loneliness was used for one of the cohorts (UKBB) and a continuous measures used for the other 6 cohorts. The binary measure was from a question asking ‘Do you feel lonely?’. The continuous measure used varied across cohorts with 2 including a 3-item questionnaire^a^, 3 asking ‘Did you feel lonely during the past week’ with a 4-point scale and another including a 9-item questionnaire^b^. | Abdellaoui et al., 2019 ^26^ | 573,604 obtained from 7 different cohorts (including UK Biobank). Where loneliness was the outcome 23andMe data were excluded (N=20,591) as only the top 10,000 genetic variants are available in the full sample. | 16 |
| Social isolation | We used data from a GWAS we have conducted in UK Biobank using a combination of two measures (same as in observational analyses):  1) Frequency of friend/family visits (“How often do you visit friends or family or have them visit you?”). Participants could respond with the following options: almost daily; 2-4 times a week; about once a week; about once a month; once every few months; never or almost never; no friends/family outside household. We combined the last two into the same category and applied the following values to each (with a higher value reflecting greater social isolation): 0, 0.2, 0.4, 0.6, 0.8 and 1, respectively.  2) Number of people in household (“Including yourself, how many people are living together in your household? (Include those who usually live in the house such as students living away from home during term, partners in the armed forces or professions such as pilots)”. Participants responded by providing a number. Answers less than one or more than 100 were rejected. Participants responding with an answer over 12 were asked to confirm. We coded responses as follows in a similar manner to the measure above: 1 in household (i.e., lived alone) = 1, 2 in household = 0.75, 3 in household = 0.5, 4 in household = 0.25, 5+ in household = 0.  We summed values for each measure to create a combined measure of social isolation.  Details of the GWAS are provided in the Supplementary Materials. | NA – see Supplementary Materials Section 2 | 457,086 from UK Biobank | 15 |
| *Physical health outcomes* | | | | |
| Coronary artery disease (CAD) | We used data from a predominantly European GWAS of CAD (from a mixture of sources including ICD9/10 codes and hospital records). | Aragam et al., 2022 ^27^ | Meta-analysis of 181,522 cases and 1,165,690 controls across 10 cohorts (including UK Biobank) | 241 |
| Heart failure | We used data from a European GWAS of heart failure. Cases of heart failure included participants with a clinical diagnosis of heart failure, of any aetiology. Definitions across the different studies included a combination of incidence and prevalence cases and are described in further detail in the GWAS paper. | Shah et al., 2020 ^28^ | 47,309 cases and 930,014 across 26 cohorts (including UK Biobank) | 11 |
| Stroke | We used data from a GWAS of ‘any stroke’ which included ischaemic stroke, stroke of unknown or undetermined type, any ischaemic stroke regardless of subtype and ischaemic stroke subtypes. | Mishra et al., 2022 ^29^ | 73,652 cases and 1,234,808 controls from 44 European cohorts (including UK Biobank) | 29 in European sample |
| Systolic blood pressure | We used data from a European GWAS of systolic blood (details of the individual cohort measurements can be found in the GWAS paper). | Evangelou et al., 2018 ^30^ | Meta-analysis of 757,601 participants across 77 cohorts (including UK Biobank) | 130 |
| Type 2 diabetes | Case-control for type 2 diabetes diagnosis. | Vujkovic et al., 2020 ^31^ | 148,726 cases and 965,732 controls of European ancestry (including UK Biobank) | 425 in European sample |
| *Mental health outcomes* | | | | |
| Suicide attempts | Case-control for suicide attempt in European subsample. | Mullins et al., 2022 ^32^ | 26,590 cases and 492,022 controls (European sample) from the International Suicide Genetics Consortium (ISGC) (including UK Biobank) | 2 in European sample |
| Depression | Case-control for major depressive disorder in a European sample GWAS. | Wray et al., 2018 ^33^ | 45,396 cases and 97,250 controls (excluding 23andMe and UK Biobank) | 44 |
| Anxiety | Used data from five core anxiety disorders: generalised anxiety disorder, panic disorder, social phobia, agoraphobia and specific phobias. From this, a case-control phenotype was derived and used in a European sample GWAS. | Otowa et al., 2016 ^34^ | 5,761 cases and 11,765 controls from 7 cohorts (including some sample overlap with loneliness GWAS) | 1 |
| Wellbeing | Three GWAS were used (from European sample):  1) Positive affect  2) Life satisfaction  3) Wellbeing spectrum encompassing measures of life satisfaction, positive affect, neuroticism and depressive symptoms (N GWAMA version). | Baselmans et al., ^35^ | 2,083,151 observations excluding 23andMe | 1) 191  2) 148  3) 231 |

*^a^3-item questionnaire asked the following with a. 3-point scale:* *1) How often do you feel left out?, 2) How often do you feel isolated from others?, 3) How often do you feel that you lack companionship?*

*^b^9-item questionnaire asked the following with a 4-point scale response: 1) I feel in tune with the people around me, 2) There are people I can turn to, 3) I feel alone, 4) I feel part of a group of friends, 5) I have a lot in common with the people around me, 6) I feel isolated from others, 7) There are people who really understand me, 8) I am unhappy being so withdrawn, 9) There are people I can talk to.*

### Table S3. Independent genome-wide significant genetic variants from social isolation^a^ genome-wide association study in UK Biobank

| **Genetic variant** | **Chromosome** | **Effect allele** | **Other allele** | **EAF** | **Beta** | **SE** | **P** |
| --- | --- | --- | --- | --- | --- | --- | --- |
| rs35393280 | 20 | A | C | 0.75 | -0.006 | 0.0008 | 1.90x10^-15^ |
| rs10099728 | 8 | C | T | 0.82 | 0.007 | 0.0009 | 5.00x10^-15^ |
| rs2044725 | 3 | C | T | 0.35 | -0.005 | 0.0007 | 7.50x10^-13^ |
| rs2568955 | 1 | T | C | 0.26 | -0.005 | 0.0008 | 1.70x10^-10^ |
| rs2188541 | 7 | A | G | 0.37 | 0.004 | 0.0007 | 1.00x10^-09^ |
| rs4587178 | 6 | T | C | 0.62 | 0.004 | 0.0007 | 3.00x10^-09^ |
| rs12155844 | 8 | T | C | 0.81 | 0.005 | 0.0009 | 4.90x10^-09^ |
| rs112221661 | 17 | A | G | 0.30 | 0.004 | 0.0008 | 5.50x10^-09^ |
| rs1421699 | 5 | G | A | 0.35 | 0.004 | 0.0007 | 7.60x10^-09^ |
| rs17499351 | 5 | T | C | 0.95 | -0.008 | 0.0015 | 1.60x10^-08^ |
| rs9907261 | 17 | C | T | 0.49 | 0.004 | 0.0007 | 1.90x10^-08^ |
| rs2410941 | 5 | C | A | 0.47 | -0.004 | 0.0007 | 2.00x10^-08^ |
| rs2721940 | 8 | A | C | 0.40 | -0.004 | 0.0007 | 3.40x10^-08^ |
| rs1851919 | 3 | C | T | 0.43 | 0.004 | 0.0007 | 3.80x10^-08^ |
| rs6691624 | 1 | C | T | 0.34 | -0.004 | 0.0007 | 4.30x10^-08^ |

*EAF=effect allele frequency, SE=standard error*

^a^Social isolation was measured on a scale from 0 to 2 based on household size and number of visits to/from friends or family (Supplementary Table S2)

### Table S4. Potential confounders in UK Biobank for observational/sibling control analyses and one sample Mendelian Randomisation analyses

| **Phenotype** | **Description** | **UK Biobank codes** |
| --- | --- | --- |
| Age | Derived from date of birth and date of attending assessment centre, rounded to a whole year and modelled as a continuous variable. | 21003 |
| Sex | Obtained from NHS records (i.e., sex assigned at birth), but may have been updated by the participant as self-reported sex. (Male or female) | 31 |
| Assessment centre | The assessment centre that the participant attended. (Categorical, 22 categories) | 54 |
| Townsend deprivation index | Calculated based on postcode from the preceding national census output areas. (Continuous variable) | 22189 |
| Education | Asked “Which of the following qualifications do you have?”. Could respond with College or University degree; A levels/AS levels or equivalent; O levels/GCSEs or equivalent; CSEs or equivalent; NVQ or HND or HNC or equivalent; Other professional qualifications e.g., nursing, teaching; None of the above; Prefer not to answer. They could select multiple responses.  A/AS levels are UK subject specific qualifications typically completed over 2 years between the ages of 16 and 18. ‘GCSEs (General Certificate of Secondary Education) or O levels are UK subject specific qualifications typically completed over 3 years towards the end of secondary school education.  We converted this to years of education as per Okbay et al ^36^ and modelled it as a continuous variable. | 6138 |
| Income | Asked “What is the average total income before tax received by your household?”. Could respond with less than 18,000; 18,000 to 30,999; 31,000 to 51,999; 52,000 to 100,000, greater than 100,000; Do not know; Prefer not to answer. | 738 |
| Ethnicity | Self-reported responses were from touchscreen questionnaire questions asking, ‘What is your ethnic group?’ Could respond: White; Mixed; Asian or Asian British; Black or Black British, Chinese, Other ethnic group, Do not know; Prefer not to answer. Branching questions were then asked from this.  We coded this as 4 groups. | 21000 |
| Disability | Asked “Do you have any long-standing illness, disability or infirmity?” (Yes/No). Could also respond with do not know or prefer not to answer. | 2188 |
| Adverse childhood events | In the mental well-being online questionnaire participants were asked each of the following questions:  1) "When I was growing up: People in my family hit me so hard that it left me with bruises or marks."  2) "When I was growing up: Someone molested me (sexually)."  3) "When I was growing up: I felt that someone in my family hated me."  4) "When I was growing up: I felt loved."  5) "When I was growing up: There was someone to take me to the doctor if I needed it."  They could respond: never true; rarely true; sometimes true; often true; very often true; prefer not to answer.  We combined these into a single measure, summing whether each of the above was present (1 for present, 0 for absent) as per Soares et al^37^. The first three were considered present if they responded as rarely true or above. The fourth was considered present if they reported as never true or rarely true. The fifth was considered present if they responded with sometimes true or below. The combined measure was modelled as a continuous variable. | 29077; 29079; 29078; 29076; 29080 |
| Principal components | We used the first 20 principal components (PCs) from PC analysis of genotype data. (Continuous variables) | Derived as described previously ^2^. |
| Genotyping chip | Two genotyping chips were used in UK Biobank; the UKBB axiom array (90% of participants were genotyped with this) and the UK BiLEVE array. (Categorical, 2 categories) | 22000 |

### Table S5. Data availability for the genome-wide association studies (GWAS) we used

| **Phenotype** | **Data access information** |
| --- | --- |
| Loneliness | GWAS summary statistics excluding 23andMe can be downloaded from <https://t.co/ARgS84uwKl>. Data for the top 10,000 genetic variants including 23andMe are available from the authors at request. |
| Coronary artery disease (CAD) | GWAS summary statistics are available here: <http://ftp.ebi.ac.uk/pub/databases/gwas/summary_statistics/GCST90132001-GCST90133000/GCST90132314/> with accession GCST90132314 |
| Heart failure | GWAS summary statistics are available here: <https://cvd.hugeamp.org/dinspector.html?dataset=GWAS_HERMES_eu>. |
| Stroke | Details on the GWAS can be found here: <https://www.megastroke.org/>. For the European subset we contacted the authors of the paper. |
| Systolic blood pressure | GWAS summary statistics are available here: <https://www.ebi.ac.uk/gwas/publications/30224653> with accession GCST006624. |
| Type 2 diabetes | GWAS summary statistics are available on request from dbGaP (  T2D: Phs001672; Pha004945; T2D.EUR.MVP_Penn_DIAMANTE_Malmo.NatGen2020) |
| Suicide attempts | The GWAS summary statistics can be requested here: <https://docs.google.com/forms/d/e/1FAIpQLSc4hGOJ181WGfP3yTjMxUmdf4d6hK0dkXSwRO0ivcPdZIOslg/viewform>. |
| Depression | The GWAS summary statistics are available to download from the Psychiatric Genomics Consortium (PGC) (<https://figshare.com/articles/dataset/mdd2018/14672085>) after agreeing to abide by their data access conditions. |
| Anxiety | The GWAS summary statistics are available to download from the PGC (<https://pgc.unc.edu/for-researchers/download-results/?choice=Other+GWAS+DataAnxiety+Neuro+Genetics+Study+%2528ANGST%2529>) after agreeing to abide by their data access conditions. |
| Wellbeing | The GWAS summary statistics excluding 23andMe are available to download from: <https://surfdrive.surf.nl/files/index.php/s/Ow1qCDpFT421ZOO?path=%2FMultivariate_GWAMA_sumstats>. |

### Table S6. Observational analysis results

| **Exposure** | **Outcome** | **Model** | **Level of adjustment** | **N** | **HR, OR or mean difference (95% CI)** | **p-value** | **Proportional hazards test (CoxPH)** |
| --- | --- | --- | --- | --- | --- | --- | --- |
| Loneliness | CAD | CoxPH | unadjusted | 392,132 | 1.27 (1.23 to 1.31) | 9.77e-57 | 5.06e-11 |
| Loneliness | CAD | CoxPH | adjusted1^a^ | 321,985 | 1.26 (1.21 to 1.30) | 1.86e-39 | 2.07e-05 |
| Loneliness | CAD | CoxPH | adjusted1 with disability^a^ | 315,593 | 1.20 (1.16 to 1.24) | 4.59e-24 | 1.65e-04 |
| Loneliness | CAD | CoxPH | adjusted1 with ACEs^a^ | 117,412 | 1.14 (1.06 to 1.23) | 7.81e-04 | 0.37 |
| Loneliness | CAD | CoxPH | adjusted2^a^ | 115,557 | 1.10 (1.02 to 1.19) | 0.02 | 0.61 |
| Loneliness | HF | CoxPH | unadjusted | 412,220 | 1.54 (1.48 to 1.60) | 8.89e-93 | 5.13e-08 |
| Loneliness | HF | CoxPH | adjusted1^a^ | 337,606 | 1.52 (1.45 to 1.59) | 3.65e-64 | 6.94e-05 |
| Loneliness | HF | CoxPH | adjusted1 with disability^a^ | 330,811 | 1.40 (1.34 to 1.47) | 2.11e-41 | 9.19e-04 |
| Loneliness | HF | CoxPH | adjusted1 with ACEs^a^ | 120,617 | 1.40 (1.21 to 1.61) | 5.75e-06 | 0.68 |
| Loneliness | HF | CoxPH | adjusted2^a^ | 118,693 | 1.29 (1.12 to 1.50) | 6.34e-04 | 0.53 |
| Loneliness | Stroke | CoxPH | unadjusted | 407,082 | 1.36 (1.29 to 1.44) | 3.78e-30 | 8.37e-04 |
| Loneliness | Stroke | CoxPH | adjusted1^a^ | 333,578 | 1.37 (1.28 to 1.45) | 1.09e-22 | 8.51e-03 |
| Loneliness | Stroke | CoxPH | adjusted1 with disability^a^ | 326,922 | 1.32 (1.24 to 1.41) | 4.18e-18 | 0.03 |
| Loneliness | Stroke | CoxPH | adjusted1 with ACEs^a^ | 119,791 | 1.13 (0.95 to 1.36) | 0.17 | 0.23 |
| Loneliness | Stroke | CoxPH | adjusted2^a^ | 117,889 | 1.11 (0.92 to 1.33) | 0.29 | 0.27 |
| Loneliness | Death | CoxPH | unadjusted | 414,431 | 1.44 (1.40 to 1.48) | 6.77e-153 | 1.53e-15 |
| Loneliness | Death | CoxPH | adjusted1^a^ | 339,324 | 1.41 (1.36 to 1.45) | 7.27e-98 | 7.84e-10 |
| Loneliness | Death | CoxPH | adjusted2^a,b^ | 332,507 | 1.34 (1.30 to 1.39) | 1.03e-70 | 9.99e-08 |
| Loneliness | Multimorbidity | logistic | unadjusted | 414,329 | 2.25 (2.21 to 2.29) | 0.00 |  |
| Loneliness | Multimorbidity | logistic | adjusted1 | 339,253 | 2.14 (2.10 to 2.18) | 0.00 |  |
| Loneliness | Multimorbidity | logistic | adjusted1 with disability | 332,439 | 1.98 (1.94 to 2.02) | 0.00 |  |
| Loneliness | Multimorbidity | logistic | adjusted1 with ACEs | 120,875 | 1.94 (1.87 to 2.00) | 0.00 |  |
| Loneliness | Multimorbidity | logistic | adjusted2 | 118,947 | 1.85 (1.78 to 1.91) | 7.30e-250 |  |
| Loneliness | T2D | logistic | unadjusted | 414,432 | 1.45 (1.40 to 1.49) | 3.25e-102 |  |
| Loneliness | T2D | logistic | adjusted1 | 339,325 | 1.41 (1.35 to 1.46) | 2.92e-61 |  |
| Loneliness | T2D | logistic | adjusted1 with disability | 332,508 | 1.15 (1.10 to 1.20) | 2.43e-10 |  |
| Loneliness | T2D | logistic | adjusted1 with ACEs | 120,893 | 1.25 (1.13 to 1.38) | 1.25e-05 |  |
| Loneliness | T2D | logistic | adjusted2 | 118,965 | 1.04 (0.93 to 1.15) | 0.50 |  |
| Loneliness | Hospital admissions | linear | unadjusted | 414,432 | 0.86 (0.74 to 0.97) | 2.52e-48 |  |
| Loneliness | Hospital admissions | linear | adjusted1 | 339,325 | 0.70 (0.58 to 0.81) | 8.52e-33 |  |
| Loneliness | Hospital admissions | linear | adjusted1 with disability | 332,508 | 0.44 (0.32 to 0.55) | 5.46e-13 |  |
| Loneliness | Hospital admissions | linear | adjusted1 with ACEs | 120,893 | 0.44 (0.34 to 0.55) | 9.61e-17 |  |
| Loneliness | Hospital admissions | linear | adjusted2 | 118,965 | 0.33 (0.22 to 0.43) | 7.98e-10 |  |
| Loneliness | SBP | linear | unadjusted | 413,588 | -2.92 (-3.06 to -2.77) | 0.00 |  |
| Loneliness | SBP | linear | adjusted1 | 338,736 | -1.61 (-1.76 to -1.45) | 1.85e-94 |  |
| Loneliness | SBP | linear | adjusted1 with disability | 331,948 | -1.55 (-1.71 to -1.40) | 2.76e-85 |  |
| Loneliness | SBP | linear | adjusted1 with ACEs | 120,801 | -1.54 (-1.81 to -1.28) | 2.79e-30 |  |
| Loneliness | SBP | linear | adjusted2 | 118,876 | -1.55 (-1.82 to -1.28) | 7.57e-30 |  |
| Loneliness | QALYs | linear | unadjusted | 336,799 | -0.05 (-0.05 to -0.05) | 0.00 |  |
| Loneliness | QALYs | linear | adjusted1 | 273,910 | -0.04 (-0.04 to -0.04) | 1.98e-323 |  |
| Loneliness | QALYs | linear | adjusted1 with disability | 268,055 | -0.03 (-0.03 to -0.02) | 4.04e-166 |  |
| Loneliness | QALYs | linear | adjusted1 with ACEs | 94,950 | -0.02 (-0.02 to -0.02) | 9.45e-59 |  |
| Loneliness | QALYs | linear | adjusted2 | 93,307 | -0.01 (-0.02 to -0.01) | 3.72e-32 |  |
| Loneliness | Self-harm | logistic | unadjusted | 131,523 | 3.01 (2.85 to 3.19) | 0.00 |  |
| Loneliness | Self-harm | logistic | adjusted1 | 114,019 | 2.43 (2.28 to 2.58) | 5.38e-171 |  |
| Loneliness | Self-harm | logistic | adjusted1 with disability | 112,132 | 2.28 (2.14 to 2.43) | 1.27e-141 |  |
| Loneliness | Self-harm | logistic | adjusted1 with ACEs | 80,247 | 1.96 (1.81 to 2.12) | 8.26e-64 |  |
| Loneliness | Self-harm | logistic | adjusted2 | 79,000 | 1.86 (1.72 to 2.02) | 6.33e-53 |  |
| Loneliness | Suicide attempt | logistic | unadjusted | 131,343 | 3.38 (3.13 to 3.64) | 2.07e-220 |  |
| Loneliness | Suicide attempt | logistic | adjusted1 | 113,863 | 2.62 (2.41 to 2.84) | 9.77e-114 |  |
| Loneliness | Suicide attempt | logistic | adjusted1 with disability | 111,982 | 2.41 (2.21 to 2.62) | 2.92e-91 |  |
| Loneliness | Suicide attempt | logistic | adjusted1 with ACEs | 80,147 | 2.02 (1.82 to 2.25) | 1.29e-38 |  |
| Loneliness | Suicide attempt | logistic | adjusted2 | 78,904 | 1.89 (1.70 to 2.11) | 8.76e-31 |  |
| Loneliness | Depression diagnosis | logistic | unadjusted | 414,432 | 3.15 (3.09 to 3.21) | 0.00 |  |
| Loneliness | Depression diagnosis | logistic | adjusted1 | 339,325 | 2.79 (2.73 to 2.85) | 0.00 |  |
| Loneliness | Depression diagnosis | logistic | adjusted1 with disability | 332,508 | 2.63 (2.57 to 2.69) | 0.00 |  |
| Loneliness | Depression diagnosis | logistic | adjusted1 with ACEs | 120,893 | 2.63 (2.53 to 2.72) | 0.00 |  |
| Loneliness | Depression diagnosis | logistic | adjusted2 | 118,965 | 2.54 (2.44 to 2.63) | 0.00 |  |
| Loneliness | Anxiety diagnosis | logistic | unadjusted | 414,432 | 2.15 (2.11 to 2.21) | 0.00 |  |
| Loneliness | Anxiety diagnosis | logistic | adjusted1 | 339,325 | 1.97 (1.92 to 2.03) | 0.00 |  |
| Loneliness | Anxiety diagnosis | logistic | adjusted1 with disability | 332,508 | 1.88 (1.83 to 1.93) | 0.00 |  |
| Loneliness | Anxiety diagnosis | logistic | adjusted1 with ACEs | 120,893 | 1.96 (1.88 to 2.05) | 2.10e-205 |  |
| Loneliness | Anxiety diagnosis | logistic | adjusted2 | 118,965 | 1.90 (1.82 to 1.98) | 1.73e-180 |  |
| Loneliness | Depression trait | linear | unadjusted | 129,518 | 2.90 (2.82 to 2.97) | 0.00 |  |
| Loneliness | Depression trait | linear | adjusted1 | 112,498 | 2.58 (2.50 to 2.66) | 0.00 |  |
| Loneliness | Depression trait | linear | adjusted1 with disability | 110,678 | 2.46 (2.39 to 2.54) | 0.00 |  |
| Loneliness | Depression trait | linear | adjusted1 with ACEs | 79,432 | 2.23 (2.14 to 2.31) | 0.00 |  |
| Loneliness | Depression trait | linear | adjusted2 | 78,213 | 2.14 (2.05 to 2.22) | 0.00 |  |
| Loneliness | Anxiety trait | linear | unadjusted | 130,109 | 2.24 (2.17 to 2.30) | 0.00 |  |
| Loneliness | Anxiety trait | linear | adjusted1 | 112,969 | 1.96 (1.89 to 2.03) | 0.00 |  |
| Loneliness | Anxiety trait | linear | adjusted1 with disability | 111,146 | 1.89 (1.82 to 1.96) | 0.00 |  |
| Loneliness | Anxiety trait | linear | adjusted1 with ACEs | 79,741 | 1.65 (1.57 to 1.73) | 0.00 |  |
| Loneliness | Anxiety trait | linear | adjusted2 | 78,513 | 1.60 (1.52 to 1.68) | 0.00 |  |
| Loneliness | Happy | linear | unadjusted | 225,943 | -0.61 (-0.62 to -0.61) | 0.00 |  |
| Loneliness | Happy | linear | adjusted1 | 189,685 | -0.57 (-0.58 to -0.56) | 0.00 |  |
| Loneliness | Happy | linear | adjusted1 with disability | 186,126 | -0.56 (-0.57 to -0.55) | 0.00 |  |
| Loneliness | Happy | linear | adjusted1 with ACEs | 94,265 | -0.53 (-0.54 to -0.51) | 0.00 |  |
| Loneliness | Happy | linear | adjusted2 | 92,761 | -0.52 (-0.53 to -0.51) | 0.00 |  |
| Loneliness | Meaning in life | linear | unadjusted | 128,845 | -0.51 (-0.52 to -0.49) | 0.00 |  |
| Loneliness | Meaning in life | linear | adjusted1 | 111,843 | -0.48 (-0.49 to -0.46) | 0.00 |  |
| Loneliness | Meaning in life | linear | adjusted1 with disability | 110,019 | -0.47 (-0.48 to -0.45) | 0.00 |  |
| Loneliness | Meaning in life | linear | adjusted1 with ACEs | 78,790 | -0.44 (-0.46 to -0.42) | 0.00 |  |
| Loneliness | Meaning in life | linear | adjusted2 | 77,577 | -0.43 (-0.45 to -0.41) | 0.00 |  |
| Social isolation | CAD | CoxPH | unadjusted | 390,606 | 1.13 (1.09 to 1.18) | 5.30e-10 | 7.28e-03 |
| Social isolation | CAD | CoxPH | adjusted1^a^ | 325,212 | 0.95 (0.91 to 0.99) | 0.02 | 0.03 |
| Social isolation | CAD | CoxPH | adjusted1 with disability^a^ | 318,510 | 0.94 (0.90 to 0.98) | 7.18e-03 | 0.16 |
| Social isolation | CAD | CoxPH | adjusted1 with ACEs^a^ | 118,431 | 0.84 (0.77 to 0.92) | 1.14e-04 | 0.84 |
| Social isolation | CAD | CoxPH | adjusted2^a^ | 116,512 | 0.84 (0.77 to 0.92) | 1.75e-04 | 0.50 |
| Social isolation | HF | CoxPH | unadjusted | 410,475 | 1.43 (1.35 to 1.51) | 1.83e-32 | 6.44e-04 |
| Social isolation | HF | CoxPH | adjusted1^a^ | 341,000 | 1.15 (1.08 to 1.23) | 1.55e-05 | 0.03 |
| Social isolation | HF | CoxPH | adjusted1 with disability^a^ | 333,885 | 1.15 (1.07 to 1.23) | 4.06e-05 | 0.12 |
| Social isolation | HF | CoxPH | adjusted1 with ACEs^a^ | 121,662 | 1.01 (0.85 to 1.20) | 0.91 | 0.29 |
| Social isolation | HF | CoxPH | adjusted2^a^ | 119,673 | 1.00 (0.84 to 1.19) | 0.98 | 0.45 |
| Social isolation | Stroke | CoxPH | unadjusted | 405,408 | 1.22 (1.13 to 1.31) | 7.91e-08 | 0.02 |
| Social isolation | Stroke | CoxPH | adjusted1^a^ | 336,937 | 1.03 (0.95 to 1.12) | 0.43 | 0.20 |
| Social isolation | Stroke | CoxPH | adjusted1 with disability^a^ | 329,964 | 1.05 (0.96 to 1.13) | 0.29 | 0.37 |
| Social isolation | Stroke | CoxPH | adjusted1 with ACEs^a^ | 120,833 | 0.95 (0.77 to 1.17) | 0.64 | 0.45 |
| Social isolation | Stroke | CoxPH | adjusted2^a^ | 118,866 | 0.97 (0.78 to 1.19) | 0.74 | 0.42 |
| Social isolation | Death | CoxPH | unadjusted | 412,649 | 1.69 (1.63 to 1.76) | 3.50e-165 | 3.98e-11 |
| Social isolation | Death | CoxPH | adjusted1^a^ | 342,733 | 1.34 (1.29 to 1.40) | 1.20e-42 | 2.59e-05 |
| Social isolation | Death | CoxPH | adjusted2^a,b^ | 335,593 | 1.34 (1.29 to 1.40) | 1.04e-41 | 3.30e-03 |
| Social isolation | Multimorbidity | logistic | unadjusted | 412,549 | 1.45 (1.43 to 1.48) | 0.00 |  |
| Social isolation | Multimorbidity | logistic | adjusted1 | 342,661 | 1.07 (1.05 to 1.10) | 4.08e-11 |  |
| Social isolation | Multimorbidity | logistic | adjusted1 with disability | 335,524 | 1.04 (1.02 to 1.07) | 5.76e-04 |  |
| Social isolation | Multimorbidity | logistic | adjusted1 with ACEs | 121,922 | 1.06 (1.02 to 1.09) | 3.51e-03 |  |
| Social isolation | Multimorbidity | logistic | adjusted2 | 119,929 | 1.05 (1.01 to 1.09) | 0.02 |  |
| Social isolation | T2D | logistic | unadjusted | 412,650 | 1.71 (1.64 to 1.78) | 5.01e-132 |  |
| Social isolation | T2D | logistic | adjusted1 | 342,734 | 1.12 (1.06 to 1.18) | 1.80e-05 |  |
| Social isolation | T2D | logistic | adjusted1 with disability | 335,594 | 1.07 (1.01 to 1.13) | 0.01 |  |
| Social isolation | T2D | logistic | adjusted1 with ACEs | 121,940 | 1.25 (1.11 to 1.41) | 2.24e-04 |  |
| Social isolation | T2D | logistic | adjusted2 | 119,947 | 1.22 (1.08 to 1.38) | 1.56e-03 |  |
| Social isolation | Hospital admissions | linear | unadjusted | 412,650 | 1.10 (0.99 to 1.21) | 5.07e-85 |  |
| Social isolation | Hospital admissions | linear | adjusted1 | 342,734 | -0.12 (-0.25 to 0.02) | 0.09 |  |
| Social isolation | Hospital admissions | linear | adjusted1 with disability | 335,594 | -0.19 (-0.32 to -0.05) | 5.70e-03 |  |
| Social isolation | Hospital admissions | linear | adjusted1 with ACEs | 121,940 | -0.28 (-0.39 to -0.18) | 6.45e-08 |  |
| Social isolation | Hospital admissions | linear | adjusted2 | 119,947 | -0.30 (-0.41 to -0.20) | 9.19e-09 |  |
| Social isolation | SBP | linear | unadjusted | 411,812 | 4.60 (4.43 to 4.77) | 0.00 |  |
| Social isolation | SBP | linear | adjusted1 | 342,135 | 0.11 (-0.07 to 0.29) | 0.23 |  |
| Social isolation | SBP | linear | adjusted1 with disability | 335,027 | 0.13 (-0.05 to 0.31) | 0.14 |  |
| Social isolation | SBP | linear | adjusted1 with ACEs | 121,844 | 0.44 (0.15 to 0.73) | 2.76e-03 |  |
| Social isolation | SBP | linear | adjusted2 | 119,854 | 0.46 (0.17 to 0.76) | 1.89e-03 |  |
| Social isolation | QALYs | linear | unadjusted | 334,902 | -0.06 (-0.06 to -0.06) | 0.00 |  |
| Social isolation | QALYs | linear | adjusted1 | 276,687 | -4.44e-03 (-6.58e-03 to -2.31e-03) | 4.38e-05 |  |
| Social isolation | QALYs | linear | adjusted1 with disability | 270,553 | -5.16e-04 (-2.56e-03 to 1.53e-03) | 0.62 |  |
| Social isolation | QALYs | linear | adjusted1 with ACEs | 95,783 | 5.20e-03 (2.72e-03 to 7.68e-03) | 4.04e-05 |  |
| Social isolation | QALYs | linear | adjusted2 | 94,086 | 6.16e-03 (3.79e-03 to 8.54e-03) | 3.62e-07 |  |
| Social isolation | Self-harm | logistic | unadjusted | 131,455 | 1.49 (1.38 to 1.62) | 8.08e-23 |  |
| Social isolation | Self-harm | logistic | adjusted1 | 114,972 | 1.61 (1.48 to 1.75) | 2.06e-28 |  |
| Social isolation | Self-harm | logistic | adjusted1 with disability | 113,013 | 1.57 (1.44 to 1.71) | 8.27e-25 |  |
| Social isolation | Self-harm | logistic | adjusted1 with ACEs | 80,890 | 1.36 (1.23 to 1.50) | 4.62e-09 |  |
| Social isolation | Self-harm | logistic | adjusted2 | 79,596 | 1.33 (1.20 to 1.48) | 5.03e-08 |  |
| Social isolation | Suicide attempt | logistic | unadjusted | 131,270 | 1.90 (1.70 to 2.12) | 8.18e-30 |  |
| Social isolation | Suicide attempt | logistic | adjusted1 | 114,810 | 1.76 (1.56 to 1.97) | 4.21e-21 |  |
| Social isolation | Suicide attempt | logistic | adjusted1 with disability | 112,857 | 1.69 (1.50 to 1.90) | 3.05e-18 |  |
| Social isolation | Suicide attempt | logistic | adjusted1 with ACEs | 80,791 | 1.39 (1.20 to 1.60) | 8.34e-06 |  |
| Social isolation | Suicide attempt | logistic | adjusted2 | 79,500 | 1.35 (1.17 to 1.56) | 5.07e-05 |  |
| Social isolation | Depression diagnosis | logistic | unadjusted | 412,650 | 1.42 (1.38 to 1.45) | 1.13e-151 |  |
| Social isolation | Depression diagnosis | logistic | adjusted1 | 342,734 | 1.38 (1.34 to 1.42) | 4.98e-98 |  |
| Social isolation | Depression diagnosis | logistic | adjusted1 with disability | 335,594 | 1.35 (1.31 to 1.39) | 7.96e-81 |  |
| Social isolation | Depression diagnosis | logistic | adjusted1 with ACEs | 121,940 | 1.24 (1.18 to 1.30) | 1.84e-18 |  |
| Social isolation | Depression diagnosis | logistic | adjusted2 | 119,947 | 1.23 (1.17 to 1.29) | 6.06e-17 |  |
| Social isolation | Anxiety diagnosis | logistic | unadjusted | 412,650 | 1.30 (1.26 to 1.34) | 7.01e-62 |  |
| Social isolation | Anxiety diagnosis | logistic | adjusted1 | 342,734 | 1.26 (1.22 to 1.31) | 2.49e-36 |  |
| Social isolation | Anxiety diagnosis | logistic | adjusted1 with disability | 335,594 | 1.24 (1.20 to 1.29) | 7.31e-31 |  |
| Social isolation | Anxiety diagnosis | logistic | adjusted1 with ACEs | 121,940 | 1.12 (1.06 to 1.19) | 6.19e-05 |  |
| Social isolation | Anxiety diagnosis | logistic | adjusted2 | 119,947 | 1.12 (1.06 to 1.18) | 1.31e-04 |  |
| Social isolation | Depression trait | linear | unadjusted | 129,422 | 0.27 (0.20 to 0.33) | 1.06e-15 |  |
| Social isolation | Depression trait | linear | adjusted1 | 113,391 | 0.42 (0.35 to 0.49) | 1.19e-30 |  |
| Social isolation | Depression trait | linear | adjusted1 with disability | 111,508 | 0.38 (0.31 to 0.45) | 1.14e-25 |  |
| Social isolation | Depression trait | linear | adjusted1 with ACEs | 80,040 | 0.28 (0.20 to 0.36) | 5.67e-12 |  |
| Social isolation | Depression trait | linear | adjusted2 | 78,782 | 0.25 (0.17 to 0.33) | 3.85e-10 |  |
| Social isolation | Anxiety trait | linear | unadjusted | 129,997 | -0.07 (-0.13 to -0.01) | 0.02 |  |
| Social isolation | Anxiety trait | linear | adjusted1 | 113,874 | 0.10 (0.04 to 0.16) | 2.33e-03 |  |
| Social isolation | Anxiety trait | linear | adjusted1 with disability | 111,980 | 0.07 (6.83e-03 to 0.14) | 0.03 |  |
| Social isolation | Anxiety trait | linear | adjusted1 with ACEs | 80,355 | -0.05 (-0.12 to 0.03) | 0.21 |  |
| Social isolation | Anxiety trait | linear | adjusted2 | 79,081 | -0.06 (-0.14 to 6.86e-03) | 0.08 |  |
| Social isolation | Happy | linear | unadjusted | 226,040 | -0.21 (-0.22 to -0.20) | 0.00 |  |
| Social isolation | Happy | linear | adjusted1 | 191,383 | -0.22 (-0.23 to -0.21) | 0.00 |  |
| Social isolation | Happy | linear | adjusted1 with disability | 187,679 | -0.22 (-0.23 to -0.21) | 0.00 |  |
| Social isolation | Happy | linear | adjusted1 with ACEs | 95,039 | -0.16 (-0.18 to -0.15) | 4.64e-102 |  |
| Social isolation | Happy | linear | adjusted2 | 93,483 | -0.16 (-0.17 to -0.14) | 2.27e-97 |  |
| Social isolation | Meaning in life | linear | unadjusted | 128,746 | -0.33 (-0.35 to -0.32) | 0.00 |  |
| Social isolation | Meaning in life | linear | adjusted1 | 112,752 | -0.32 (-0.34 to -0.31) | 0.00 |  |
| Social isolation | Meaning in life | linear | adjusted1 with disability | 110,865 | -0.32 (-0.34 to -0.30) | 0.00 |  |
| Social isolation | Meaning in life | linear | adjusted1 with ACEs | 79,402 | -0.30 (-0.32 to -0.28) | 2.61e-225 |  |
| Social isolation | Meaning in life | linear | adjusted2 | 78,148 | -0.30 (-0.32 to -0.28) | 3.90e-220 |  |

*HR=hazard ratio, OR=odds ratio, CI=confidence interval, CoxPH=Cox proportional hazards, CAD=coronary artery disease, HF=heart failure, T2D=type 2 diabetes, SBP=systolic blood pressure, QALYs=quality-adjusted life years*

*Adjusted 1: age when attended assessment centre, sex, assessment centre, Townsend deprivation index, education years based on qualifications, household income before tax and ethnicity*

*Adjusted 2: adjusted 1 as well as long-standing illness/disability/infirmity and a score for adverse childhood experiences (ACEs)*

^a^ *For all Cox proportional hazards models the covariate ‘centre’ was not included due to issues with fitting the model*

*^b^ For all Cox proportional hazards models with death as an outcome the covariate ACEs was also not included due to issues with fitting the model*

### Table S7. Observational sensitivity analysis results

| **Exposure** | **Outcome** | **Model** | **Level of adjustment** | **N** | **Mean difference (95% CI)** | **p-value** |
| --- | --- | --- | --- | --- | --- | --- |
| Loneliness | Hospital admissions | Gamma | adjusted2 | 118,965 | 0.08 (0.06 to 0.10) | 1.90e-12 |
| Loneliness | Hospital admissions | Negative binomial | adjusted2 | 118,965 | 0.12 (0.09 to 0.16) | 1.09e-13 |
| Loneliness | Depression trait | Gamma | adjusted2 | 78,213 | 0.47 (0.46 to 0.49) | 0.00 |
| Loneliness | Depression trait | Negative binomial | adjusted2 | 78,213 | 0.63 (0.61 to 0.65) | 0.00 |
| Loneliness | Anxiety trait | Gamma | adjusted2 | 78,513 | 0.43 (0.41 to 0.45) | 0.00 |
| Loneliness | Anxiety trait | Negative binomial | adjusted2 | 78,513 | 0.62 (0.59 to 0.64) | 0.00 |
| Social isolation | Hospital admissions | Gamma | adjusted2 | 119,947 | -0.06 (-0.09 to -0.04) | 1.13e-06 |
| Social isolation | Hospital admissions | Negative binomial | adjusted2 | 119,947 | -0.09 (-0.12 to -0.05) | 8.77e-06 |
| Social isolation | Depression trait | Gamma | adjusted2 | 78,782 | 0.06 (0.04 to 0.08) | 1.40e-08 |
| Social isolation | Depression trait | Negative binomial | adjusted2 | 78,782 | 0.08 (0.05 to 0.11) | 1.86e-08 |
| Social isolation | Anxiety trait | Gamma | adjusted2 | 79,081 | -0.02 (-0.04 to 4.89e-05) | 0.05 |
| Social isolation | Anxiety trait | Negative binomial | adjusted2 | 79,081 | -0.03 (-0.07 to 7.64e-04) | 0.06 |

*OR=odds ratio, CI=confidence intervals*

*Adjusted 2: age when attended assessment centre, sex, assessment centre, Townsend deprivation index, education years based on qualifications, household income before tax and ethnicity, long-standing illness/disability/infirmity and a score for adverse childhood experiences (ACEs)*

### Table S8. Observational analyses for social isolation measure; sensitivity analysis excluding children

| **Exposure** | **Outcome** | **Model** | **Level of adjustment** | **N** | **HR, OR or mean difference (95% CI)** | **p-value** | **Proportional hazards test (CoxPH)** |
| --- | --- | --- | --- | --- | --- | --- | --- |
| Social isolation | CAD | CoxPH | unadjusted | 390,216 | 1.11 (1.06 to 1.16) | 9.74e-06 | 0.49 |
| Social isolation | CAD | CoxPH | adjusted1^a^ | 324,974 | 0.91 (0.86 to 0.96) | 2.31e-04 | 0.26 |
| Social isolation | CAD | CoxPH | adjusted2^a^ | 116,463 | 0.76 (0.68 to 0.85) | 1.87e-06 | 0.68 |
| Social isolation | HF | CoxPH | unadjusted | 410,050 | 1.46 (1.37 to 1.56) | 1.08e-28 | 0.40 |
| Social isolation | HF | CoxPH | adjusted1^a^ | 340,739 | 1.16 (1.08 to 1.25) | 1.06e-04 | 0.60 |
| Social isolation | HF | CoxPH | adjusted2^a^ | 119,621 | 1.01 (0.82 to 1.25) | 0.90 | 0.26 |
| Social isolation | Stroke | CoxPH | unadjusted | 404,985 | 1.24 (1.14 to 1.35) | 5.16e-07 | 0.27 |
| Social isolation | Stroke | CoxPH | adjusted1^a^ | 336,679 | 1.04 (0.95 to 1.15) | 0.36 | 0.37 |
| Social isolation | Stroke | CoxPH | adjusted2^a^ | 118,815 | 0.96 (0.74 to 1.23) | 0.74 | 0.64 |
| Social isolation | Death | CoxPH | unadjusted | 412,221 | 1.70 (1.63 to 1.77) | 4.71e-128 | 1.35e-03 |
| Social isolation | Death | CoxPH | adjusted1^a,b^ | 342,471 | 1.38 (1.31 to 1.44) | 6.88e-39 | 0.02 |
| Social isolation | Death | CoxPH | adjusted2^a,b^ | 335,351 | 1.39 (1.32 to 1.46) | 1.93e-40 | 0.08 |
| Social isolation | Multimorbidity | logistic | unadjusted | 412,122 | 1.04 (1.01 to 1.06) | 1.43e-03 |  |
| Social isolation | Multimorbidity | logistic | adjusted1 | 342,399 | 1.06 (1.03 to 1.09) | 4.47e-05 |  |
| Social isolation | Multimorbidity | logistic | adjusted2 | 119,877 | 1.04 (0.99 to 1.10) | 0.10 |  |
| Social isolation | T2D | logistic | unadjusted | 412,222 | 1.20 (1.14 to 1.27) | 6.73e-11 |  |
| Social isolation | T2D | logistic | adjusted1 | 342,472 | 1.08 (1.02 to 1.15) | 0.01 |  |
| Social isolation | T2D | logistic | adjusted2 | 119,895 | 1.20 (1.03 to 1.39) | 0.02 |  |
| Social isolation | Hospital admissions | linear | unadjusted | 412,222 | -0.34 (-0.49 to -0.19) | 7.43e-06 |  |
| Social isolation | Hospital admissions | linear | adjusted1 | 342,472 | -0.23 (-0.39 to -0.07) | 5.88e-03 |  |
| Social isolation | Hospital admissions | linear | adjusted2 | 119,895 | -0.41 (-0.56 to -0.27) | 2.18e-08 |  |
| Social isolation | SBP | linear | unadjusted | 411,404 | -2.34 (-2.56 to -2.12) | 2.41e-95 |  |
| Social isolation | SBP | linear | adjusted1 | 341,882 | -0.82 (-1.05 to -0.59) | 3.22e-12 |  |
| Social isolation | SBP | linear | adjusted2 | 119,802 | -0.64 (-1.04 to -0.25) | 1.40e-03 |  |
| Social isolation | QALYs | linear | unadjusted | 334,549 | 6.93e-04 (-2.04e-03 to 3.43e-03) | 0.62 |  |
| Social isolation | QALYs | linear | adjusted1 | 276,476 | 1.40e-03 (-1.45e-03 to 4.24e-03) | 0.34 |  |
| Social isolation | QALYs | linear | adjusted2 | 94,045 | 0.01 (0.01 to 0.02) | 4.27e-16 |  |
| Social isolation | Self-harm | logistic | unadjusted | 131,411 | 2.44 (2.20 to 2.71) | 4.85e-64 |  |
| Social isolation | Self-harm | logistic | adjusted1 | 114,932 | 1.80 (1.61 to 2.01) | 2.44e-24 |  |
| Social isolation | Self-harm | logistic | adjusted2 | 79,574 | 1.36 (1.19 to 1.57) | 1.28e-05 |  |
| Social isolation | Suicide attempt | logistic | unadjusted | 131,226 | 2.67 (2.32 to 3.07) | 3.51e-42 |  |
| Social isolation | Suicide attempt | logistic | adjusted1 | 114,770 | 1.84 (1.58 to 2.14) | 5.03e-15 |  |
| Social isolation | Suicide attempt | logistic | adjusted2 | 79,478 | 1.24 (1.03 to 1.50) | 0.03 |  |
| Social isolation | Depression diagnosis | logistic | unadjusted | 412,222 | 1.52 (1.47 to 1.57) | 1.98e-132 |  |
| Social isolation | Depression diagnosis | logistic | adjusted1 | 342,472 | 1.40 (1.34 to 1.45) | 4.02e-66 |  |
| Social isolation | Depression diagnosis | logistic | adjusted2 | 119,895 | 1.23 (1.15 to 1.31) | 5.42e-10 |  |
| Social isolation | Anxiety diagnosis | logistic | unadjusted | 412,222 | 1.23 (1.18 to 1.28) | 5.20e-23 |  |
| Social isolation | Anxiety diagnosis | logistic | adjusted1 | 342,472 | 1.24 (1.18 to 1.30) | 9.87e-20 |  |
| Social isolation | Anxiety diagnosis | logistic | adjusted2 | 119,895 | 1.07 (0.99 to 1.15) | 0.09 |  |
| Social isolation | Depression trait | linear | unadjusted | 129,379 | 0.84 (0.75 to 0.93) | 8.86e-74 |  |
| Social isolation | Depression trait | linear | adjusted1 | 113,351 | 0.55 (0.46 to 0.65) | 2.06e-30 |  |
| Social isolation | Depression trait | linear | adjusted2 | 78,759 | 0.33 (0.22 to 0.43) | 7.94e-10 |  |
| Social isolation | Anxiety trait | linear | unadjusted | 129,952 | 0.35 (0.27 to 0.43) | 3.97e-17 |  |
| Social isolation | Anxiety trait | linear | adjusted1 | 113,833 | 0.15 (0.07 to 0.24) | 4.25e-04 |  |
| Social isolation | Anxiety trait | linear | adjusted2 | 79,058 | -0.05 (-0.15 to 0.05) | 0.32 |  |
| Social isolation | Happy | linear | unadjusted | 225,839 | -0.42 (-0.43 to -0.41) | 0.00 |  |
| Social isolation | Happy | linear | adjusted1 | 191,255 | -0.37 (-0.38 to -0.35) | 0.00 |  |
| Social isolation | Happy | linear | adjusted2 | 93,449 | -0.31 (-0.33 to -0.29) | 1.88e-212 |  |
| Social isolation | Meaning in life | linear | unadjusted | 128,700 | -0.45 (-0.47 to -0.44) | 0.00 |  |
| Social isolation | Meaning in life | linear | adjusted1 | 112,711 | -0.40 (-0.42 to -0.38) | 1.07e-313 |  |
| Social isolation | Meaning in life | linear | adjusted2 | 78,125 | -0.37 (-0.40 to -0.35) | 1.58e-189 |  |

*HR=hazards ratio, OR=odds ratio, CI=confidence intervals, CoxPH=Cox proportional hazards, CAD=coronary artery disease, HF=heart failure, T2D=type 2 diabetes, SBP=systolic blood pressure, QALYs=quality-adjusted life years*

*Adjusted 1: age when attended assessment centre, sex, assessment centre, Townsend deprivation index, education years based on qualifications, household income before tax and ethnicity*

*Adjusted 2: adjusted 1 as well as long-standing illness/disability/infirmity and a score for adverse childhood experiences (ACEs)*

^a^ *For all Cox proportional hazards models the covariate ‘centre’ was not included due to issues with fitting the model*

*^b^ For all Cox proportional hazards models with death as an outcome the covariate ACEs was also not included due to issues with fitting the model*

### Table S9. Sibling control analysis results

| **Exposure** | **Outcome** | **Model** | **Level of adjustment** | **N** | **Within-family HR, OR or mean difference (95% CI)** | **Within-family p-value** | **Within-family proportional hazards test (CoxPH)** | **Between-family HR, OR or mean difference (95% CI)** | **Between-family p-value** | **Between-family proportional hazards test (CoxPH)** |
| --- | --- | --- | --- | --- | --- | --- | --- | --- | --- | --- |
| Loneliness | CAD | CoxPH | unadjusted | 37,899 | 1.11 (0.97 to 1.28) | 0.13 | 0.39 | 1.37 (1.20 to 1.56) | 4.55e-06 | 3.31e-03 |
| Loneliness | CAD | CoxPH | adjusted1^a^ | 31,845 | 1.14 (0.98 to 1.33) | 0.09 | 0.60 | 1.32 (1.13 to 1.53) | 3.70e-04 | 0.12 |
| Loneliness | CAD | CoxPH | adjusted2^a^ | 11,959 | 1.27 (0.92 to 1.75) | 0.14 | 0.47 | 1.30 (0.94 to 1.78) | 0.11 | 0.14 |
| Loneliness | HF | CoxPH | unadjusted | 39,736 | 1.33 (1.08 to 1.63) | 6.24e-03 | 0.36 | 1.86 (1.53 to 2.26) | 3.65e-10 | 0.07 |
| Loneliness | HF | CoxPH | adjusted1^a^ | 33,350 | 1.39 (1.10 to 1.75) | 5.56e-03 | 0.43 | 1.81 (1.46 to 2.26) | 1.09e-07 | 0.24 |
| Loneliness | HF | CoxPH | adjusted2^a^ | 32,667 | 1.26 (1.00 to 1.59) | 0.05 | 0.36 | 1.58 (1.26 to 1.97) | 6.18e-05 | 0.23 |
| Loneliness | Stroke | CoxPH | unadjusted | 39,286 | 1.22 (0.93 to 1.59) | 0.15 | 0.37 | 1.30 (1.01 to 1.68) | 0.05 | 0.04 |
| Loneliness | Stroke | CoxPH | adjusted1^a^ | 32,983 | 1.32 (0.98 to 1.78) | 0.07 | 0.11 | 1.41 (1.06 to 1.88) | 0.02 | 8.68e-03 |
| Loneliness | Stroke | CoxPH | adjusted2^a^ | 32,310 | 1.28 (0.94 to 1.73) | 0.12 | 0.10 | 1.27 (0.95 to 1.71) | 0.11 | 0.01 |
| Loneliness | Death | CoxPH | unadjusted | 39,950 | 1.34 (1.17 to 1.53) | 2.49e-05 | 0.02 | 1.81 (1.59 to 2.05) | 1.10e-19 | 1.02e-03 |
| Loneliness | Death | CoxPH | adjusted1^a,b^ | 33,515 | 1.28 (1.10 to 1.50) | 1.43e-03 | 4.54e-03 | 1.68 (1.45 to 1.95) | 2.94e-12 | 6.72e-03 |
| Loneliness | Death | CoxPH | adjusted2^a.b^ | 32,828 | 1.25 (1.07 to 1.46) | 5.93e-03 | 8.11e-03 | 1.53 (1.32 to 1.78) | 2.00e-08 | 4.87e-03 |
| Loneliness | Multimorbidity | logistic | Unadjusted | 39,942 | 1.93 (1.79 to 2.08) | 3.74e-62 |  | 2.77 (2.57 to 2.99) | 7.44e-155 |  |
| Loneliness | Multimorbidity | logistic | adjusted1 | 33,508 | 1.88 (1.72 to 2.04) | 1.29e-46 |  | 2.55 (2.34 to 2.77) | 5.69e-103 |  |
| Loneliness | Multimorbidity | logistic | adjusted2 | 12,319 | 1.73 (1.49 to 2.02) | 1.52e-12 |  | 2.05 (1.76 to 2.39) | 2.93e-20 |  |
| Loneliness | T2D | logistic | unadjusted | 39,950 | 1.17 (0.99 to 1.39) | 0.06 |  | 1.80 (1.53 to 2.11) | 6.82e-13 |  |
| Loneliness | T2D | logistic | adjusted1 | 33,515 | 1.27 (1.04 to 1.54) | 0.02 |  | 1.80 (1.49 to 2.17) | 9.73e-10 |  |
| Loneliness | T2D | logistic | adjusted2 | 12,322 | 0.97 (0.63 to 1.48) | 0.87 |  | 1.33 (0.87 to 2.04) | 0.18 |  |
| Loneliness | Hospital admissions | linear | unadjusted | 39,950 | 0.49 (0.14 to 0.84) | 6.12e-03 |  | 0.90 (0.53 to 1.27) | 1.85e-06 |  |
| Loneliness | Hospital admissions | linear | adjusted1 | 33,515 | 0.42 (0.07 to 0.77) | 0.02 |  | 0.91 (0.52 to 1.30) | 4.10e-06 |  |
| Loneliness | Hospital admissions | linear | adjusted2 | 12,322 | 0.15 (-0.27 to 0.57) | 0.50 |  | 0.52 (0.02 to 1.03) | 0.04 |  |
| Loneliness | SBP | linear | unadjusted | 39,922 | -2.41 (-3.10 to -1.73) | 3.65e-12 |  | -3.31 (-3.96 to -2.66) | 1.88e-23 |  |
| Loneliness | SBP | linear | adjusted1 | 33,492 | -1.37 (-2.07 to -0.68) | 1.13e-04 |  | -1.71 (-2.39 to -1.03) | 7.85e-07 |  |
| Loneliness | SBP | linear | adjusted2 | 12,317 | -1.43 (-2.59 to -0.27) | 0.02 |  | -1.95 (-3.11 to -0.80) | 9.40e-04 |  |
| Loneliness | QALYs | linear | unadjusted | 32,873 | -0.03 (-0.04 to -0.02) | 1.47e-13 |  | -0.07 (-0.07 to -0.06) | 2.69e-55 |  |
| Loneliness | QALYs | linear | adjusted1 | 27,455 | -0.02 (-0.03 to -0.01) | 3.92e-08 |  | -0.06 (-0.06 to -0.05) | 2.75e-38 |  |
| Loneliness | QALYs | linear | adjusted2 | 9,837 | -0.01 (-0.02 to -1.37e-03) | 0.03 |  | -0.03 (-0.04 to -0.02) | 4.12e-08 |  |
| Loneliness | Self-harm | logistic | unadjusted | 13,113 | 2.17 (1.61 to 2.92) | 3.06e-07 |  | 3.49 (2.62 to 4.65) | 1.66e-17 |  |
| Loneliness | Self-harm | logistic | adjusted1 | 11,558 | 1.92 (1.39 to 2.64) | 7.18e-05 |  | 2.75 (2.01 to 3.77) | 2.53e-10 |  |
| Loneliness | Self-harm | logistic | adjusted2 | 8,154 | 1.69 (1.13 to 2.54) | 0.01 |  | 1.56 (1.04 to 2.33) | 0.03 |  |
| Loneliness | Suicide attempt | logistic | unadjusted | 13,099 | 2.02 (1.36 to 3.02) | 5.44e-04 |  | 4.36 (2.97 to 6.41) | 6.44e-14 |  |
| Loneliness | Suicide attempt | logistic | adjusted1 | 11,547 | 1.75 (1.14 to 2.70) | 0.01 |  | 3.55 (2.34 to 5.38) | 2.83e-09 |  |
| Loneliness | Suicide attempt | logistic | adjusted2 | 8,146 | 1.85 (1.06 to 3.24) | 0.03 |  | 1.90 (1.10 to 3.31) | 0.02 |  |
| Loneliness | Depression diagnosis | logistic | unadjusted | 39,950 | 2.66 (2.41 to 2.92) | 9.97e-89 |  | 3.76 (3.43 to 4.12) | 1.41e-174 |  |
| Loneliness | Depression diagnosis | logistic | adjusted1 | 33,515 | 2.34 (2.11 to 2.61) | 4.30e-56 |  | 3.27 (2.95 to 3.63) | 1.62e-110 |  |
| Loneliness | Depression diagnosis | logistic | adjusted2 | 12,322 | 2.25 (1.89 to 2.67) | 8.17e-20 |  | 2.66 (2.23 to 3.17) | 4.64e-28 |  |
| Loneliness | Anxiety diagnosis | logistic | unadjusted | 39,950 | 1.89 (1.68 to 2.12) | 3.54e-27 |  | 2.45 (2.20 to 2.74) | 2.26e-57 |  |
| Loneliness | Anxiety diagnosis | logistic | adjusted1 | 33,515 | 1.77 (1.56 to 2.01) | 1.15e-18 |  | 2.17 (1.92 to 2.46) | 1.49e-34 |  |
| Loneliness | Anxiety diagnosis | logistic | adjusted2 | 12,322 | 1.69 (1.39 to 2.06) | 1.93e-07 |  | 2.01 (1.65 to 2.46) | 5.56e-12 |  |
| Loneliness | Depression trait | linear | unadjusted | 12,921 | 2.29 (2.00 to 2.58) | 9.24e-53 |  | 3.51 (3.22 to 3.80) | 9.03e-121 |  |
| Loneliness | Depression trait | linear | adjusted1 | 11,400 | 2.07 (1.76 to 2.38) | 1.92e-39 |  | 3.12 (2.81 to 3.42) | 1.91e-87 |  |
| Loneliness | Depression trait | linear | adjusted2 | 8,077 | 1.77 (1.41 to 2.12) | 1.39e-22 |  | 2.56 (2.22 to 2.90) | 1.20e-48 |  |
| Loneliness | Anxiety trait | linear | unadjusted | 12,985 | 1.59 (1.33 to 1.85) | 5.11e-33 |  | 2.70 (2.43 to 2.96) | 2.22e-86 |  |
| Loneliness | Anxiety trait | linear | adjusted1 | 11,454 | 1.36 (1.09 to 1.64) | 4.64e-22 |  | 2.44 (2.15 to 2.72) | 1.29e-63 |  |
| Loneliness | Anxiety trait | linear | adjusted2 | 8,100 | 1.27 (0.94 to 1.59) | 2.51e-14 |  | 1.95 (1.64 to 2.27) | 2.61e-33 |  |
| Loneliness | Happy | linear | unadjusted | 21,059 | -0.51 (-0.55 to -0.47) | 1.13e-156 |  | -0.68 (-0.72 to -0.65) | 2.51e-285 |  |
| Loneliness | Happy | linear | adjusted1 | 18,077 | -0.49 (-0.53 to -0.45) | 2.98e-124 |  | -0.63 (-0.67 to -0.59) | 2.46e-203 |  |
| Loneliness | Happy | linear | adjusted2 | 9,464 | -0.44 (-0.50 to -0.39) | 9.56e-52 |  | -0.56 (-0.62 to -0.51) | 5.58e-85 |  |
| Loneliness | Meaning in life | linear | unadjusted | 12,842 | -0.41 (-0.47 to -0.35) | 5.95e-43 |  | -0.58 (-0.64 to -0.53) | 2.78e-87 |  |
| Loneliness | Meaning in life | linear | adjusted1 | 11,341 | -0.39 (-0.45 to -0.33) | 6.13e-35 |  | -0.55 (-0.61 to -0.49) | 1.95e-67 |  |
| Loneliness | Meaning in life | linear | adjusted2 | 8,004 | -0.34 (-0.41 to -0.26) | 2.67e-19 |  | -0.49 (-0.56 to -0.42) | 4.59e-40 |  |
| Social isolation | CAD | CoxPH | unadjusted | 38,369 | 1.13 (0.94 to 1.36) | 0.21 | 0.69 | 1.15 (0.97 to 1.36) | 0.11 | 0.50 |
| Social isolation | CAD | CoxPH | adjusted1^a^ | 32,355 | 0.90 (0.73 to 1.10) | 0.29 | 0.95 | 0.84 (0.70 to 1.01) | 0.07 | 0.42 |
| Social isolation | CAD | CoxPH | adjusted2^a^ | 12,090 | 0.75 (0.50 to 1.12) | 0.16 | 0.97 | 0.85 (0.59 to 1.24) | 0.40 | 0.93 |
| Social isolation | HF | CoxPH | unadjusted | 40,232 | 1.29 (0.95 to 1.75) | 0.10 | 0.10 | 0.86 (0.65 to 1.13) | 0.28 | 0.07 |
| Social isolation | HF | CoxPH | adjusted1^a^ | 33,888 | 1.00 (0.72 to 1.39) | 1.00 | 0.04 | 0.69 (0.51 to 0.94) | 0.02 | 0.16 |
| Social isolation | HF | CoxPH | adjusted2^a^ | 33,163 | 1.08 (0.78 to 1.50) | 0.65 | 0.11 | 0.68 (0.50 to 0.92) | 0.01 | 0.16 |
| Social isolation | Stroke | CoxPH | unadjusted | 39,770 | 0.95 (0.66 to 1.38) | 0.79 | 0.31 | 0.77 (0.55 to 1.08) | 0.12 | 0.89 |
| Social isolation | Stroke | CoxPH | adjusted1^a^ | 33,510 | 0.85 (0.56 to 1.27) | 0.42 | 0.29 | 0.74 (0.51 to 1.07) | 0.11 | 0.94 |
| Social isolation | Stroke | CoxPH | adjusted2^a^ | 32,796 | 0.96 (0.64 to 1.45) | 0.85 | 0.25 | 0.74 (0.51 to 1.08) | 0.11 | 0.85 |
| Social isolation | Death | CoxPH | unadjusted | 40,440 | 1.71 (1.40 to 2.08) | 1.17e-07 | 0.13 | 1.07 (0.89 to 1.28) | 0.47 | 0.05 |
| Social isolation | Death | CoxPH | adjusted1^a,b^ | 34,053 | 1.57 (1.27 to 1.95) | 4.16e-05 | 0.02 | 0.96 (0.78 to 1.17) | 0.66 | 0.01 |
| Social isolation | Death | CoxPH | adjusted2^a,b^ | 33,325 | 1.67 (1.34 to 2.08) | 5.23e-06 | 0.04 | 0.94 (0.77 to 1.15) | 0.54 | 0.01 |
| Social isolation | Multimorbidity | logistic | unadjusted | 40,431 | 1.18 (1.08 to 1.30) | 3.95e-04 |  | 1.72 (1.59 to 1.86) | 2.42e-40 |  |
| Social isolation | Multimorbidity | logistic | adjusted1 | 34,046 | 0.99 (0.90 to 1.10) | 0.88 |  | 1.14 (1.04 to 1.26) | 4.99e-03 |  |
| Social isolation | Multimorbidity | logistic | adjusted2 | 12,451 | 1.05 (0.88 to 1.25) | 0.61 |  | 1.15 (0.98 to 1.36) | 0.09 |  |
| Social isolation | T2D | logistic | unadjusted | 40,440 | 1.62 (1.29 to 2.04) | 2.91e-05 |  | 2.16 (1.77 to 2.63) | 4.43e-14 |  |
| Social isolation | T2D | logistic | adjusted1 | 34,053 | 1.14 (0.88 to 1.48) | 0.33 |  | 1.00 (0.79 to 1.28) | 0.99 |  |
| Social isolation | T2D | logistic | adjusted2 | 12,454 | 0.94 (0.53 to 1.65) | 0.82 |  | 0.95 (0.57 to 1.59) | 0.85 |  |
| Social isolation | Hospital admissions | linear | unadjusted | 40,440 | 0.41 (-0.10 to 0.91) | 0.11 |  | 1.74 (1.34 to 2.14) | 7.86e-18 |  |
| Social isolation | Hospital admissions | linear | adjusted1 | 34,053 | -0.23 (-0.74 to 0.28) | 0.38 |  | -0.03 (-0.45 to 0.38) | 0.87 |  |
| Social isolation | Hospital admissions | linear | adjusted2 | 12,454 | -0.26 (-0.75 to 0.24) | 0.31 |  | -0.19 (-0.56 to 0.19) | 0.33 |  |
| Social isolation | SBP | linear | unadjusted | 40,410 | 1.29 (0.44 to 2.14) | 3.06e-03 |  | 7.48 (6.76 to 8.20) | 3.30e-92 |  |
| Social isolation | SBP | linear | adjusted1 | 34,028 | -0.25 (-1.12 to 0.62) | 0.58 |  | 0.23 (-0.55 to 1.00) | 0.57 |  |
| Social isolation | SBP | linear | adjusted2 | 12,449 | 0.69 (-0.65 to 2.03) | 0.31 |  | 1.02 (-0.21 to 2.24) | 0.10 |  |
| Social isolation | QALYs | linear | unadjusted | 33,251 | -0.04 (-0.05 to -0.03) | 5.97e-14 |  | -0.08 (-0.09 to -0.07) | 6.32e-80 |  |
| Social isolation | QALYs | linear | adjusted1 | 27,882 | -5.30e-03 (-0.02 to 4.69e-03) | 0.30 |  | -5.34e-03 (-0.01 to 3.76e-03) | 0.25 |  |
| Social isolation | QALYs | linear | adjusted2 | 9,950 | 2.10e-03 (-8.64e-03 to 0.01) | 0.70 |  | 0.01 (1.07e-03 to 0.02) | 0.03 |  |
| Social isolation | Self-harm | logistic | unadjusted | 13,226 | 2.45 (1.63 to 3.69) | 1.85e-05 |  | 1.02 (0.71 to 1.46) | 0.92 |  |
| Social isolation | Self-harm | logistic | adjusted1 | 11,702 | 1.77 (1.16 to 2.70) | 7.78e-03 |  | 1.71 (1.16 to 2.51) | 6.71e-03 |  |
| Social isolation | Self-harm | logistic | adjusted2 | 8,228 | 1.22 (0.73 to 2.02) | 0.45 |  | 1.37 (0.85 to 2.18) | 0.19 |  |
| Social isolation | Suicide attempt | logistic | unadjusted | 13,213 | 2.86 (1.61 to 5.06) | 3.20e-04 |  | 1.39 (0.84 to 2.30) | 0.20 |  |
| Social isolation | Suicide attempt | logistic | adjusted1 | 11,692 | 1.79 (0.99 to 3.23) | 0.05 |  | 1.83 (1.07 to 3.14) | 0.03 |  |
| Social isolation | Suicide attempt | logistic | adjusted2 | 8,221 | 1.43 (0.68 to 2.98) | 0.35 |  | 1.42 (0.72 to 2.80) | 0.31 |  |
| Social isolation | Depression diagnosis | logistic | unadjusted | 40,440 | 1.42 (1.24 to 1.61) | 2.01e-07 |  | 1.41 (1.26 to 1.58) | 3.09e-09 |  |
| Social isolation | Depression diagnosis | logistic | adjusted1 | 34,053 | 1.28 (1.11 to 1.48) | 8.96e-04 |  | 1.49 (1.31 to 1.70) | 3.35e-09 |  |
| Social isolation | Depression diagnosis | logistic | adjusted2 | 12,454 | 1.21 (0.97 to 1.52) | 0.10 |  | 1.36 (1.10 to 1.67) | 4.00e-03 |  |
| Social isolation | Anxiety diagnosis | logistic | unadjusted | 40,440 | 1.30 (1.11 to 1.52) | 1.06e-03 |  | 1.42 (1.24 to 1.62) | 6.14e-07 |  |
| Social isolation | Anxiety diagnosis | logistic | adjusted1 | 34,053 | 1.30 (1.10 to 1.55) | 2.61e-03 |  | 1.49 (1.27 to 1.74) | 9.41e-07 |  |
| Social isolation | Anxiety diagnosis | logistic | adjusted2 | 12,454 | 1.21 (0.94 to 1.57) | 0.15 |  | 1.26 (0.99 to 1.60) | 0.06 |  |
| Social isolation | Depression trait | linear | unadjusted | 13,027 | 0.44 (0.13 to 0.74) | 4.89e-03 |  | -0.07 (-0.34 to 0.21) | 0.63 |  |
| Social isolation | Depression trait | linear | adjusted1 | 11,540 | 0.24 (-0.08 to 0.55) | 0.14 |  | 0.36 (0.06 to 0.66) | 0.02 |  |
| Social isolation | Depression trait | linear | adjusted2 | 8,153 | 0.24 (-0.10 to 0.59) | 0.17 |  | 0.13 (-0.19 to 0.45) | 0.42 |  |
| Social isolation | Anxiety trait | linear | unadjusted | 13,092 | 0.04 (-0.23 to 0.32) | 0.76 |  | -0.46 (-0.70 to -0.21) | 2.17e-04 |  |
| Social isolation | Anxiety trait | linear | adjusted1 | 11,593 | -0.12 (-0.40 to 0.17) | 0.43 |  | -0.12 (-0.39 to 0.14) | 0.36 |  |
| Social isolation | Anxiety trait | linear | adjusted2 | 8,176 | 2.55e-03 (-0.31 to 0.31) | 0.99 |  | -0.29 (-0.59 to 8.94e-03) | 0.06 |  |
| Social isolation | Happy | linear | unadjusted | 21,282 | -0.20 (-0.24 to -0.15) | 1.31e-16 |  | -0.16 (-0.20 to -0.12) | 6.68e-14 |  |
| Social isolation | Happy | linear | adjusted1 | 18,324 | -0.16 (-0.21 to -0.11) | 1.37e-10 |  | -0.23 (-0.28 to -0.18) | 7.64e-23 |  |
| Social isolation | Happy | linear | adjusted2 | 9,561 | -0.14 (-0.21 to -0.08) | 2.39e-05 |  | -0.20 (-0.26 to -0.14) | 7.74e-11 |  |
| Social isolation | Meaning in life | linear | unadjusted | 12,953 | -0.32 (-0.39 to -0.25) | 1.25e-20 |  | -0.30 (-0.36 to -0.24) | 2.91e-24 |  |
| Social isolation | Meaning in life | linear | adjusted1 | 11,481 | -0.30 (-0.37 to -0.23) | 3.47e-16 |  | -0.35 (-0.42 to -0.29) | 2.18e-26 |  |
| Social isolation | Meaning in life | linear | adjusted2 | 8,075 | -0.30 (-0.38 to -0.22) | 1.40e-13 |  | -0.33 (-0.40 to -0.25) | 1.25e-17 |  |

*HR=hazards ratio, OR=odds ratio, CI=confidence intervals, CoxPH=Cox proportional hazards, CAD=coronary artery disease, HF=heart failure, T2D=type 2 diabetes, SBP=systolic blood pressure, QALYs=quality-adjusted life years*

*Adjusted 1: age when attended assessment centre, sex, assessment centre, Townsend deprivation index, education years based on qualifications, household income before tax and ethnicity*

*Adjusted 2: adjusted 1 as well as long-standing illness/disability/infirmity and a score for adverse childhood experiences (ACEs)*

*^a^ For all Cox proportional hazards models the covariate ‘centre’ was not included due to issues with fitting the model*

*^b^ For all Cox proportional hazards models with death as an outcome the covariate ACEs was also not included due to issues with fitting the model*

### Table S10. One-sample Mendelian Randomisation analysis results from split sample approach

| **Exposure** | **Outcome** | **N** | **HR, RD or mean difference (MD) (95% CI)** | **p-value** | **Meta-analysis heterogeneity test** | **F statistic** |
| --- | --- | --- | --- | --- | --- | --- |
| Loneliness | CAD | 314,205 | HR: 3.65 (0.40 to 33.72) | 0.25 | 1.33; p=0.25 | Split sample 1: 26.09; Split sample 2: 54.68 |
| Loneliness | HF | 330,211 | HR: 18.51 (0.67 to 509.34) | 0.08 | 0.74; p=0.39 | Split sample 1: 26.40; Split sample 2: 53.30 |
| Loneliness | Stroke | 326,048 | HR: 0.18 (3.28e-03 to 9.87) | 0.40 | 1.75e-03; p=0.97 | Split sample 1: 25.67; Split sample 2: 54.96 |
| Loneliness | Death | 331,995 | HR: 1.26 (0.16 to 9.79) | 0.83 | 0.05; p=0.83 | Split sample 1: 27.57; Split sample 2: 55.98 |
| Loneliness | Multimorbidity | 331,918 | RD: 0.71 (0.41 to 1.01) | 3.94e-06 | 2.37; p=0.12 | Split sample 1: 28.19; Split sample 2: 55.13 |
| Loneliness | T2D | 331,996 | RD: 0.22 (0.10 to 0.35) | 3.60e-04 | 2.34; p=0.13 | Split sample 1: 28.07; Split sample 2: 55.18 |
| Loneliness | Hospital admissions | 331,996 | MD: 4.17 (-2.98 to 11.33) | 0.25 | 0.97; p=0.33 | Split sample 1: 28.07; Split sample 2: 55.18 |
| Loneliness | SBP | 331,696 | MD: -0.59 (-10.51 to 9.33) | 0.91 | 0.07; p=0.78 | Split sample 1: 27.99; Split sample 2: 54.08 |
| Loneliness | QALYs | 269,905 | MD: -0.25 (-0.38 to -0.13) | 1.00e-04 | 0.46; p=0.50 | Split sample 1: 21.68; Split sample 2: 42.47 |
| Loneliness | Self-harm | 108,966 | RD: 0.31 (0.08 to 0.54) | 7.51e-03 | 0.06; p=0.81 | Split sample 1: 14.65; Split sample 2: 13.13 |
| Loneliness | Suicide attempt | 108,827 | RD: 0.15 (-7.43e-03 to 0.31) | 0.06 | 1.14; p=0.29 | Split sample 1: 14.55; Split sample 2: 13.46 |
| Loneliness | Depression diagnosis | 331,996 | RD: 0.35 (0.15 to 0.55) | 5.18e-04 | 0.62; p=0.43 | Split sample 1: 28.07; Split sample 2: 55.18 |
| Loneliness | Anxiety diagnosis | 331,996 | RD: 0.13 (-0.04 to 0.29) | 0.13 | 4.77e-03; p=0.94 | Split sample 1: 28.07; Split sample 2: 55.18 |
| Loneliness | Depression trait | 107,357 | MD: 13.00 (7.62 to 18.37) | 2.16e-06 | 6.43e-04; p=0.98 | Split sample 1: 12.77; Split sample 2: 13.66 |
| Loneliness | Anxiety trait | 107,853 | MD: 7.81 (3.88 to 11.74) | 9.81e-05 | 0.11; p=0.73 | Split sample 1: 16.07; Split sample 2: 12.49 |
| Loneliness | Happy | 178,666 | MD: -1.76 (-2.40 to -1.12) | 7.98e-08 | 0.27; p=0.61 | Split sample 1: 20.04; Split sample 2: 25.30 |
| Loneliness | Meaning in life | 106,828 | MD: -0.92 (-1.74 to -0.10) | 0.03 | 5.14e-03; p=0.94 | Split sample 1: 14.00; Split sample 2: 15.50 |
| Social isolation | CAD | 315,599 | HR: 7.43 (0.20 to 277.95) | 0.28 | 2.12; p=0.15 | Split sample 1: 32.21; Split sample 2: 9.98 |
| Social isolation | HF | 331,588 | HR: 0.03 (1.55e-04 to 5.49) | 0.19 | 0.13; p=0.72 | Split sample 1: 33.62; Split sample 2: 9.54 |
| Social isolation | Stroke | 321,451 | HR: 0.53 (9.13e-04 to 302.48) | 0.84 | 0.03; p=0.85 | Split sample 1: 32.20; Split sample 2: 10.66 |
| Social isolation | Death | 333,357 | HR: 9.90 (0.31 to 314.75) | 0.19 | 1.49; p=0.22 | Split sample 1: 33.38; Split sample 2: 9.46 |
| Social isolation | Multimorbidity | 333,280 | RD: 0.02 (-0.41 to 0.45) | 0.92 | 0.11; p=0.74 | Split sample 1: 38.74; Split sample 2: 11.30 |
| Social isolation | T2D | 333,358 | RD: 0.05 (-0.13 to 0.23) | 0.58 | 1.07; p=0.30 | Split sample 1: 39.26; Split sample 2: 11.20 |
| Social isolation | Hospital admissions | 333,358 | MD: -0.25 (-12.18 to 11.67) | 0.97 | 3.66; p=0.06 | Split sample 1: 39.26; Split sample 2: 11.20 |
| Social isolation | SBP | 333,057 | MD: 11.11 (-4.40 to 26.62) | 0.16 | 0.32; p=0.57 | Split sample 1: 39.13; Split sample 2: 11.17 |
| Social isolation | QALYs | 270,935 | MD: -0.05 (-0.24 to 0.14) | 0.62 | 1.11; p=0.29 | Split sample 1: 28.16; Split sample 2: 8.69 |
| Social isolation | Self-harm | 109,432 | RD: 0.14 (-0.08 to 0.37) | 0.20 | 0.16; p=0.69 | Split sample 1: 23.20; Split sample 2: 9.02 |
| Social isolation | Suicide attempt | 109,291 | RD: 0.07 (-0.09 to 0.23) | 0.39 | 1.58; p=0.21 | Split sample 1: 23.53; Split sample 2: 9.21 |
| Social isolation | Depression diagnosis | 333,358 | RD: 0.06 (-0.24 to 0.37) | 0.69 | 0.86; p=0.35 | Split sample 1: 39.26; Split sample 2: 11.20 |
| Social isolation | Anxiety diagnosis | 333,358 | RD: -0.12 (-0.38 to 0.13) | 0.35 | 2.69e-03; p=0.96 | Split sample 1: 39.26; Split sample 2: 11.20 |
| Social isolation | Depression trait | 107,790 | MD: 0.12 (-3.75 to 4.00) | 0.95 | 0.14; p=0.71 | Split sample 1: 22.04; Split sample 2: 10.56 |
| Social isolation | Anxiety trait | 108,274 | MD: -1.29 (-4.95 to 2.37) | 0.49 | 0.27; p=0.60 | Split sample 1: 22.74; Split sample 2: 9.46 |
| Social isolation | Happy | 179,367 | MD: -1.11 (-1.88 to -0.35) | 4.42e-03 | 0.05; p=0.83 | Split sample 1: 27.53; Split sample 2: 10.60 |
| Social isolation | Meaning in life | 107,258 | MD: -1.42 (-2.42 to -0.42) | 5.35e-03 | 0.13; p=0.72 | Split sample 1: 19.29; Split sample 2: 10.36 |

*HR=hazards ratio, RD=risk difference, CI=confidence intervals, CAD=coronary artery disease, HF=heart failure, T2D=type 2 diabetes, SBP=systolic blood pressure, QALYs=quality-adjusted life years*

*Adjusted for age, sex, the first 20 principal components (PCs) from PC analysis of genotype data, assessment centre and genotyping chip*

### Table S11. One-sample Mendelian Randomisation analysis results from unweighted loneliness approach

| **Exposure** | **Outcome** | **N** | **HR, RD or mean difference (95% CI)** | **p-value** | **F statistic** |
| --- | --- | --- | --- | --- | --- |
| Loneliness | CAD | 314,205 | 1.25 (0.44 to 3.51) | 0.68 | 360.30 |
| Loneliness | HF | 330,211 | 1.81 (0.41 to 7.92) | 0.43 | 385.74 |
| Loneliness | Stroke | 326,048 | 2.40 (0.38 to 15.24) | 0.35 | 377.52 |
| Loneliness | Death | 331,995 | 1.04 (0.40 to 2.71) | 0.93 | 387.44 |
| Loneliness | Multimorbidity | 331,918 | 0.44 (0.31 to 0.57) | 2.29e-11 | 388.26 |
| Loneliness | T2D | 331,996 | 0.05 (-3.68e-04 to 0.10) | 0.05 | 389.09 |
| Loneliness | Hospital admissions | 331,996 | 1.00 (-2.33 to 4.33) | 0.56 | 389.09 |
| Loneliness | SBP | 331,696 | -10.76 (-15.42 to -6.11) | 5.82e-06 | 387.04 |
| Loneliness | QALYs | 269,905 | -0.14 (-0.20 to -0.09) | 2.47e-07 | 310.99 |
| Loneliness | Self-harm | 108,966 | 0.23 (0.13 to 0.32) | 2.27e-06 | 147.69 |
| Loneliness | Suicide attempt | 108,827 | 0.16 (0.09 to 0.22) | 8.29e-06 | 147.61 |
| Loneliness | Depression diagnosis | 331,996 | 0.34 (0.25 to 0.43) | 3.73e-13 | 389.09 |
| Loneliness | Anxiety diagnosis | 331,996 | 0.19 (0.11 to 0.26) | 1.55e-06 | 389.09 |
| Loneliness | Depression trait | 107,357 | 7.06 (5.29 to 8.83) | 5.72e-15 | 137.24 |
| Loneliness | Anxiety trait | 107,853 | 4.55 (3.04 to 6.06) | 3.30e-09 | 147.86 |
| Loneliness | Happy | 178,666 | -1.15 (-1.40 to -0.90) | 1.15e-19 | 235.58 |
| Loneliness | Meaning in life | 106,828 | -0.97 (-1.34 to -0.60) | 2.35e-07 | 146.14 |

*HR=hazards ratio, RD=risk difference, CI=confidence intervals, CAD=coronary artery disease, HF=heart failure, T2D=type 2 diabetes, SBP=systolic blood pressure, QALYs=quality-adjusted life years*

*Adjusted for age, sex, the first 20 principal components (PCs) from PC analysis of genotype data, assessment centre and genotyping chip*

### Table S12. One-sample Mendelian Randomisation analysis sensitivity analyses

| **Exposure** | **Outcome** | **Method** | **Mean difference (95% CI)** | **p-value** |
| --- | --- | --- | --- | --- |
| Loneliness | CAD | Weighted median | 0.08 (-0.01 to 0.18) | 0.09 |
| Loneliness | CAD | Weighted mode | 0.07 (-0.15 to 0.29) | 0.52 |
| Loneliness | HF | Weighted median | 0.05 (-0.09 to 0.19) | 0.50 |
| Loneliness | HF | Weighted mode | -0.09 (-0.43 to 0.25) | 0.60 |
| Loneliness | Stroke | Weighted median | -0.06 (-0.23 to 0.12) | 0.53 |
| Loneliness | Stroke | Weighted mode | -0.11 (-0.52 to 0.29) | 0.58 |
| Loneliness | Death | Weighted median | 0.02 (-0.07 to 0.11) | 0.65 |
| Loneliness | Death | Weighted mode | 0.06 (-0.14 to 0.27) | 0.56 |
| Loneliness | Multimorbidity | Weighted median | 0.06 (7.66e-03 to 0.11) | 0.03 |
| Loneliness | Multimorbidity | Weighted mode | 0.05 (-0.07 to 0.18) | 0.41 |
| Loneliness | T2D | Weighted median | 0.22 (0.10 to 0.34) | 3.73e-04 |
| Loneliness | T2D | Weighted mode | 0.26 (-0.03 to 0.55) | 0.07 |
| Loneliness | Hospital admissions | Weighted median | 0.09 (-0.22 to 0.40) | 0.56 |
| Loneliness | Hospital admissions | Weighted mode | -0.03 (-0.78 to 0.72) | 0.94 |
| Loneliness | SBP | Weighted median | 0.01 (-0.43 to 0.45) | 0.96 |
| Loneliness | SBP | Weighted mode | -0.26 (-1.23 to 0.71) | 0.60 |
| Loneliness | QALYs | Weighted median | -5.78e-03 (-0.01 to -4.10e-04) | 0.03 |
| Loneliness | QALYs | Weighted mode | -1.35e-03 (-0.01 to 0.01) | 0.84 |
| Loneliness | Self-harm | Weighted median | 0.26 (0.04 to 0.48) | 0.02 |
| Loneliness | Self-harm | Weighted mode | 0.31 (-0.27 to 0.88) | 0.30 |
| Loneliness | Suicide attempt | Weighted median | 0.32 (0.02 to 0.62) | 0.04 |
| Loneliness | Suicide attempt | Weighted mode | 0.54 (-0.23 to 1.32) | 0.17 |
| Loneliness | Depression diagnosis | Weighted median | 0.07 (8.17e-04 to 0.14) | 0.05 |
| Loneliness | Depression diagnosis | Weighted mode | 0.04 (-0.13 to 0.22) | 0.65 |
| Loneliness | Anxiety diagnosis | Weighted median | 0.08 (-6.69e-03 to 0.16) | 0.07 |
| Loneliness | Anxiety diagnosis | Weighted mode | 0.03 (-0.18 to 0.23) | 0.81 |
| Loneliness | Depression trait | Weighted median | 0.34 (0.19 to 0.50) | 2.09e-05 |
| Loneliness | Depression trait | Weighted mode | 0.55 (0.16 to 0.93) | 5.33e-03 |
| Loneliness | Anxiety trait | Weighted median | 0.25 (0.11 to 0.39) | 3.95e-04 |
| Loneliness | Anxiety trait | Weighted mode | 0.25 (-0.07 to 0.58) | 0.13 |
| Loneliness | Happy | Weighted median | -0.04 (-0.07 to -0.02) | 3.23e-04 |
| Loneliness | Happy | Weighted mode | -0.04 (-0.11 to 0.02) | 0.16 |
| Loneliness | Meaning in life | Weighted median | -0.02 (-0.06 to 0.02) | 0.34 |
| Loneliness | Meaning in life | Weighted mode | 1.85e-03 (-0.09 to 0.09) | 0.97 |
| Social isolation | CAD | Weighted median | 0.79 (-0.01 to 1.60) | 0.05 |
| Social isolation | CAD | Weighted mode | 1.38 (-0.59 to 3.35) | 0.17 |
| Social isolation | HF | Weighted median | -0.02 (-1.14 to 1.11) | 0.98 |
| Social isolation | HF | Weighted mode | 0.09 (-2.39 to 2.58) | 0.94 |
| Social isolation | Stroke | Weighted median | 0.05 (-1.33 to 1.43) | 0.94 |
| Social isolation | Stroke | Weighted mode | 0.29 (-3.02 to 3.61) | 0.86 |
| Social isolation | Death | Weighted median | 0.23 (-0.50 to 0.97) | 0.53 |
| Social isolation | Death | Weighted mode | -0.73 (-2.44 to 0.98) | 0.40 |
| Social isolation | Multimorbidity | Weighted median | 0.04 (-0.38 to 0.46) | 0.85 |
| Social isolation | Multimorbidity | Weighted mode | -0.04 (-1.01 to 0.92) | 0.93 |
| Social isolation | T2D | Weighted median | -0.42 (-1.44 to 0.60) | 0.42 |
| Social isolation | T2D | Weighted mode | -1.44 (-3.66 to 0.78) | 0.20 |
| Social isolation | Hospital admissions | Weighted median | 1.08 (-1.44 to 3.59) | 0.40 |
| Social isolation | Hospital admissions | Weighted mode | 0.09 (-6.11 to 6.30) | 0.98 |
| Social isolation | SBP | Weighted median | 0.56 (-3.30 to 4.41) | 0.78 |
| Social isolation | SBP | Weighted mode | 2.13 (-6.05 to 10.30) | 0.61 |
| Social isolation | QALYs | Weighted median | -0.01 (-0.06 to 0.03) | 0.60 |
| Social isolation | QALYs | Weighted mode | -0.01 (-0.12 to 0.09) | 0.78 |
| Social isolation | Self-harm | Weighted median | 0.28 (-1.52 to 2.08) | 0.76 |
| Social isolation | Self-harm | Weighted mode | 0.02 (-4.04 to 4.08) | 0.99 |
| Social isolation | Suicide attempt | Weighted median | 0.90 (-1.66 to 3.47) | 0.49 |
| Social isolation | Suicide attempt | Weighted mode | 4.15 (-1.84 to 10.15) | 0.17 |
| Social isolation | Depression diagnosis | Weighted median | 0.04 (-0.55 to 0.63) | 0.90 |
| Social isolation | Depression diagnosis | Weighted mode | -0.22 (-1.74 to 1.31) | 0.78 |
| Social isolation | Anxiety diagnosis | Weighted median | -0.11 (-0.83 to 0.61) | 0.76 |
| Social isolation | Anxiety diagnosis | Weighted mode | 0.19 (-1.35 to 1.72) | 0.81 |
| Social isolation | Depression trait | Weighted median | -0.13 (-1.48 to 1.22) | 0.85 |
| Social isolation | Depression trait | Weighted mode | -0.99 (-4.19 to 2.21) | 0.55 |
| Social isolation | Anxiety trait | Weighted median | -0.68 (-1.85 to 0.50) | 0.26 |
| Social isolation | Anxiety trait | Weighted mode | -0.86 (-3.68 to 1.96) | 0.55 |
| Social isolation | Happy | Weighted median | -0.24 (-0.45 to -0.03) | 0.02 |
| Social isolation | Happy | Weighted mode | -0.37 (-0.89 to 0.15) | 0.16 |
| Social isolation | Meaning in life | Weighted median | -0.15 (-0.46 to 0.15) | 0.31 |
| Social isolation | Meaning in life | Weighted mode | 0.23 (-0.45 to 0.91) | 0.51 |

*CI=confidence intervals, CAD=coronary artery disease, HF=heart failure, T2D=type 2 diabetes, SBP=systolic blood pressure, QALYs=quality-adjusted life years*

*Effect estimates are presented as the mean difference due to the approach used to generate split sample results and subsequent meta-analyses.*

### Table S13. I-squared values for Two-sample Mendelian Randomisation analyses

| **Exposure** | **Outcome** | **Unweighted I squared** | **Weighted I squared** | **Mean F statistic** |
| --- | --- | --- | --- | --- |
| Loneliness | CAD | 0.30 | 0.00 | 35.55 |
| Loneliness | Systolic blood pressure | 0.30 | 0.00 | 35.55 |
| Loneliness | Heart failure | 0.30 | 0.43 | 35.55 |
| Loneliness | Stroke | 0.30 | 0.00 | 35.55 |
| Loneliness | T2D | 0.30 | 0.00 | 35.55 |
| Loneliness | Suicide attempt | 0.38 | 0.00 | 36.31 |
| Loneliness | Depression | 0.30 | 0.00 | 35.55 |
| Loneliness | Anxiety | 0.30 | 0.83 | 35.55 |
| Loneliness | Wellbeing spectrum | 0.00 | 0.00 | 34.94 |
| Loneliness | Positive affect | 0.00 | 0.00 | 34.94 |
| Loneliness | Life satisfaction | 0.00 | 0.00 | 34.94 |
| Social isolation | CAD | 0.48 | 0.00 | 38.43 |
| Social isolation | Systolic blood pressure | 0.51 | 0.00 | 38.66 |
| Social isolation | Heart failure | 0.48 | 0.54 | 38.43 |
| Social isolation | Stroke | 0.48 | 0.24 | 38.43 |
| Social isolation | T2D | 0.48 | 0.00 | 38.43 |
| Social isolation | Suicide attempt | 0.55 | 0.00 | 40.07 |
| Social isolation | Depression | 0.48 | 0.00 | 38.43 |
| Social isolation | Anxiety | 0.48 | 0.68 | 38.43 |
| Social isolation | Wellbeing spectrum | 0.52 | 0.00 | 38.51 |
| Social isolation | Positive affect | 0.52 | 0.00 | 38.51 |
| Social isolation | Life satisfaction | 0.52 | 0.00 | 38.51 |
| CAD | Loneliness | 0.95 | 0.94 | 84.91 |
| Systolic blood pressure | Loneliness | 0.66 | 0.73 | 44.63 |
| Heart failure | Loneliness | 0.73 | 0.18 | 42.34 |
| Stroke | Loneliness | 0.64 | 0.58 | 23.16 |
| T2D | Loneliness | 0.96 | 0.97 | 103.11 |
| Suicide attempt | Loneliness | 0.97 | 1.00 | 7573.13 |
| Depression | Loneliness | 0.20 | 0.08 | 11.83 |
| Anxiety | Loneliness | 0.38 | 0.81 | 21.75 |
| Wellbeing spectrum | Loneliness | 0.39 | 0.02 | 41.00 |
| Positive affect | Loneliness | 0.16 | 0.36 | 13.20 |
| Life satisfaction | Loneliness | 0.00 | 0.00 | 3.49 |
| CAD | Social isolation | 0.95 | 0.94 | 85.02 |
| Systolic blood pressure | Social isolation | 0.65 | 0.17 | 44.51 |
| Heart failure | Social isolation | 0.73 | 0.11 | 42.34 |
| Stroke | Social isolation | 0.64 | 0.59 | 23.16 |
| T2D | Social isolation | 0.96 | 0.96 | 102.51 |
| Suicide attempt | Social isolation | 0.97 | 1.00 | 7573.13 |
| Depression | Social isolation | 0.20 | 0.08 | 11.83 |
| Anxiety | Social isolation | 0.55 | 0.00 | 21.84 |
| Wellbeing spectrum | Social isolation | 0.39 | 0.00 | 41.00 |
| Positive affect | Social isolation | 0.15 | 0.06 | 13.28 |
| Life satisfaction | Social isolation | 0.00 | 0.00 | 3.49 |

*CAD=coronary artery disease, HF=heart failure, T2D=type 2 diabetes, SBP=systolic blood pressure, QALYs=quality-adjusted life years*

### Table S14. Two-sample Mendelian Randomisation analysis results with loneliness and social isolation as exposures

| **Exposure** | **Outcome** | **Method** | **N genetic variants** | **OR or mean difference (95% CI)** | **p-value** | **Heterogeneity test p-value** | **Directional pleiotropy intercept (95% CI; p-value)** | **Additional tests for MR approaches** | **Steiger filtering - percentage of genetic variants** |
| --- | --- | --- | --- | --- | --- | --- | --- | --- | --- |
| Loneliness | CAD | Inverse variance weighted | 16 | 1.15 (0.92 to 1.44) | 0.22 | 4.68e-04 |  |  | 100.00 |
| Loneliness | CAD | MR-Egger | 16 | 2.14 (0.67 to 6.86) | 0.22 | 7.58e-04 | -0.01 (-0.04 to 0.01, 0.31) |  |  |
| Loneliness | CAD | SIMEX adjusted MR-Egger | 16 | 1.75 (0.58 to 5.23) | 0.33 |  | -8.47e-03 (-0.03 to 0.02, 0.51) |  |  |
| Loneliness | CAD | Weighted median | 16 | 1.19 (0.95 to 1.49) | 0.14 |  |  |  |  |
| Loneliness | CAD | MR-RAPS | 16 | 1.21 (0.96 to 1.51) | 0.10 |  |  |  |  |
| Loneliness | CAD | PRESSO - raw | 16 | 1.15 (0.92 to 1.44) | 0.24 |  |  | Global = 45.23; p=1.50e-03, Distortion = NA; p=NA |  |
| Loneliness | CAD | PRESSO - outlier corrected | 15 (1 removed) | NA (NA to NA) |  |  |  |  |  |
| Loneliness | CAD | MRLAP - observed | 14 | 1.04 (0.94 to 1.15) | 0.43 |  |  |  |  |
| Loneliness | CAD | MRLAP - corrected | 14 | 1.05 (0.90 to 1.23) | 0.52 |  |  | Corrected vs observed estimate difference test = -0.41; p=0.68 |  |
| Loneliness | CAD | MR-CAUSE^a^ |  | 1.23 (1.07 to 1.42) |  |  |  | Causal vs sharing model ELPD: delta (SE) = -3.12 (1.90); p=0.05 |  |
| Loneliness | Systolic blood pressure | Inverse variance weighted | 16 | -2.63 (-5.90 to 0.63) | 0.11 | 2.00e-40 |  |  | 87.50 |
| Loneliness | Systolic blood pressure | MR-Egger | 16 | -6.72 (-24.10 to 10.67) | 0.46 | 2.60e-40 | 0.09 (-0.27 to 0.44, 0.65) |  |  |
| Loneliness | Systolic blood pressure | SIMEX adjusted MR-Egger | 16 | -2.66 (-19.64 to 14.33) | 0.76 |  | -7.27e-04 (-0.38 to 0.38, 1.00) |  |  |
| Loneliness | Systolic blood pressure | Weighted median | 16 | -2.07 (-3.80 to -0.34) | 0.02 |  |  |  |  |
| Loneliness | Systolic blood pressure | MR-RAPS | 16 | -3.03 (-6.26 to 0.19) | 0.07 |  |  |  |  |
| Loneliness | Systolic blood pressure | PRESSO - raw | 16 | -2.63 (-5.90 to 0.63) | 0.13 |  |  | Global = 259.59; p=<5e-04, Distortion = -45.25; p=<5e-04 |  |
| Loneliness | Systolic blood pressure | PRESSO - outlier corrected | 9 (7 removed) | -1.81 (-3.62 to -6.28e-03) | 0.08 |  |  |  |  |
| Loneliness | Systolic blood pressure | MRLAP - observed | 8 | -0.27 (-0.71 to 0.16) | 0.22 |  |  |  |  |
| Loneliness | Systolic blood pressure | MRLAP - corrected | 8 | -0.44 (-1.17 to 0.28) | 0.23 |  |  | Corrected vs observed estimate difference test = 1.18; p=0.24 |  |
| Loneliness | Systolic blood pressure | MR-CAUSE^a^ |  | -0.46 (-1.78 to 0.86) |  |  |  | Causal vs sharing model ELPD: delta (SE) = 0.49 (0.48); p=0.85 |  |
| Loneliness | Heart failure | Inverse variance weighted | 16 | 0.95 (0.73 to 1.24) | 0.72 | 0.13 |  |  | 100.00 |
| Loneliness | Heart failure | MR-Egger | 16 | 1.51 (0.37 to 6.12) | 0.57 | 0.11 | -9.67e-03 (-0.04 to 0.02, 0.52) |  |  |
| Loneliness | Heart failure | SIMEX adjusted MR-Egger | 16 | 1.95 (0.26 to 14.44) | 0.52 |  | -0.01 (-0.06 to 0.03, 0.49) |  |  |
| Loneliness | Heart failure | Weighted median | 16 | 1.02 (0.75 to 1.39) | 0.89 |  |  |  |  |
| Loneliness | Heart failure | MR-RAPS | 16 | 1.01 (0.79 to 1.29) | 0.95 |  |  |  |  |
| Loneliness | Heart failure | PRESSO - raw | 16 | 0.95 (0.73 to 1.24) | 0.72 |  |  | Global = 24.05; p=0.14, Distortion = NA; p=NA |  |
| Loneliness | Heart failure | PRESSO - outlier corrected | 16 (0 removed) | NA (NA to NA) |  |  |  |  |  |
| Loneliness | Heart failure | MRLAP - observed | 15 | 0.98 (0.90 to 1.07) | 0.70 |  |  |  |  |
| Loneliness | Heart failure | MRLAP - corrected | 15 | 0.97 (0.85 to 1.10) | 0.61 |  |  | Corrected vs observed estimate difference test = 0.74; p=0.46 |  |
| Loneliness | Heart failure | MR-CAUSE^a^ |  | 1.12 (0.96 to 1.30) |  |  |  | Causal vs sharing model ELPD: delta (SE) = -0.40 (1.25); p=0.38 |  |
| Loneliness | Stroke | Inverse variance weighted | 16 | 0.90 (0.61 to 1.31) | 0.58 | 9.25e-03 |  |  | 50.00 |
| Loneliness | Stroke | MR-Egger | 16 | 1.54 (0.20 to 12.10) | 0.69 | 7.06e-03 | -0.01 (-0.05 to 0.03, 0.61) |  |  |
| Loneliness | Stroke | SIMEX adjusted MR-Egger | 16 | 0.77 (0.11 to 5.19) | 0.79 |  | 3.66e-03 (-0.04 to 0.05, 0.87) |  |  |
| Loneliness | Stroke | Weighted median | 16 | 1.09 (0.73 to 1.61) | 0.68 |  |  |  |  |
| Loneliness | Stroke | MR-RAPS | 16 | 0.95 (0.66 to 1.36) | 0.78 |  |  |  |  |
| Loneliness | Stroke | PRESSO - raw | 16 | 0.90 (0.61 to 1.31) | 0.58 |  |  | Global = 35.03; p=0.01, Distortion = -1049.68; p=0.04 |  |
| Loneliness | Stroke | PRESSO - outlier corrected | 15 (1 removed) | 1.01 (0.74 to 1.38) | 0.94 |  |  |  |  |
| Loneliness | Stroke | MRLAP - observed | 8 | 0.92 (0.83 to 1.04) | 0.18 |  |  |  |  |
| Loneliness | Stroke | MRLAP - corrected | 8 | 0.88 (0.73 to 1.06) | 0.17 |  |  | Corrected vs observed estimate difference test = 1.39; p=0.16 |  |
| Loneliness | Stroke | MR-CAUSE^a^ |  | 1.09 (0.88 to 1.38) |  |  |  | Causal vs sharing model ELPD: delta (SE) = 0.62 (0.68); p=0.82 |  |
| Loneliness | T2D | Inverse variance weighted | 16 | 1.19 (0.85 to 1.67) | 0.31 | 2.60e-14 |  |  | 100.00 |
| Loneliness | T2D | MR-Egger | 16 | 2.83 (0.47 to 17.18) | 0.28 | 1.38e-13 | -0.02 (-0.06 to 0.02, 0.35) |  |  |
| Loneliness | T2D | SIMEX adjusted MR-Egger | 16 | 2.68 (0.43 to 16.78) | 0.31 |  | -0.02 (-0.06 to 0.02, 0.45) |  |  |
| Loneliness | T2D | Weighted median | 16 | 1.00 (0.80 to 1.25) | 0.98 |  |  |  |  |
| Loneliness | T2D | MR-RAPS | 16 | 1.23 (0.89 to 1.71) | 0.21 |  |  |  |  |
| Loneliness | T2D | PRESSO - raw | 16 | 1.19 (0.85 to 1.67) | 0.33 |  |  | Global = 111.57; p=<5e-04, Distortion = 76.99; p=0.15 |  |
| Loneliness | T2D | PRESSO - outlier corrected | 13 (3 removed) | 1.10 (0.90 to 1.36) | 0.37 |  |  |  |  |
| Loneliness | T2D | MRLAP - observed | 14 | 1.16 (1.00 to 1.34) | 0.05 |  |  |  |  |
| Loneliness | T2D | MRLAP - corrected | 14 | 1.23 (0.99 to 1.54) | 0.06 |  |  | Corrected vs observed estimate difference test = -1.69; p=0.09 |  |
| Loneliness | T2D | MR-CAUSE^a^ |  | 1.26 (1.04 to 1.51) |  |  |  | Causal vs sharing model ELPD: delta (SE) = -2.12 (1.70); p=0.11 |  |
| Loneliness | Suicide attempt | Inverse variance weighted | 14 | 2.42 (1.46 to 3.99) | 5.57e-04 | 2.58e-04 |  |  | 100.00 |
| Loneliness | Suicide attempt | MR-Egger | 14 | 7.10 (0.52 to 97.31) | 0.17 | 2.94e-04 | -0.02 (-0.08 to 0.03, 0.43) |  |  |
| Loneliness | Suicide attempt | SIMEX adjusted MR-Egger | 14 | 4.18 (0.29 to 61.01) | 0.32 |  | -0.01 (-0.07 to 0.05, 0.72) |  |  |
| Loneliness | Suicide attempt | Weighted median | 14 | 1.76 (1.05 to 2.93) | 0.03 |  |  |  |  |
| Loneliness | Suicide attempt | MR-RAPS | 14 | 2.60 (1.52 to 4.45) | 4.84e-04 |  |  |  |  |
| Loneliness | Suicide attempt | PRESSO - raw | 14 | 2.42 (1.46 to 3.99) | 4.30e-03 |  |  | Global = 44.86; p=<5e-04, Distortion = NA; p=NA |  |
| Loneliness | Suicide attempt | PRESSO - outlier corrected | 13 (1 removed) | NA (NA to NA) |  |  |  |  |  |
| Loneliness | Suicide attempt | MRLAP - observed | 13 | 1.30 (1.11 to 1.53) | 1.41e-03 |  |  |  |  |
| Loneliness | Suicide attempt | MRLAP - corrected | 13 | 1.48 (1.15 to 1.90) | 2.25e-03 |  |  | Corrected vs observed estimate difference test = -2.99; p=2.82e-03 |  |
| Loneliness | Suicide attempt | MR-CAUSE^a^ |  | 1.72 (1.28 to 2.29) |  |  |  | Causal vs sharing model ELPD: delta (SE) = -1.87 (1.81); p=0.15 |  |
| Loneliness | Depression | Inverse variance weighted | 16 | 2.13 (1.42 to 3.20) | 2.74e-04 | 4.77e-04 |  |  | 87.50 |
| Loneliness | Depression | MR-Egger | 16 | 0.52 (0.07 to 4.11) | 0.55 | 1.37e-03 | 0.03 (-0.01 to 0.07, 0.20) |  |  |
| Loneliness | Depression | SIMEX adjusted MR-Egger | 16 | 0.14 (0.02 to 0.99) | 0.07 |  | 0.06 (0.01 to 0.10, 0.02) |  |  |
| Loneliness | Depression | Weighted median | 16 | 2.08 (1.40 to 3.09) | 3.06e-04 |  |  |  |  |
| Loneliness | Depression | MR-RAPS | 16 | 1.95 (1.20 to 3.18) | 7.33e-03 |  |  |  |  |
| Loneliness | Depression | PRESSO - raw | 16 | 2.13 (1.42 to 3.20) | 2.43e-03 |  |  | Global = 45.25; p=2.00e-03, Distortion = 20.86; p=0.30 |  |
| Loneliness | Depression | PRESSO - outlier corrected | 15 (1 removed) | 1.87 (1.33 to 2.62) | 2.71e-03 |  |  |  |  |
| Loneliness | Depression | MRLAP - observed | 14 | 1.61 (1.21 to 2.15) | 1.23e-03 |  |  |  |  |
| Loneliness | Depression | MRLAP - corrected | 14 | 2.07 (1.33 to 3.21) | 1.25e-03 |  |  | Corrected vs observed estimate difference test = -3.30; p=9.81e-04 |  |
| Loneliness | Depression | MR-CAUSE^a^ |  | 1.72 (1.39 to 2.12) |  |  |  | Causal vs sharing model ELPD: delta (SE) = -5.37 (1.83); p=1.63e-03 |  |
| Loneliness | Anxiety | Inverse variance weighted | 16 | 1.91 (0.84 to 4.36) | 0.12 | 0.38 |  |  | 43.75 |
| Loneliness | Anxiety | MR-Egger | 16 | 0.50 (5.26e-03 to 47.21) | 0.77 | 0.33 | 0.03 (-0.06 to 0.12, 0.56) |  |  |
| Loneliness | Anxiety | SIMEX adjusted MR-Egger | 16 | 0.34 (4.44e-04 to 260.60) | 0.75 |  | 0.04 (-0.10 to 0.17, 0.61) |  |  |
| Loneliness | Anxiety | Weighted median | 16 | 0.99 (0.33 to 2.99) | 0.98 |  |  |  |  |
| Loneliness | Anxiety | MR-RAPS | 16 | 1.88 (0.79 to 4.46) | 0.15 |  |  |  |  |
| Loneliness | Anxiety | PRESSO - raw | 16 | 1.91 (0.84 to 4.36) | 0.14 |  |  | Global = 18.30; p=0.39, Distortion = NA; p=NA |  |
| Loneliness | Anxiety | PRESSO - outlier corrected | 16 (0 removed) | NA (NA to NA) |  |  |  |  |  |
| Loneliness | Anxiety | MRLAP - observed | 15 | 1.44 (0.85 to 2.41) | 0.17 |  |  |  |  |
| Loneliness | Anxiety | MRLAP - corrected | 15 | 1.76 (0.80 to 3.86) | 0.16 |  |  | Corrected vs observed estimate difference test = -1.52; p=0.13 |  |
| Loneliness | Anxiety | MR-CAUSE^a^ |  | 1.55 (0.91 to 2.51) |  |  |  | Causal vs sharing model ELPD: delta (SE) = -0.70 (1.27); p=0.29 |  |
| Loneliness | Wellbeing spectrum | Inverse variance weighted | 12 | -0.28 (-0.32 to -0.23) | 4.55e-33 | 6.50e-03 |  |  | 100.00 |
| Loneliness | Wellbeing spectrum | MR-Egger | 12 | -0.25 (-0.56 to 0.06) | 0.14 | 3.85e-03 | -5.19e-04 (-6.62e-03 to 5.58e-03, 0.87) |  |  |
| Loneliness | Wellbeing spectrum | SIMEX adjusted MR-Egger | 12 | -0.46 (-0.93 to 8.80e-03) | 0.08 |  | 3.52e-03 (-5.65e-03 to 0.01, 0.47) |  |  |
| Loneliness | Wellbeing spectrum | Weighted median | 12 | -0.25 (-0.30 to -0.19) | 1.68e-17 |  |  |  |  |
| Loneliness | Wellbeing spectrum | MR-RAPS | 12 | -0.29 (-0.33 to -0.24) | 0.00 |  |  |  |  |
| Loneliness | Wellbeing spectrum | PRESSO - raw | 12 | -0.28 (-0.32 to -0.23) | 1.18e-07 |  |  | Global = 30.85; p=0.22, Distortion = NA; p=NA |  |
| Loneliness | Wellbeing spectrum | PRESSO - outlier corrected | 12 (0 removed) | NA (NA to NA) |  |  |  |  |  |
| Loneliness | Wellbeing spectrum | MRLAP - observed | 9 | -0.42 (-0.51 to -0.33) | 4.26e-21 |  |  |  |  |
| Loneliness | Wellbeing spectrum | MRLAP - corrected | 9 | -0.57 (-0.72 to -0.43) | 1.58e-15 |  |  | Corrected vs observed estimate difference test = 5.40; p=6.71e-08 |  |
| Loneliness | Wellbeing spectrum | MR-CAUSE^a^ |  | -0.23 (-0.27 to -0.19) |  |  |  | Causal vs sharing model ELPD: delta (SE) = -7.87 (1.29); p=5.23e-10 |  |
| Loneliness | Positive affect | Inverse variance weighted | 12 | -0.35 (-0.46 to -0.24) | 3.37e-10 | 6.02e-03 |  |  | 91.67 |
| Loneliness | Positive affect | MR-Egger | 12 | -0.34 (-1.08 to 0.39) | 0.38 | 3.45e-03 | -6.41e-05 (-0.01 to 0.01, 0.99) |  |  |
| Loneliness | Positive affect | SIMEX adjusted MR-Egger | 12 | -0.62 (-1.75 to 0.51) | 0.31 |  | 5.25e-03 (-0.02 to 0.03, 0.65) |  |  |
| Loneliness | Positive affect | Weighted median | 12 | -0.30 (-0.40 to -0.20) | 3.89e-09 |  |  |  |  |
| Loneliness | Positive affect | MR-RAPS | 12 | -0.31 (-0.39 to -0.23) | 7.77e-15 |  |  |  |  |
| Loneliness | Positive affect | PRESSO - raw | 12 | -0.35 (-0.46 to -0.24) | 6.00e-05 |  |  | Global = 30.81; p=0.03, Distortion = -15.38; p=0.04 |  |
| Loneliness | Positive affect | PRESSO - outlier corrected | 11 (1 removed) | -0.30 (-0.36 to -0.25) | 9.17e-07 |  |  |  |  |
| Loneliness | Positive affect | MRLAP - observed | 9 | -0.61 (-0.81 to -0.41) | 3.43e-09 |  |  |  |  |
| Loneliness | Positive affect | MRLAP - corrected | 9 | -0.87 (-1.20 to -0.55) | 1.15e-07 |  |  | Corrected vs observed estimate difference test = 3.97; p=7.26e-05 |  |
| Loneliness | Positive affect | MR-CAUSE^a^ |  | -0.23 (-0.29 to -0.16) |  |  |  | Causal vs sharing model ELPD: delta (SE) = -5.89 (1.78); p=4.85e-04 |  |
| Loneliness | Life satisfaction | Inverse variance weighted | 12 | -0.47 (-0.69 to -0.24) | 4.28e-05 | 0.02 |  |  | 58.33 |
| Loneliness | Life satisfaction | MR-Egger | 12 | 0.28 (-1.17 to 1.73) | 0.71 | 0.03 | -0.01 (-0.04 to 0.01, 0.33) |  |  |
| Loneliness | Life satisfaction | SIMEX adjusted MR-Egger | 12 | 0.51 (-1.72 to 2.75) | 0.66 |  | -0.02 (-0.06 to 0.02, 0.40) |  |  |
| Loneliness | Life satisfaction | Weighted median | 12 | -0.29 (-0.52 to -0.05) | 0.02 |  |  |  |  |
| Loneliness | Life satisfaction | MR-RAPS | 12 | -0.46 (-0.70 to -0.22) | 1.61e-04 |  |  |  |  |
| Loneliness | Life satisfaction | PRESSO - raw | 12 | -0.47 (-0.69 to -0.24) | 1.78e-03 |  |  | Global = 26.48; p=0.04, Distortion = -16.63; p=0.37 |  |
| Loneliness | Life satisfaction | PRESSO - outlier corrected | 11 (1 removed) | -0.40 (-0.60 to -0.21) | 2.28e-03 |  |  |  |  |
| Loneliness | Life satisfaction | MRLAP - observed | 9 | -0.77 (-1.16 to -0.38) | 1.14e-04 |  |  |  |  |
| Loneliness | Life satisfaction | MRLAP - corrected | 9 | -1.26 (-1.88 to -0.63) | 8.06e-05 |  |  | Corrected vs observed estimate difference test = 3.79; p=1.49e-04 |  |
| Loneliness | Life satisfaction | MR-CAUSE^a^ |  | -0.22 (-0.34 to -0.08) |  |  |  | Causal vs sharing model ELPD: delta (SE) = -1.58 (1.79); p=0.19 |  |
| Social isolation | CAD | Inverse variance weighted | 15 | 0.66 (0.25 to 1.79) | 0.42 | 7.01e-04 |  |  | 100.00 |
| Social isolation | CAD | MR-Egger | 15 | 0.03 (3.64e-04 to 2.93) | 0.16 | 1.95e-03 | 0.01 (-6.50e-03 to 0.03, 0.20) |  |  |
| Social isolation | CAD | SIMEX adjusted MR-Egger | 15 | 0.01 (2.26e-04 to 0.79) | 0.06 |  | 0.02 (-1.07e-03 to 0.04, 0.09) |  |  |
| Social isolation | CAD | Weighted median | 15 | 0.42 (0.17 to 1.06) | 0.07 |  |  |  |  |
| Social isolation | CAD | MR-RAPS | 15 | 0.57 (0.20 to 1.58) | 0.28 |  |  |  |  |
| Social isolation | CAD | PRESSO - raw | 15 | 0.66 (0.25 to 1.79) | 0.43 |  |  | Global = 42.92; p=<5e-04, Distortion = -13.91; p=0.87 |  |
| Social isolation | CAD | PRESSO - outlier corrected | 13 (2 removed) | 0.70 (0.29 to 1.68) | 0.44 |  |  |  |  |
| Social isolation | CAD | MRLAP - observed | 16 | 0.96 (0.88 to 1.04) | 0.30 |  |  |  |  |
| Social isolation | CAD | MRLAP - corrected | 16 | 0.94 (0.84 to 1.05) | 0.29 |  |  | Corrected vs observed estimate difference test = 1.00; p=0.32 |  |
| Social isolation | CAD | MR-CAUSE^a^ |  | 1.00 (0.53 to 1.84) |  |  |  | Causal vs sharing model ELPD: delta (SE) = 0.89 (0.07); p=1.00 |  |
| Social isolation | Systolic blood pressure | Inverse variance weighted | 14 | 1.80 (-10.71 to 14.31) | 0.78 | 2.70e-23 |  |  | 100.00 |
| Social isolation | Systolic blood pressure | MR-Egger | 14 | -30.28 (-85.70 to 25.14) | 0.31 | 5.36e-21 | 0.15 (-0.11 to 0.41, 0.27) |  |  |
| Social isolation | Systolic blood pressure | SIMEX adjusted MR-Egger | 14 | -50.99 (-102.65 to 0.68) | 0.08 |  | 0.25 (-0.01 to 0.51, 0.09) |  |  |
| Social isolation | Systolic blood pressure | Weighted median | 14 | 1.44 (-5.73 to 8.62) | 0.69 |  |  |  |  |
| Social isolation | Systolic blood pressure | MR-RAPS | 14 | -2.64 (-16.77 to 11.49) | 0.71 |  |  |  |  |
| Social isolation | Systolic blood pressure | PRESSO - raw | 14 | 1.80 (-10.71 to 14.31) | 0.78 |  |  | Global = 166.46; p=<5e-04, Distortion = -71.24; p=0.64 |  |
| Social isolation | Systolic blood pressure | PRESSO - outlier corrected | 10 (4 removed) | 6.26 (-0.96 to 13.48) | 0.12 |  |  |  |  |
| Social isolation | Systolic blood pressure | MRLAP - observed | 15 | -5.76e-04 (-0.22 to 0.21) | 1.00 |  |  |  |  |
| Social isolation | Systolic blood pressure | MRLAP - corrected | 15 | 3.81e-04 (-0.31 to 0.31) | 1.00 |  |  | Corrected vs observed estimate difference test = -0.02; p=0.98 |  |
| Social isolation | Systolic blood pressure | MR-CAUSE^a^ |  | 2.13 (-3.66 to 7.91) |  |  |  | Causal vs sharing model ELPD: delta (SE) = 0.41 (0.55); p=0.77 |  |
| Social isolation | Heart failure | Inverse variance weighted | 15 | 0.63 (0.18 to 2.22) | 0.48 | 0.07 |  |  | 100.00 |
| Social isolation | Heart failure | MR-Egger | 15 | 0.16 (4.28e-04 to 58.58) | 0.55 | 0.06 | 6.67e-03 (-0.02 to 0.03, 0.65) |  |  |
| Social isolation | Heart failure | SIMEX adjusted MR-Egger | 15 | 0.07 (2.59e-05 to 204.68) | 0.53 |  | 0.01 (-0.03 to 0.05, 0.60) |  |  |
| Social isolation | Heart failure | Weighted median | 15 | 0.70 (0.18 to 2.75) | 0.61 |  |  |  |  |
| Social isolation | Heart failure | MR-RAPS | 15 | 0.66 (0.19 to 2.28) | 0.51 |  |  |  |  |
| Social isolation | Heart failure | PRESSO - raw | 15 | 0.63 (0.18 to 2.22) | 0.49 |  |  | Global = 25.28; p=0.08, Distortion = NA; p=NA |  |
| Social isolation | Heart failure | PRESSO - outlier corrected | 15 (0 removed) | NA (NA to NA) |  |  |  |  |  |
| Social isolation | Heart failure | MRLAP - observed | 15 | 0.98 (0.91 to 1.05) | 0.52 |  |  |  |  |
| Social isolation | Heart failure | MRLAP - corrected | 15 | 0.96 (0.86 to 1.07) | 0.48 |  |  | Corrected vs observed estimate difference test = 0.81; p=0.42 |  |
| Social isolation | Heart failure | MR-CAUSE^a^ |  | 0.88 (0.41 to 1.88) |  |  |  | Causal vs sharing model ELPD: delta (SE) = 0.76 (0.29); p=1.00 |  |
| Social isolation | Stroke | Inverse variance weighted | 15 | 1.32 (0.42 to 4.15) | 0.64 | 0.92 |  |  | 50.00 |
| Social isolation | Stroke | MR-Egger | 15 | 4.25 (0.02 to 786.29) | 0.60 | 0.89 | -5.57e-03 (-0.03 to 0.02, 0.66) |  |  |
| Social isolation | Stroke | SIMEX adjusted MR-Egger | 15 | 11.02 (0.28 to 431.28) | 0.22 |  | -9.82e-03 (-0.03 to 8.38e-03, 0.31) |  |  |
| Social isolation | Stroke | Weighted median | 15 | 0.76 (0.17 to 3.39) | 0.71 |  |  |  |  |
| Social isolation | Stroke | MR-RAPS | 15 | 1.29 (0.38 to 4.35) | 0.68 |  |  |  |  |
| Social isolation | Stroke | PRESSO - raw | 15 | 1.32 (0.57 to 3.04) | 0.53 |  |  | Global = 8.73; p=0.92, Distortion = NA; p=NA |  |
| Social isolation | Stroke | PRESSO - outlier corrected | 15 (0 removed) | NA (NA to NA) |  |  |  |  |  |
| Social isolation | Stroke | MRLAP - observed | 15 | 1.01 (0.96 to 1.06) | 0.63 |  |  |  |  |
| Social isolation | Stroke | MRLAP - corrected | 15 | 1.02 (0.95 to 1.09) | 0.67 |  |  | Corrected vs observed estimate difference test = -0.31; p=0.76 |  |
| Social isolation | Stroke | MR-CAUSE^a^ |  | 1.07 (0.48 to 2.39) |  |  |  | Causal vs sharing model ELPD: delta (SE) = 0.88 (0.11); p=1.00 |  |
| Social isolation | T2D | Inverse variance weighted | 15 | 0.95 (0.14 to 6.31) | 0.96 | 3.66e-23 |  |  | 100.00 |
| Social isolation | T2D | MR-Egger | 15 | 1.68 (1.86e-04 to 15287.64) | 0.91 | 1.17e-23 | -2.67e-03 (-0.04 to 0.04, 0.90) |  |  |
| Social isolation | T2D | SIMEX adjusted MR-Egger | 15 | 0.04 (1.24e-05 to 135.00) | 0.45 |  | 0.01 (-0.03 to 0.05, 0.50) |  |  |
| Social isolation | T2D | Weighted median | 15 | 1.30 (0.48 to 3.47) | 0.61 |  |  |  |  |
| Social isolation | T2D | MR-RAPS | 15 | 0.76 (0.14 to 4.23) | 0.75 |  |  |  |  |
| Social isolation | T2D | PRESSO - raw | 15 | 0.95 (0.14 to 6.31) | 0.96 |  |  | Global = 160.73; p=<5e-04, Distortion = -254.40; p=0.04 |  |
| Social isolation | T2D | PRESSO - outlier corrected | 13 (2 removed) | 1.03 (0.36 to 2.96) | 0.95 |  |  |  |  |
| Social isolation | T2D | MRLAP - observed | 16 | 0.99 (0.84 to 1.17) | 0.90 |  |  |  |  |
| Social isolation | T2D | MRLAP - corrected | 16 | 0.98 (0.77 to 1.24) | 0.88 |  |  | Corrected vs observed estimate difference test = 0.20; p=0.84 |  |
| Social isolation | T2D | MR-CAUSE^a^ |  | 0.87 (0.41 to 1.82) |  |  |  | Causal vs sharing model ELPD: delta (SE) = 0.70 (0.31); p=0.99 |  |
| Social isolation | Suicide attempt | Inverse variance weighted | 12 | 4.85 (0.26 to 89.71) | 0.29 | 1.16e-07 |  |  | 100.00 |
| Social isolation | Suicide attempt | MR-Egger | 12 | 3.51e-03 (8.10e-09 to 1519.03) | 0.41 | 6.17e-07 | 0.04 (-0.03 to 0.10, 0.29) |  |  |
| Social isolation | Suicide attempt | SIMEX adjusted MR-Egger | 12 | 0.27 (2.13e-06 to 34280.17) | 0.83 |  | 0.02 (-0.05 to 0.08, 0.63) |  |  |
| Social isolation | Suicide attempt | Weighted median | 12 | 1.22 (0.16 to 8.98) | 0.85 |  |  |  |  |
| Social isolation | Suicide attempt | MR-RAPS | 12 | 2.90 (0.32 to 26.58) | 0.35 |  |  |  |  |
| Social isolation | Suicide attempt | PRESSO - raw | 12 | 4.85 (0.26 to 89.71) | 0.31 |  |  | Global = 65.25; p=<5e-04, Distortion = 16.17; p=0.72 |  |
| Social isolation | Suicide attempt | PRESSO - outlier corrected | 10 (2 removed) | 3.89 (0.72 to 21.02) | 0.15 |  |  |  |  |
| Social isolation | Suicide attempt | MRLAP - observed | 15 | 1.11 (0.94 to 1.31) | 0.23 |  |  |  |  |
| Social isolation | Suicide attempt | MRLAP - corrected | 15 | 1.15 (0.90 to 1.47) | 0.26 |  |  | Corrected vs observed estimate difference test = -0.99; p=0.32 |  |
| Social isolation | Suicide attempt | MR-CAUSE^a^ |  | 2.94 (1.02 to 8.33) |  |  |  | Causal vs sharing model ELPD: delta (SE) = -1.27 (1.55); p=0.21 |  |
| Social isolation | Depression | Inverse variance weighted | 15 | 1.42 (0.22 to 9.21) | 0.72 | 1.91e-04 |  |  | 100.00 |
| Social isolation | Depression | MR-Egger | 15 | 0.12 (1.71e-05 to 826.61) | 0.65 | 1.47e-04 | 0.01 (-0.03 to 0.05, 0.58) |  |  |
| Social isolation | Depression | SIMEX adjusted MR-Egger | 15 | 0.30 (3.58e-05 to 2501.49) | 0.80 |  | 8.01e-03 (-0.04 to 0.05, 0.73) |  |  |
| Social isolation | Depression | Weighted median | 15 | 2.18 (0.44 to 10.88) | 0.34 |  |  |  |  |
| Social isolation | Depression | MR-RAPS | 15 | 1.66 (0.26 to 10.75) | 0.60 |  |  |  |  |
| Social isolation | Depression | PRESSO - raw | 15 | 1.42 (0.22 to 9.21) | 0.72 |  |  | Global = 48.09; p=<5e-04, Distortion = -30.19; p=0.85 |  |
| Social isolation | Depression | PRESSO - outlier corrected | 13 (2 removed) | 1.65 (0.38 to 7.09) | 0.52 |  |  |  |  |
| Social isolation | Depression | MR-CAUSE^a^ |  | 1.97 (0.77 to 5.37) |  |  |  | Causal vs sharing model ELPD: delta (SE) = -0.14 (1.21); p=0.45 |  |
| Social isolation | Anxiety | Inverse variance weighted | 15 | 0.42 (0.01 to 13.32) | 0.62 | 0.73 |  |  | 80.00 |
| Social isolation | Anxiety | MR-Egger | 15 | 0.02 (1.97e-09 to 124753.93) | 0.62 | 0.67 | 0.02 (-0.06 to 0.09, 0.68) |  |  |
| Social isolation | Anxiety | SIMEX adjusted MR-Egger | 15 | 2.28e-03 (1.84e-11 to 283669.57) | 0.53 |  | 0.02 (-0.06 to 0.11, 0.58) |  |  |
| Social isolation | Anxiety | Weighted median | 15 | 0.30 (3.25e-03 to 27.96) | 0.60 |  |  |  |  |
| Social isolation | Anxiety | MR-RAPS | 15 | 0.45 (0.01 to 17.50) | 0.67 |  |  |  |  |
| Social isolation | Anxiety | PRESSO - raw | 15 | 0.42 (0.02 to 8.30) | 0.58 |  |  | Global = 11.70; p=0.74, Distortion = NA; p=NA |  |
| Social isolation | Anxiety | PRESSO - outlier corrected | 15 (0 removed) | NA (NA to NA) |  |  |  |  |  |
| Social isolation | Anxiety | MR-CAUSE^a^ |  | 0.31 (0.02 to 4.57) |  |  |  | Causal vs sharing model ELPD: delta (SE) = 0.43 (0.83); p=0.70 |  |
| Social isolation | Wellbeing spectrum | Inverse variance weighted | 14 | -0.30 (-0.65 to 0.04) | 0.08 | 6.39e-18 |  |  | 100.00 |
| Social isolation | Wellbeing spectrum | MR-Egger | 14 | -1.01 (-2.55 to 0.52) | 0.22 | 6.12e-17 | 3.38e-03 (-3.78e-03 to 0.01, 0.37) |  |  |
| Social isolation | Wellbeing spectrum | SIMEX adjusted MR-Egger | 14 | -0.82 (-2.23 to 0.59) | 0.27 |  | 2.62e-03 (-4.40e-03 to 9.65e-03, 0.48) |  |  |
| Social isolation | Wellbeing spectrum | Weighted median | 14 | -0.30 (-0.52 to -0.08) | 8.06e-03 |  |  |  |  |
| Social isolation | Wellbeing spectrum | MR-RAPS | 14 | -0.35 (-0.64 to -0.05) | 0.02 |  |  |  |  |
| Social isolation | Wellbeing spectrum | PRESSO - raw | 14 | -0.30 (-0.65 to 0.04) | 0.11 |  |  | Global = 129.90; p=<5e-04, Distortion = 21.15; p=0.49 |  |
| Social isolation | Wellbeing spectrum | PRESSO - outlier corrected | 9 (5 removed) | -0.39 (-0.63 to -0.14) | 0.01 |  |  |  |  |
| Social isolation | Wellbeing spectrum | MRLAP - observed | 14 | -0.10 (-0.21 to 0.01) | 0.08 |  |  |  |  |
| Social isolation | Wellbeing spectrum | MRLAP - corrected | 14 | -0.13 (-0.29 to 0.04) | 0.14 |  |  | Corrected vs observed estimate difference test = 1.05; p=0.30 |  |
| Social isolation | Wellbeing spectrum | MR-CAUSE^a^ |  | -0.26 (-0.39 to -0.12) |  |  |  | Causal vs sharing model ELPD: delta (SE) = -4.44 (2.05); p=0.02 |  |
| Social isolation | Positive affect | Inverse variance weighted | 14 | -0.69 (-1.28 to -0.11) | 0.02 | 1.47e-07 |  |  | 100.00 |
| Social isolation | Positive affect | MR-Egger | 14 | -2.02 (-4.62 to 0.58) | 0.15 | 4.33e-07 | 6.32e-03 (-5.79e-03 to 0.02, 0.33) |  |  |
| Social isolation | Positive affect | SIMEX adjusted MR-Egger | 14 | -0.85 (-3.73 to 2.02) | 0.57 |  | 1.16e-03 (-0.01 to 0.02, 0.88) |  |  |
| Social isolation | Positive affect | Weighted median | 14 | -0.30 (-0.75 to 0.14) | 0.18 |  |  |  |  |
| Social isolation | Positive affect | MR-RAPS | 14 | -0.39 (-0.74 to -0.03) | 0.03 |  |  |  |  |
| Social isolation | Positive affect | PRESSO - raw | 14 | -0.69 (-1.28 to -0.11) | 0.04 |  |  | Global = 69.75; p=<5e-04, Distortion = -78.10; p=0.02 |  |
| Social isolation | Positive affect | PRESSO - outlier corrected | 13 (1 removed) | -0.39 (-0.78 to -2.34e-03) | 0.07 |  |  |  |  |
| Social isolation | Positive affect | MRLAP - observed | 14 | -0.22 (-0.41 to -0.04) | 0.02 |  |  |  |  |
| Social isolation | Positive affect | MRLAP - corrected | 14 | -0.29 (-0.57 to -0.02) | 0.03 |  |  | Corrected vs observed estimate difference test = 1.47; p=0.14 |  |
| Social isolation | Positive affect | MR-CAUSE^a^ |  | -0.46 (-0.69 to -0.22) |  |  |  | Causal vs sharing model ELPD: delta (SE) = -4.58 (2.05); p=0.01 |  |
| Social isolation | Life satisfaction | Inverse variance weighted | 14 | -0.40 (-1.03 to 0.23) | 0.21 | 0.44 |  |  | 100.00 |
| Social isolation | Life satisfaction | MR-Egger | 14 | -2.37 (-5.18 to 0.44) | 0.12 | 0.51 | 9.37e-03 (-3.67e-03 to 0.02, 0.18) |  |  |
| Social isolation | Life satisfaction | SIMEX adjusted MR-Egger | 14 | -1.57 (-4.40 to 1.26) | 0.30 |  | 5.98e-03 (-8.06e-03 to 0.02, 0.42) |  |  |
| Social isolation | Life satisfaction | Weighted median | 14 | -0.70 (-1.57 to 0.17) | 0.12 |  |  |  |  |
| Social isolation | Life satisfaction | MR-RAPS | 14 | -0.46 (-1.12 to 0.21) | 0.18 |  |  |  |  |
| Social isolation | Life satisfaction | PRESSO - raw | 14 | -0.40 (-1.03 to 0.23) | 0.23 |  |  | Global = 15.43; p=0.44, Distortion = NA; p=NA |  |
| Social isolation | Life satisfaction | PRESSO - outlier corrected | 14 (0 removed) | NA (NA to NA) |  |  |  |  |  |
| Social isolation | Life satisfaction | MR-CAUSE^a^ |  | -0.42 (-0.82 to -0.03) |  |  |  | Causal vs sharing model ELPD: delta (SE) = -1.32 (1.66); p=0.21 |  |

*OR=odds ratio, CI=confidence intervals, MR=Mendelian Randomisation, SIMEX=* *simulation extrapolation, MR-RAPS=* *MR using robust adjusted profile score, PRESSO=Pleiotropy RESidual Sum and Outlier, CAD=coronary artery disease, HF=heart failure, T2D=type 2 diabetes, SBP=systolic blood pressure, ELPD=expected log pointwise posterior density, SE=standard error*

*^a^ MR-CAUSE is a Bayesian method so the values reported are the posterior median as the estimate and the posterior credible interval as the CI.*

### Table S15. Two-sample Mendelian Randomisation analysis results with loneliness and social isolation as outcomes

| **Exposure** | **Outcome** | **Method** | **N genetic variants** | **OR or mean difference (95% CI)** | **p-value** | **Heterogeneity test p-value** | **Directional pleiotropy intercept (95% CI; p-value)** | **Additional tests for MR approaches** | **Steiger filtering - percentage of genetic variants** |
| --- | --- | --- | --- | --- | --- | --- | --- | --- | --- |
| CAD | Loneliness | Inverse variance weighted | 137 | 1.00 (0.99 to 1.02) | 0.57 | 1.11e-06 |  |  | 100.00 |
| CAD | Loneliness | MR-Egger | 137 | 1.00 (0.97 to 1.02) | 0.80 | 9.72e-07 | 5.00e-04 (-1.04e-03 to 2.04e-03, 0.53) |  |  |
| CAD | Loneliness | SIMEX adjusted MR-Egger | 137 | 1.01 (0.98 to 1.03) | 0.54 |  | 1.48e-04 (-1.55e-03 to 1.85e-03, 0.86) |  |  |
| CAD | Loneliness | Weighted median | 137 | 1.00 (0.98 to 1.02) | 0.78 |  |  |  |  |
| CAD | Loneliness | MR-RAPS | 137 | 1.00 (0.99 to 1.02) | 0.54 |  |  |  |  |
| CAD | Loneliness | PRESSO - raw | 137 | 1.00 (0.99 to 1.02) | 0.57 |  |  | Global = 232.12; p=<0.0003, Distortion = -45.73; p=0.70 |  |
| CAD | Loneliness | PRESSO - outlier corrected | 134 (3 removed) | 1.01 (0.99 to 1.02) | 0.27 |  |  |  |  |
| CAD | Loneliness | MRLAP - observed | 177 | 1.02 (0.99 to 1.05) | 0.17 |  |  |  |  |
| CAD | Loneliness | MRLAP - corrected | 177 | 1.02 (0.98 to 1.06) | 0.30 |  |  | Corrected vs observed estimate difference test = 1.19; p=0.23 |  |
| CAD | Loneliness | MR-CAUSE^a^ |  | 1.01 (1.00 to 1.03) |  |  |  | Causal vs sharing model ELPD: delta (SE) = -0.83 (1.39); p=0.28 |  |
| CAD | Social isolation | Inverse variance weighted | 138 | -6.70e-04 (-3.51e-03 to 2.17e-03) | 0.64 | 1.46e-03 |  |  | 100.00 |
| CAD | Social isolation | MR-Egger | 138 | 3.48e-03 (-2.16e-03 to 9.11e-03) | 0.23 | 2.21e-03 | -2.70e-04 (-5.87e-04 to 4.73e-05, 0.10) |  |  |
| CAD | Social isolation | SIMEX adjusted MR-Egger | 138 | 2.92e-03 (-2.06e-03 to 7.91e-03) | 0.25 |  | -2.07e-04 (-5.50e-04 to 1.36e-04, 0.24) |  |  |
| CAD | Social isolation | Weighted median | 138 | 1.27e-03 (-3.13e-03 to 5.67e-03) | 0.57 |  |  |  |  |
| CAD | Social isolation | MR-RAPS | 138 | -5.53e-04 (-3.29e-03 to 2.19e-03) | 0.69 |  |  |  |  |
| CAD | Social isolation | PRESSO - raw | 138 | -6.70e-04 (-3.51e-03 to 2.17e-03) | 0.64 |  |  | Global = 194.40; p=3.33e-04, Distortion = -13.97; p=0.95 |  |
| CAD | Social isolation | PRESSO - outlier corrected | 136 (2 removed) | -5.88e-04 (-3.25e-03 to 2.08e-03) | 0.67 |  |  |  |  |
| CAD | Social isolation | MRLAP - observed | 177 | -3.46e-03 (-0.03 to 0.03) | 0.83 |  |  |  |  |
| CAD | Social isolation | MRLAP - corrected | 177 | -4.28e-03 (-0.04 to 0.03) | 0.82 |  |  | Corrected vs observed estimate difference test = 2.79; p=5.20e-03 |  |
| CAD | Social isolation | MR-CAUSE^a^ |  | 0.00 (0.00 to 0.00) |  |  |  | Causal vs sharing model ELPD: delta (SE) = 0.97 (0.36); p=1.00 |  |
| Systolic blood pressure | Loneliness | Inverse variance weighted | 94 | 1.00 (1.00 to 1.00) | 0.78 | 3.59e-04 |  |  | 100.00 |
| Systolic blood pressure | Loneliness | MR-Egger | 94 | 1.00 (0.99 to 1.02) | 0.58 | 3.22e-04 | -1.08e-03 (-4.27e-03 to 2.11e-03, 0.51) |  |  |
| Systolic blood pressure | Loneliness | SIMEX adjusted MR-Egger | 94 | 1.01 (0.99 to 1.02) | 0.54 |  | -1.35e-03 (-5.25e-03 to 2.55e-03, 0.50) |  |  |
| Systolic blood pressure | Loneliness | Weighted median | 94 | 1.00 (1.00 to 1.00) | 0.90 |  |  |  |  |
| Systolic blood pressure | Loneliness | MR-RAPS | 94 | 1.00 (1.00 to 1.00) | 0.95 |  |  |  |  |
| Systolic blood pressure | Loneliness | PRESSO - raw | 94 | 1.00 (1.00 to 1.00) | 0.78 |  |  | Global = 149.16; p=<5e-04, Distortion = NA; p=NA |  |
| Systolic blood pressure | Loneliness | PRESSO - outlier corrected | 93 (1 removed) | NA (NA to NA) |  |  |  |  |  |
| Systolic blood pressure | Loneliness | MRLAP - observed | 400 | 0.99 (0.97 to 1.00) | 0.16 |  |  |  |  |
| Systolic blood pressure | Loneliness | MRLAP - corrected | 400 | 0.99 (0.97 to 1.01) | 0.29 |  |  | Corrected vs observed estimate difference test = -0.70; p=0.48 |  |
| Systolic blood pressure | Loneliness | MR-CAUSE^a^ |  | 1.00 (1.00 to 1.00) |  |  |  | Causal vs sharing model ELPD: delta (SE) = 0.53 (0.79); p=0.75 |  |
| Systolic blood pressure | Social isolation | Inverse variance weighted | 96 | -7.03e-04 (-1.50e-03 to 9.54e-05) | 0.08 | 2.76e-03 |  |  | 100.00 |
| Systolic blood pressure | Social isolation | MR-Egger | 96 | -1.78e-04 (-3.29e-03 to 2.94e-03) | 0.91 | 2.30e-03 | -1.30e-04 (-8.74e-04 to 6.14e-04, 0.73) |  |  |
| Systolic blood pressure | Social isolation | SIMEX adjusted MR-Egger | 96 | -1.65e-03 (-3.56e-03 to 2.58e-04) | 0.09 |  | 2.22e-04 (-3.29e-04 to 7.73e-04, 0.43) |  |  |
| Systolic blood pressure | Social isolation | Weighted median | 96 | -8.82e-04 (-1.83e-03 to 6.98e-05) | 0.07 |  |  |  |  |
| Systolic blood pressure | Social isolation | MR-RAPS | 96 | -8.56e-04 (-1.56e-03 to -1.50e-04) | 0.02 |  |  |  |  |
| Systolic blood pressure | Social isolation | PRESSO - raw | 96 | -7.03e-04 (-1.50e-03 to 9.54e-05) | 0.09 |  |  | Global = 140.83; p=2.00e-03, Distortion = 8.50; p=0.87 |  |
| Systolic blood pressure | Social isolation | PRESSO - outlier corrected | 94 (2 removed) | -7.69e-04 (-1.47e-03 to -6.96e-05) | 0.03 |  |  |  |  |
| Systolic blood pressure | Social isolation | MRLAP - observed | 441 | 0.02 (-1.77e-03 to 0.03) | 0.08 |  |  |  |  |
| Systolic blood pressure | Social isolation | MRLAP - corrected | 441 | 0.02 (-1.59e-03 to 0.04) | 0.07 |  |  | Corrected vs observed estimate difference test = -1.63; p=0.10 |  |
| Systolic blood pressure | Social isolation | MR-CAUSE^a^ |  | 0.00 (0.00 to 0.00) |  |  |  | Causal vs sharing model ELPD: delta (SE) = 0.66 (0.55); p=0.89 |  |
| Heart failure | Loneliness | Inverse variance weighted | 9 | 1.00 (0.94 to 1.06) | 0.87 | 0.02 |  |  | 100.00 |
| Heart failure | Loneliness | MR-Egger | 9 | 0.89 (0.73 to 1.08) | 0.26 | 0.03 | 7.66e-03 (-4.68e-03 to 0.02, 0.26) |  |  |
| Heart failure | Loneliness | SIMEX adjusted MR-Egger | 9 | 0.96 (0.79 to 1.17) | 0.70 |  | 1.40e-03 (-0.01 to 0.01, 0.84) |  |  |
| Heart failure | Loneliness | Weighted median | 9 | 0.98 (0.93 to 1.04) | 0.56 |  |  |  |  |
| Heart failure | Loneliness | MR-RAPS | 9 | 0.99 (0.94 to 1.04) | 0.68 |  |  |  |  |
| Heart failure | Loneliness | PRESSO - raw | 9 | 1.00 (0.94 to 1.06) | 0.87 |  |  | Global = 22.64; p=0.03, Distortion = -222.74; p=0.12 |  |
| Heart failure | Loneliness | PRESSO - outlier corrected | 7 (2 removed) | 1.00 (0.96 to 1.04) | 0.94 |  |  |  |  |
| Heart failure | Loneliness | MRLAP - observed | 12 | 1.01 (0.82 to 1.25) | 0.91 |  |  |  |  |
| Heart failure | Loneliness | MRLAP - corrected | 12 | 1.01 (0.75 to 1.35) | 0.96 |  |  | Corrected vs observed estimate difference test = 0.11; p=0.91 |  |
| Heart failure | Loneliness | MR-CAUSE^a^ |  | 1.00 (0.97 to 1.04) |  |  |  | Causal vs sharing model ELPD: delta (SE) = 0.87 (0.18); p=1.00 |  |
| Heart failure | Social isolation | Inverse variance weighted | 9 | 4.39e-03 (-4.30e-03 to 0.01) | 0.32 | 0.68 |  |  | 100.00 |
| Heart failure | Social isolation | MR-Egger | 9 | 4.61e-03 (-0.03 to 0.03) | 0.77 | 0.57 | -1.44e-05 (-1.89e-03 to 1.86e-03, 0.99) |  |  |
| Heart failure | Social isolation | SIMEX adjusted MR-Egger | 9 | 2.90e-03 (-0.02 to 0.03) | 0.81 |  | 1.87e-05 (-1.55e-03 to 1.59e-03, 0.98) |  |  |
| Heart failure | Social isolation | Weighted median | 9 | 4.13e-03 (-7.25e-03 to 0.02) | 0.48 |  |  |  |  |
| Heart failure | Social isolation | MR-RAPS | 9 | 3.08e-03 (-5.11e-03 to 0.01) | 0.46 |  |  |  |  |
| Heart failure | Social isolation | PRESSO - raw | 9 | 4.39e-03 (-2.96e-03 to 0.01) | 0.28 |  |  | Global = 7.00; p=0.71, Distortion = NA; p=NA |  |
| Heart failure | Social isolation | PRESSO - outlier corrected | 9 (0 removed) | NA (NA to NA) |  |  |  |  |  |
| Heart failure | Social isolation | MRLAP - observed | 12 | 0.09 (-0.04 to 0.22) | 0.16 |  |  |  |  |
| Heart failure | Social isolation | MRLAP - corrected | 12 | 0.12 (-0.05 to 0.29) | 0.18 |  |  | Corrected vs observed estimate difference test = -1.19; p=0.23 |  |
| Heart failure | Social isolation | MR-CAUSE^a^ |  | 0.01 (0.00 to 0.01) |  |  |  | Causal vs sharing model ELPD: delta (SE) = -0.98 (1.51); p=0.26 |  |
| Stroke | Loneliness | Inverse variance weighted | 25 | 0.99 (0.95 to 1.02) | 0.52 | 5.73e-03 |  |  | 92.00 |
| Stroke | Loneliness | MR-Egger | 25 | 0.95 (0.85 to 1.06) | 0.35 | 5.47e-03 | 2.30e-03 (-3.38e-03 to 7.98e-03, 0.44) |  |  |
| Stroke | Loneliness | SIMEX adjusted MR-Egger | 25 | 0.91 (0.78 to 1.06) | 0.26 |  | 4.28e-03 (-3.89e-03 to 0.01, 0.32) |  |  |
| Stroke | Loneliness | Weighted median | 25 | 0.99 (0.96 to 1.03) | 0.77 |  |  |  |  |
| Stroke | Loneliness | MR-RAPS | 25 | 0.99 (0.95 to 1.03) | 0.62 |  |  |  |  |
| Stroke | Loneliness | PRESSO - raw | 25 | 0.99 (0.95 to 1.02) | 0.53 |  |  | Global = 49.43; p=6.00e-03, Distortion = -250.78; p=0.19 |  |
| Stroke | Loneliness | PRESSO - outlier corrected | 23 (2 removed) | 1.00 (0.97 to 1.02) | 0.81 |  |  |  |  |
| Stroke | Loneliness | MRLAP - observed | 25 | 0.95 (0.80 to 1.13) | 0.56 |  |  |  |  |
| Stroke | Loneliness | MRLAP - corrected | 25 | 0.93 (0.74 to 1.17) | 0.56 |  |  | Corrected vs observed estimate difference test = 0.85; p=0.40 |  |
| Stroke | Loneliness | MR-CAUSE^a^ |  | 1.01 (0.97 to 1.04) |  |  |  | Causal vs sharing model ELPD: delta (SE) = 0.71 (0.31); p=0.99 |  |
| Stroke | Social isolation | Inverse variance weighted | 25 | 9.54e-04 (-5.75e-03 to 7.66e-03) | 0.78 | 0.16 |  |  | 96.00 |
| Stroke | Social isolation | MR-Egger | 25 | -8.60e-03 (-0.03 to 0.01) | 0.41 | 0.16 | 5.29e-04 (-5.13e-04 to 1.57e-03, 0.33) |  |  |
| Stroke | Social isolation | SIMEX adjusted MR-Egger | 25 | -8.49e-03 (-0.04 to 0.02) | 0.55 |  | 4.83e-04 (-9.72e-04 to 1.94e-03, 0.52) |  |  |
| Stroke | Social isolation | Weighted median | 25 | 2.75e-03 (-6.07e-03 to 0.01) | 0.54 |  |  |  |  |
| Stroke | Social isolation | MR-RAPS | 25 | 1.37e-03 (-5.85e-03 to 8.59e-03) | 0.71 |  |  |  |  |
| Stroke | Social isolation | PRESSO - raw | 25 | 9.54e-04 (-5.75e-03 to 7.66e-03) | 0.78 |  |  | Global = 33.68; p=0.15, Distortion = NA; p=NA |  |
| Stroke | Social isolation | PRESSO - outlier corrected | 25 (0 removed) | NA (NA to NA) |  |  |  |  |  |
| Stroke | Social isolation | MRLAP - observed | 25 | 0.02 (-0.13 to 0.18) | 0.77 |  |  |  |  |
| Stroke | Social isolation | MRLAP - corrected | 25 | 0.03 (-0.18 to 0.23) | 0.80 |  |  | Corrected vs observed estimate difference test = -0.15; p=0.88 |  |
| Stroke | Social isolation | MR-CAUSE^a^ |  | 0.00 (-0.01 to 0.01) |  |  |  | Causal vs sharing model ELPD: delta (SE) = 0.74 (0.17); p=1.00 |  |
| T2D | Loneliness | Inverse variance weighted | 226 | 1.01 (1.00 to 1.02) | 0.06 | 2.66e-21 |  |  | 99.12 |
| T2D | Loneliness | MR-Egger | 226 | 0.98 (0.95 to 1.00) | 0.05 | 8.61e-19 | 2.23e-03 (9.20e-04 to 3.54e-03, 9.90e-04) |  |  |
| T2D | Loneliness | SIMEX adjusted MR-Egger | 226 | 0.98 (0.95 to 1.00) | 0.06 |  | 2.21e-03 (8.76e-04 to 3.55e-03, 1.34e-03) |  |  |
| T2D | Loneliness | Weighted median | 226 | 0.99 (0.98 to 1.00) | 0.20 |  |  |  |  |
| T2D | Loneliness | MR-RAPS | 226 | 1.01 (1.00 to 1.02) | 0.11 |  |  |  |  |
| T2D | Loneliness | PRESSO - raw | 226 | 1.01 (1.00 to 1.02) | 0.07 |  |  | Global = 492.23; p=<2e-04, Distortion = 22.69; p=0.70 |  |
| T2D | Loneliness | PRESSO - outlier corrected | 220 (6 removed) | 1.01 (1.00 to 1.02) | 0.09 |  |  |  |  |
| T2D | Loneliness | MRLAP - observed | 302 | 1.04 (1.01 to 1.06) | 1.52e-03 |  |  |  |  |
| T2D | Loneliness | MRLAP - corrected | 302 | 1.04 (1.01 to 1.07) | 5.99e-03 |  |  | Corrected vs observed estimate difference test = 0.27; p=0.79 |  |
| T2D | Loneliness | MR-CAUSE^a^ |  | 1.02 (1.00 to 1.03) |  |  |  | Causal vs sharing model ELPD: delta (SE) = -1.02 (1.88); p=0.29 |  |
| T2D | Social isolation | Inverse variance weighted | 227 | -9.10e-05 (-2.61e-03 to 2.43e-03) | 0.94 | 1.67e-13 |  |  | 99.12 |
| T2D | Social isolation | MR-Egger | 227 | 4.71e-03 (-3.03e-04 to 9.71e-03) | 0.07 | 8.87e-13 | -3.02e-04 (-5.76e-04 to -2.89e-05, 0.03) |  |  |
| T2D | Social isolation | SIMEX adjusted MR-Egger | 227 | 4.39e-03 (-8.30e-04 to 9.62e-03) | 0.10 |  | -2.72e-04 (-5.53e-04 to 9.38e-06, 0.06) |  |  |
| T2D | Social isolation | Weighted median | 227 | 1.12e-03 (-2.45e-03 to 4.68e-03) | 0.54 |  |  |  |  |
| T2D | Social isolation | MR-RAPS | 227 | 2.35e-04 (-2.18e-03 to 2.65e-03) | 0.85 |  |  |  |  |
| T2D | Social isolation | PRESSO - raw | 227 | -9.10e-05 (-2.61e-03 to 2.43e-03) | 0.94 |  |  | Global = 420.16; p=<2e-04, Distortion = -120.84; p=0.17 |  |
| T2D | Social isolation | PRESSO - outlier corrected | 221 (6 removed) | 4.37e-04 (-1.83e-03 to 2.70e-03) | 0.71 |  |  |  |  |
| T2D | Social isolation | MRLAP - observed | 248 | 0.01 (-0.01 to 0.04) | 0.29 |  |  |  |  |
| T2D | Social isolation | MRLAP - corrected | 248 | 0.01 (-0.02 to 0.04) | 0.37 |  |  | Corrected vs observed estimate difference test = 0.12; p=0.90 |  |
| T2D | Social isolation | MR-CAUSE^a^ |  | 0.00 (0.00 to 0.00) |  |  |  | Causal vs sharing model ELPD: delta (SE) = 0.79 (0.43); p=0.97 |  |
| Suicide attempt^b^ | Loneliness | Inverse variance weighted | 14 | 1.00 (1.00 to 1.00) | 0.48 | 3.17e-04 |  |  | 100.00 |
| Suicide attempt | Loneliness | MR-Egger | 14 | 1.08 (1.04 to 1.12) | 9.22e-04 | 0.25 | -0.08 (-0.11 to -0.04, 1.02e-03) |  |  |
| Suicide attempt | Loneliness | SIMEX adjusted MR-Egger | 14 | 1.10 (0.95 to 1.28) | 0.22 |  | -1.27e-03 (-0.01 to 8.09e-03, 0.80) |  |  |
| Suicide attempt | Loneliness | Weighted median | 14 | 1.00 (1.00 to 1.00) | 0.56 |  |  |  |  |
| Suicide attempt | Loneliness | MR-RAPS | 14 | 1.08 (1.04 to 1.12) | 3.88e-05 |  |  |  |  |
| Suicide attempt | Loneliness | PRESSO - raw | 14 | 1.08 (1.05 to 1.12) | 4.61e-04 |  |  | Global = 17.43; p=0.37, Distortion = NA; p=NA |  |
| Suicide attempt | Loneliness | PRESSO - outlier corrected | 14 (0 removed) | NA (NA to NA) |  |  |  |  |  |
| Suicide attempt | Loneliness | MRLAP - observed | 14 | 1.23 (1.09 to 1.38) | 6.38e-04 |  |  |  |  |
| Suicide attempt | Loneliness | MRLAP - corrected | 14 | 1.43 (1.15 to 1.77) | 1.29e-03 |  |  | Corrected vs observed estimate difference test = -3.01; p=2.60e-03 |  |
| Suicide attempt | Loneliness | MR-CAUSE^a^ |  | 1.04 (1.00 to 1.08) |  |  |  | Causal vs sharing model ELPD: delta (SE) = -1.61 (1.59); p=0.16 |  |
| Suicide attempt^b^ | Social isolation | Inverse variance weighted | 14 | 1.55e-05 (-6.71e-04 to 7.02e-04) | 0.96 | 4.86e-04 |  |  | 100.00 |
| Suicide attempt | Social isolation | MR-Egger | 14 | 0.01 (7.56e-04 to 0.02) | 0.06 | 8.57e-03 | -0.01 (-0.02 to -7.74e-04, 0.06) |  |  |
| Suicide attempt | Social isolation | SIMEX adjusted MR-Egger | 14 | 0.01 (7.64e-04 to 0.02) | 0.06 |  | -0.01 (-0.02 to -7.84e-04, 0.06) |  |  |
| Suicide attempt | Social isolation | Weighted median | 14 | 1.04e-05 (-6.13e-04 to 6.34e-04) | 0.97 |  |  |  |  |
| Suicide attempt | Social isolation | MR-RAPS | 14 | 1.10e-04 (-4.30e-04 to 6.50e-04) | 0.69 |  |  |  |  |
| Suicide attempt | Social isolation | PRESSO - raw | 14 | 1.55e-05 (-6.71e-04 to 7.02e-04) | 0.97 |  |  | Global = 43.96; p=5.00e-04, Distortion = 134.99; p=0.09 |  |
| Suicide attempt | Social isolation | PRESSO - outlier corrected | 12 (2 removed) | -4.44e-05 (-5.12e-04 to 4.24e-04) | 0.86 |  |  |  |  |
| Suicide attempt | Social isolation | MRLAP - observed | 11 | 0.14 (-1.42e-03 to 0.28) | 0.05 |  |  |  |  |
| Suicide attempt | Social isolation | MRLAP - corrected | 11 | 0.26 (-0.02 to 0.54) | 0.07 |  |  | Corrected vs observed estimate difference test = -1.64; p=0.10 |  |
| Suicide attempt | Social isolation | MR-CAUSE^a^ |  | 0.01 (0.00 to 0.01) |  |  |  | Causal vs sharing model ELPD: delta (SE) = -0.86 (1.35); p=0.26 |  |
| Depression | Loneliness | Inverse variance weighted | 29 | 1.20 (1.14 to 1.27) | 1.96e-11 | 1.20e-04 |  |  | 100.00 |
| Depression | Loneliness | MR-Egger | 29 | 1.01 (0.86 to 1.18) | 0.90 | 1.68e-03 | 6.12e-03 (8.82e-04 to 0.01, 0.03) |  |  |
| Depression | Loneliness | SIMEX adjusted MR-Egger | 29 | 1.02 (0.84 to 1.25) | 0.84 |  | 5.76e-03 (-9.12e-04 to 0.01, 0.10) |  |  |
| Depression | Loneliness | Weighted median | 29 | 1.12 (1.05 to 1.19) | 3.20e-04 |  |  |  |  |
| Depression | Loneliness | MR-RAPS | 29 | 1.21 (1.14 to 1.28) | 4.17e-11 |  |  |  |  |
| Depression | Loneliness | PRESSO - raw | 29 | 1.20 (1.14 to 1.27) | 2.78e-07 |  |  | Global = 68.96; p=8.00e-03, Distortion = 5.49; p=0.71 |  |
| Depression | Loneliness | PRESSO - outlier corrected | 28 (1 removed) | 1.19 (1.13 to 1.25) | 1.82e-07 |  |  |  |  |
| Depression | Loneliness | MRLAP - observed | 2 | 1.26 (0.88 to 1.80) | 0.20 |  |  |  |  |
| Depression | Loneliness | MRLAP - corrected | 2 | 1.54 (0.79 to 3.01) | 0.20 |  |  | Corrected vs observed estimate difference test = -1.26; p=0.21 |  |
| Depression | Loneliness | MR-CAUSE^a^ |  | 1.05 (1.01 to 1.09) |  |  |  | Causal vs sharing model ELPD: delta (SE) = -2.34 (1.74); p=0.09 |  |
| Depression | Social isolation | Inverse variance weighted | 29 | 0.02 (4.47e-03 to 0.03) | 6.66e-03 | 2.82e-04 |  |  | 100.00 |
| Depression | Social isolation | MR-Egger | 29 | 0.03 (-9.50e-03 to 0.06) | 0.16 | 2.38e-04 | -4.00e-04 (-1.63e-03 to 8.35e-04, 0.53) |  |  |
| Depression | Social isolation | SIMEX adjusted MR-Egger | 29 | 0.04 (-0.02 to 0.09) | 0.21 |  | -7.83e-04 (-2.57e-03 to 1.00e-03, 0.40) |  |  |
| Depression | Social isolation | Weighted median | 29 | 7.15e-03 (-5.77e-03 to 0.02) | 0.28 |  |  |  |  |
| Depression | Social isolation | MR-RAPS | 29 | 0.01 (-9.70e-04 to 0.02) | 0.07 |  |  |  |  |
| Depression | Social isolation | PRESSO - raw | 29 | 0.02 (4.47e-03 to 0.03) | 0.01 |  |  | Global = 67.67; p=<5e-04, Distortion = 50.49; p=0.12 |  |
| Depression | Social isolation | PRESSO - outlier corrected | 28 (1 removed) | 0.01 (6.85e-04 to 0.02) | 0.05 |  |  |  |  |
| Depression | Social isolation | MR-CAUSE^a^ |  | 0.00 (0.00 to 0.01) |  |  |  | Causal vs sharing model ELPD: delta (SE) = 0.40 (0.82); p=0.69 |  |
| Anxiety^c^ | Loneliness | Inverse variance weighted | 18 | 1.00 (0.99 to 1.01) | 0.88 | 0.02 |  |  | 100.00 |
| Anxiety | Loneliness | MR-Egger | 18 | 1.01 (0.97 to 1.04) | 0.78 | 0.01 | -8.38e-04 (-7.57e-03 to 5.89e-03, 0.81) |  |  |
| Anxiety | Loneliness | SIMEX adjusted MR-Egger | 18 | 1.01 (0.96 to 1.06) | 0.77 |  | -1.13e-03 (-9.55e-03 to 7.28e-03, 0.79) |  |  |
| Anxiety | Loneliness | Weighted median | 18 | 1.01 (0.99 to 1.02) | 0.36 |  |  |  |  |
| Anxiety | Loneliness | MR-RAPS | 18 | 1.00 (0.99 to 1.02) | 0.74 |  |  |  |  |
| Anxiety | Loneliness | PRESSO - raw | 18 | 1.00 (0.99 to 1.01) | 0.88 |  |  | Global = 35.83; p=0.02, Distortion = NA; p=NA |  |
| Anxiety | Loneliness | PRESSO - outlier corrected | 17 (1 removed) | NA (NA to NA) |  |  |  |  |  |
| Anxiety | Loneliness | MRLAP - observed | 15 | 1.00 (0.98 to 1.03) | 0.68 |  |  |  |  |
| Anxiety | Loneliness | MRLAP - corrected | 15 | 1.03 (0.58 to 1.84) | 0.92 |  |  | Corrected vs observed estimate difference test = -0.08; p=0.94 |  |
| Anxiety | Loneliness | MR-CAUSE^a^ |  | 0.97 (0.94 to 1.01) |  |  |  | Causal vs sharing model ELPD: delta (SE) = -1.10 (1.06); p=0.15 |  |
| Anxiety^c^ | Social isolation | Inverse variance weighted | 20 | -9.07e-04 (-2.84e-03 to 1.03e-03) | 0.36 | 0.28 |  |  | 100.00 |
| Anxiety | Social isolation | MR-Egger | 20 | -1.29e-03 (-6.41e-03 to 3.83e-03) | 0.63 | 0.23 | 8.16e-05 (-9.20e-04 to 1.08e-03, 0.87) |  |  |
| Anxiety | Social isolation | SIMEX adjusted MR-Egger | 20 | -3.03e-03 (-8.20e-03 to 2.14e-03) | 0.27 |  | 4.19e-04 (-8.10e-04 to 1.65e-03, 0.51) |  |  |
| Anxiety | Social isolation | Weighted median | 20 | -1.44e-04 (-2.88e-03 to 2.59e-03) | 0.92 |  |  |  |  |
| Anxiety | Social isolation | MR-RAPS | 20 | -1.06e-03 (-3.25e-03 to 1.13e-03) | 0.34 |  |  |  |  |
| Anxiety | Social isolation | PRESSO - raw | 20 | -9.07e-04 (-2.84e-03 to 1.03e-03) | 0.37 |  |  | Global = 24.28; p=0.31, Distortion = NA; p=NA |  |
| Anxiety | Social isolation | PRESSO - outlier corrected | 20 (0 removed) | NA (NA to NA) |  |  |  |  |  |
| Anxiety | Social isolation | MR-CAUSE^a^ |  | 0.00 (-0.02 to 0.01) |  |  |  | Causal vs sharing model ELPD: delta (SE) = 0.44 (0.21); p=0.98 |  |
| Wellbeing spectrum | Loneliness | Inverse variance weighted | 132 | 0.27 (0.24 to 0.30) | 4.41e-136 | 1.17e-03 |  |  | 69.70 |
| Wellbeing spectrum | Loneliness | MR-Egger | 132 | 0.33 (0.20 to 0.55) | 4.07e-05 | 1.09e-03 | -1.26e-03 (-4.65e-03 to 2.13e-03, 0.47) |  |  |
| Wellbeing spectrum | Loneliness | SIMEX adjusted MR-Egger | 132 | 0.25 (0.13 to 0.47) | 2.41e-05 |  | 5.60e-04 (-3.66e-03 to 4.78e-03, 0.80) |  |  |
| Wellbeing spectrum | Loneliness | Weighted median | 132 | 0.30 (0.27 to 0.35) | 3.57e-67 |  |  |  |  |
| Wellbeing spectrum | Loneliness | MR-RAPS | 132 | 0.26 (0.23 to 0.29) | 0.00 |  |  |  |  |
| Wellbeing spectrum | Loneliness | PRESSO - raw | 132 | 0.27 (0.24 to 0.30) | 2.21e-51 |  |  | Global = 188.63; p=0.04, Distortion = NA; p=NA |  |
| Wellbeing spectrum | Loneliness | PRESSO - outlier corrected | 131 (1 removed) | NA (NA to NA) |  |  |  |  |  |
| Wellbeing spectrum | Loneliness | MRLAP - observed | 120 | 0.42 (0.40 to 0.45) | 6.84e-139 |  |  |  |  |
| Wellbeing spectrum | Loneliness | MRLAP - corrected | 120 | 0.41 (0.37 to 0.44) | 2.01e-92 |  |  | Corrected vs observed estimate difference test = 3.28; p=1.02e-03 |  |
| Wellbeing spectrum | Loneliness | MR-CAUSE^a^ |  | 0.35 (0.32 to 0.39) |  |  |  | Causal vs sharing model ELPD: delta (SE) = -8.40 (1.16); p=2.74e-13 |  |
| Wellbeing spectrum | Social isolation | Inverse variance weighted | 133 | -0.09 (-0.12 to -0.06) | 1.83e-08 | 1.21e-23 |  |  | 86.47 |
| Wellbeing spectrum | Social isolation | MR-Egger | 133 | -0.06 (-0.22 to 0.10) | 0.48 | 8.46e-24 | -2.24e-04 (-1.27e-03 to 8.22e-04, 0.68) |  |  |
| Wellbeing spectrum | Social isolation | SIMEX adjusted MR-Egger | 133 | -0.07 (-0.24 to 0.10) | 0.44 |  | -1.83e-04 (-1.36e-03 to 9.95e-04, 0.76) |  |  |
| Wellbeing spectrum | Social isolation | Weighted median | 133 | -0.09 (-0.12 to -0.06) | 3.49e-08 |  |  |  |  |
| Wellbeing spectrum | Social isolation | MR-RAPS | 133 | -0.10 (-0.13 to -0.07) | 1.01e-09 |  |  |  |  |
| Wellbeing spectrum | Social isolation | PRESSO - raw | 133 | -0.09 (-0.12 to -0.06) | 1.05e-07 |  |  | Global = 369.87; p=<0.0003, Distortion = 1.00; p=0.95 |  |
| Wellbeing spectrum | Social isolation | PRESSO - outlier corrected | 129 (4 removed) | -0.09 (-0.12 to -0.06) | 2.28e-09 |  |  |  |  |
| Wellbeing spectrum | Social isolation | MRLAP - observed | 11 | 0.74 (0.52 to 1.05) | 0.09 |  |  |  |  |
| Wellbeing spectrum | Social isolation | MRLAP - corrected | 11 | 0.69 (0.37 to 1.29) | 0.25 |  |  | Corrected vs observed estimate difference test = 0.49; p=0.62 |  |
| Wellbeing spectrum | Social isolation | MR-CAUSE^a^ |  | -0.07 (-0.11 to -0.03) |  |  |  | Causal vs sharing model ELPD: delta (SE) = -0.73 (2.09); p=0.36 |  |
| Positive affect | Loneliness | Inverse variance weighted | 92 | 0.37 (0.33 to 0.41) | 2.13e-66 | 2.02e-09 |  |  | 73.91 |
| Positive affect | Loneliness | MR-Egger | 92 | 0.78 (0.56 to 1.09) | 0.14 | 1.81e-05 | -7.08e-03 (-0.01 to -4.09e-03, 1.15e-05) |  |  |
| Positive affect | Loneliness | SIMEX adjusted MR-Egger | 92 | 0.63 (0.38 to 1.03) | 0.07 |  | -5.28e-03 (-9.59e-03 to -9.60e-04, 0.02) |  |  |
| Positive affect | Loneliness | Weighted median | 92 | 0.43 (0.37 to 0.50) | 9.13e-32 |  |  |  |  |
| Positive affect | Loneliness | MR-RAPS | 92 | 0.34 (0.30 to 0.38) | 0.00 |  |  |  |  |
| Positive affect | Loneliness | PRESSO - raw | 92 | 0.37 (0.33 to 0.41) | 2.29e-30 |  |  | Global = 198.71; p=4.50e-03, Distortion = 2.70; p=0.61 |  |
| Positive affect | Loneliness | PRESSO - outlier corrected | 91 (1 removed) | 0.36 (0.32 to 0.40) | 9.97e-32 |  |  |  |  |
| Positive affect | Loneliness | MRLAP - observed | 9 | 0.73 (0.65 to 0.81) | 1.13e-08 |  |  |  |  |
| Positive affect | Loneliness | MRLAP - corrected | 9 | 0.65 (0.55 to 0.78) | 1.37e-06 |  |  | Corrected vs observed estimate difference test = 3.19; p=1.42e-03 |  |
| Positive affect | Loneliness | MR-CAUSE^a^ |  | 0.73 (0.65 to 0.81) |  |  |  | Causal vs sharing model ELPD: delta (SE) = -5.36 (1.73); p=9.50e-04 |  |
| Positive affect | Social isolation | Inverse variance weighted | 93 | -0.09 (-0.11 to -0.06) | 9.96e-11 | 3.49e-12 |  |  | 93.55 |
| Positive affect | Social isolation | MR-Egger | 93 | -0.09 (-0.18 to -2.89e-03) | 0.05 | 2.24e-12 | 2.33e-05 (-7.55e-04 to 8.02e-04, 0.95) |  |  |
| Positive affect | Social isolation | SIMEX adjusted MR-Egger | 93 | -0.10 (-0.23 to 0.02) | 0.10 |  | 1.33e-04 (-9.95e-04 to 1.26e-03, 0.82) |  |  |
| Positive affect | Social isolation | Weighted median | 93 | -0.05 (-0.08 to -0.02) | 1.37e-03 |  |  |  |  |
| Positive affect | Social isolation | MR-RAPS | 93 | -0.08 (-0.11 to -0.06) | 6.23e-10 |  |  |  |  |
| Positive affect | Social isolation | PRESSO - raw | 93 | -0.09 (-0.11 to -0.06) | 4.73e-09 |  |  | Global = 224.91; p=<5e-04, Distortion = -18.18; p=0.19 |  |
| Positive affect | Social isolation | PRESSO - outlier corrected | 91 (2 removed) | -0.07 (-0.10 to -0.05) | 5.72e-08 |  |  |  |  |
| Positive affect | Social isolation | MRLAP - observed | 98 | -0.28 (-0.36 to -0.20) | 1.92e-12 |  |  |  |  |
| Positive affect | Social isolation | MRLAP - corrected | 98 | -0.35 (-0.45 to -0.25) | 3.01e-11 |  |  | Corrected vs observed estimate difference test = 5.33; p=9.77e-08 |  |
| Positive affect | Social isolation | MR-CAUSE^a^ |  | -0.02 (-0.05 to 0.00) |  |  |  | Causal vs sharing model ELPD: delta (SE) = -0.87 (1.41); p=0.27 |  |
| Life satisfaction | Loneliness | Inverse variance weighted | 73 | 0.42 (0.38 to 0.47) | 5.79e-61 | 8.07e-06 |  |  | 83.56 |
| Life satisfaction | Loneliness | MR-Egger | 73 | 0.79 (0.64 to 0.99) | 0.04 | 0.06 | -6.97e-03 (-9.24e-03 to -4.70e-03, 7.33e-08) |  |  |
| Life satisfaction | Loneliness | SIMEX adjusted MR-Egger | 73 | 0.61 (0.43 to 0.87) | 8.28e-03 |  | -4.38e-03 (-7.91e-03 to -8.57e-04, 0.02) |  |  |
| Life satisfaction | Loneliness | Weighted median | 73 | 0.55 (0.46 to 0.66) | 6.01e-11 |  |  |  |  |
| Life satisfaction | Loneliness | MR-RAPS | 73 | 0.36 (0.29 to 0.45) | 0.00 |  |  |  |  |
| Life satisfaction | Loneliness | PRESSO - raw | 73 | 0.42 (0.38 to 0.47) | 3.97e-26 |  |  | Global = 139.97; p=0.95, Distortion = NA; p=NA |  |
| Life satisfaction | Loneliness | PRESSO - outlier corrected | 73 (0 removed) | NA (NA to NA) |  |  |  |  |  |
| Life satisfaction | Loneliness | MRLAP - observed | 75 | 0.58 (0.55 to 0.62) | 5.79e-67 |  |  |  |  |
| Life satisfaction | Loneliness | MRLAP - corrected | 75 | 0.49 (0.45 to 0.54) | 3.67e-57 |  |  | Corrected vs observed estimate difference test = 7.72; p=1.12e-14 |  |
| Life satisfaction | Loneliness | MR-CAUSE^a^ |  | 0.88 (0.73 to 1.06) |  |  |  | Causal vs sharing model ELPD: delta (SE) = -0.21 (1.02); p=0.42 |  |
| Life satisfaction | Social isolation | Inverse variance weighted | 73 | -0.06 (-0.09 to -0.04) | 7.61e-07 | 1.75e-10 |  |  | 94.52 |
| Life satisfaction | Social isolation | MR-Egger | 73 | -0.05 (-0.12 to 0.01) | 0.12 | 1.19e-10 | -1.15e-04 (-8.19e-04 to 5.89e-04, 0.75) |  |  |
| Life satisfaction | Social isolation | SIMEX adjusted MR-Egger | 73 | -0.10 (-0.22 to 0.02) | 0.11 |  | 3.56e-04 (-8.13e-04 to 1.53e-03, 0.55) |  |  |
| Life satisfaction | Social isolation | Weighted median | 73 | -0.04 (-0.07 to -0.01) | 6.61e-03 |  |  |  |  |
| Life satisfaction | Social isolation | MR-RAPS | 73 | -0.08 (-0.12 to -0.04) | 2.06e-05 |  |  |  |  |
| Life satisfaction | Social isolation | PRESSO - raw | 73 | -0.06 (-0.09 to -0.04) | 4.81e-06 |  |  | Global = 181.40; p=<5e-04, Distortion = -18.75; p=0.29 |  |
| Life satisfaction | Social isolation | PRESSO - outlier corrected | 72 (1 removed) | -0.05 (-0.08 to -0.03) | 3.96e-05 |  |  |  |  |
| Life satisfaction | Social isolation | MR-CAUSE^a^ |  | 0.00 (-0.04 to 0.04) |  |  |  | Causal vs sharing model ELPD: delta (SE) = 0.66 (0.11); p=1.00 |  |

*OR=odds ratio, CI=confidence intervals, MR=Mendelian Randomisation, SIMEX=* *simulation extrapolation, MR-RAPS=* *MR using robust adjusted profile score, PRESSO=Pleiotropy RESidual Sum and Outlier, CAD=coronary artery disease, HF=heart failure, T2D=type 2 diabetes, SBP=systolic blood pressure, ELPD=expected log pointwise posterior density, SE=standard error*

*^a^ MR-CAUSE is a Bayesian method so the values reported are the posterior median as the estimate and the posterior credible interval as the CI.*

*^b^Where suicide attempt was the exposure a p-value threshold of 1e-06 was used*

*^c^Where anxiety was the exposure a p-value threshold of 1e-05 was used*

### Table S16. Multivariable Mendelian Randomisation analysis results with loneliness and social isolation as exposures

| **Exposure** | **Outcome** | **Method** | **OR or mean difference (95% CI)** | **p-value** | **Conditional F statistic** | **Additional tests for MR approaches** | **Pleiotropy test**  **(p-value)** |
| --- | --- | --- | --- | --- | --- | --- | --- |
| Loneliness | CAD | Inverse variance weighted | 1.21 (0.93 to 1.57) | 0.16 | 17.50 |  | 62.91 (1.43e-05) |
| Social isolation | CAD | Inverse variance weighted | 0.66 (0.20 to 2.15) | 0.50 | 16.90 |  |  |
| Loneliness | CAD | Egger - orientation Lone | 1.15 (0.63 to 2.09) | 0.66 |  |  |  |
| Social isolation | CAD | Egger - orientation SI | 0.81 (0.09 to 7.27) | 0.86 |  |  |  |
| Loneliness | CAD | PRESSO - raw | 1.21 (0.93 to 1.57) | 0.16 |  | Global = 74.50; p=5.00e-04, Distortion = -2.47; p=1.12 |  |
| Social isolation | CAD | PRESSO - raw | 0.66 (0.20 to 2.15) | 0.50 |  | Global = 74.50; p=5.00e-04, Distortion = -525.28; p=1.12 |  |
| Loneliness | CAD | PRESSO - outlier corrected | 1.22 (0.96 to 1.55) | 0.12 |  |  |  |
| Social isolation | CAD | PRESSO - outlier corrected | 0.94 (0.30 to 2.91) | 0.91 |  |  |  |
| Loneliness | Systolic blood pressure | Inverse variance weighted | -3.33 (-6.95 to 0.29) | 0.09 | 16.32 |  | 243.25 (9.29e-40) |
| Social isolation | Systolic blood pressure | Inverse variance weighted | 10.36 (-6.19 to 26.91) | 0.23 | 15.55 |  |  |
| Loneliness | Systolic blood pressure | Egger - orientation Lone | -2.66 (-10.99 to 5.68) | 0.54 |  |  |  |
| Social isolation | Systolic blood pressure | Egger - orientation SI | -14.70 (-44.33 to 14.92) | 0.34 |  |  |  |
| Loneliness | Systolic blood pressure | PRESSO - raw | -3.33 (-6.95 to 0.29) | 0.09 |  | Global = 326.95; p=<5e-04, Distortion = -51.02; p=1.18 |  |
| Social isolation | Systolic blood pressure | PRESSO - raw | 10.36 (-6.19 to 26.91) | 0.23 |  | Global = 326.95; p=<5e-04, Distortion = 197.95; p=1.18 |  |
| Loneliness | Systolic blood pressure | PRESSO - outlier corrected | -2.20 (-5.91 to 1.51) | 0.26 |  |  |  |
| Social isolation | Systolic blood pressure | PRESSO - outlier corrected | 3.48 (-14.29 to 21.24) | 0.71 |  |  |  |
| Loneliness | Heart failure | Inverse variance weighted | 1.02 (0.72 to 1.44) | 0.93 | 17.50 |  | 40.73 (0.01) |
| Social isolation | Heart failure | Inverse variance weighted | 0.70 (0.15 to 3.40) | 0.67 | 16.90 |  |  |
| Loneliness | Heart failure | Egger - orientation Lone | 0.81 (0.36 to 1.79) | 0.60 |  |  |  |
| Social isolation | Heart failure | Egger - orientation SI | 5.56 (0.34 to 91.85) | 0.24 |  |  |  |
| Loneliness | Heart failure | PRESSO - raw | 1.02 (0.72 to 1.44) | 0.93 |  | Global = 49.88; p=0.01, Distortion = NA; p=NA |  |
| Social isolation | Heart failure | PRESSO - raw | 0.70 (0.15 to 3.40) | 0.67 |  | Global = 49.88; p=0.01, Distortion = NA; p=NA |  |
| Loneliness | Heart failure | PRESSO - outlier corrected | NA (NA to NA) |  |  |  |  |
| Social isolation | Heart failure | PRESSO - outlier corrected | NA (NA to NA) |  |  |  |  |
| Loneliness | Stroke | Inverse variance weighted | 0.90 (0.62 to 1.32) | 0.60 | 17.50 |  | 36.27 (0.04) |
| Social isolation | Stroke | Inverse variance weighted | 1.57 (0.29 to 8.54) | 0.61 | 16.90 |  |  |
| Loneliness | Stroke | Egger - orientation Lone | 0.53 (0.23 to 1.22) | 0.15 |  |  |  |
| Social isolation | Stroke | Egger - orientation SI | 63.28 (4.45 to 900.26) | 5.53e-03 |  |  |  |
| Loneliness | Stroke | PRESSO - raw | 0.90 (0.62 to 1.32) | 0.60 |  | Global = 42.49; p=0.06, Distortion = NA; p=NA |  |
| Social isolation | Stroke | PRESSO - raw | 1.57 (0.29 to 8.54) | 0.61 |  | Global = 42.49; p=0.06, Distortion = NA; p=NA |  |
| Loneliness | Stroke | PRESSO - outlier corrected | NA (NA to NA) |  |  |  |  |
| Social isolation | Stroke | PRESSO - outlier corrected | NA (NA to NA) |  |  |  |  |
| Loneliness | T2D | Inverse variance weighted | 1.22 (0.79 to 1.88) | 0.37 | 17.50 |  | 187.47 (9.37e-28) |
| Social isolation | T2D | Inverse variance weighted | 0.62 (0.09 to 4.43) | 0.64 | 16.90 |  |  |
| Loneliness | T2D | Egger - orientation Lone | 1.73 (0.65 to 4.61) | 0.28 |  |  |  |
| Social isolation | T2D | Egger - orientation SI | 3.14 (0.09 to 107.14) | 0.53 |  |  |  |
| Loneliness | T2D | PRESSO - raw | 1.22 (0.79 to 1.88) | 0.37 |  | Global = 219.54; p=<5e-04, Distortion = 38.20; p=1.42 |  |
| Social isolation | T2D | PRESSO - raw | 0.62 (0.09 to 4.43) | 0.64 |  | Global = 219.54; p=<5e-04, Distortion = -118.65; p=1.42 |  |
| Loneliness | T2D | PRESSO - outlier corrected | 1.16 (0.85 to 1.56) | 0.36 |  |  |  |
| Social isolation | T2D | PRESSO - outlier corrected | 0.81 (0.20 to 3.31) | 0.77 |  |  |  |
| Loneliness | Suicide attempt | Inverse variance weighted | 1.96 (1.07 to 3.60) | 0.04 | 17.50 |  | 83.85 (7.49e-09) |
| Social isolation | Suicide attempt | Inverse variance weighted | 2.26 (0.14 to 35.51) | 0.57 | 16.90 |  |  |
| Loneliness | Suicide attempt | Egger - orientation Lone | 1.76 (0.42 to 7.28) | 0.44 |  |  |  |
| Social isolation | Suicide attempt | Egger - orientation SI | 2.69 (0.02 to 413.49) | 0.70 |  |  |  |
| Loneliness | Suicide attempt | PRESSO - raw | 1.96 (1.07 to 3.60) | 0.04 |  | Global = 105.22; p=<5e-04, Distortion = -4.05; p=1.00 |  |
| Social isolation | Suicide attempt | PRESSO - raw | 2.26 (0.14 to 35.51) | 0.57 |  | Global = 105.22; p=<5e-04, Distortion = 744.25; p=1.00 |  |
| Loneliness | Suicide attempt | PRESSO - outlier corrected | 2.02 (1.20 to 3.38) | 0.01 |  |  |  |
| Social isolation | Suicide attempt | PRESSO - outlier corrected | 0.88 (0.08 to 9.79) | 0.92 |  |  |  |
| Loneliness | Depression | Inverse variance weighted | 2.09 (1.28 to 3.41) | 7.17e-03 | 17.50 |  | 64.45 (8.49e-06) |
| Social isolation | Depression | Inverse variance weighted | 0.51 (0.06 to 4.63) | 0.55 | 16.90 |  |  |
| Loneliness | Depression | Egger - orientation Lone | 2.12 (0.68 to 6.58) | 0.21 |  |  |  |
| Social isolation | Depression | Egger - orientation SI | 0.03 (6.68e-04 to 1.48) | 0.09 |  |  |  |
| Loneliness | Depression | PRESSO - raw | 2.09 (1.28 to 3.41) | 7.17e-03 |  | Global = 79.99; p=<5e-04, Distortion = 3.16; p=1.00 |  |
| Social isolation | Depression | PRESSO - raw | 0.51 (0.06 to 4.63) | 0.55 |  | Global = 79.99; p=<5e-04, Distortion = -536.56; p=1.00 |  |
| Loneliness | Depression | PRESSO - outlier corrected | 2.04 (1.32 to 3.17) | 4.12e-03 |  |  |  |
| Social isolation | Depression | PRESSO - outlier corrected | 1.17 (0.15 to 9.30) | 0.88 |  |  |  |
| Loneliness | Anxiety | Inverse variance weighted | 2.52 (0.98 to 6.50) | 0.07 | 16.61 |  | 22.54 (0.43) |
| Social isolation | Anxiety | Inverse variance weighted | 0.10 (1.62e-03 to 6.39) | 0.29 | 16.69 |  |  |
| Loneliness | Anxiety | Egger - orientation Lone | 0.94 (0.11 to 7.64) | 0.95 |  |  |  |
| Social isolation | Anxiety | Egger - orientation SI | 0.19 (3.89e-05 to 921.42) | 0.70 |  |  |  |
| Loneliness | Anxiety | PRESSO - raw | 2.52 (0.98 to 6.50) | 0.07 |  | Global = 26.10; p=0.55, Distortion = NA; p=NA |  |
| Social isolation | Anxiety | PRESSO - raw | 0.10 (1.62e-03 to 6.39) | 0.29 |  | Global = 26.10; p=0.55, Distortion = NA; p=NA |  |
| Loneliness | Anxiety | PRESSO - outlier corrected | NA (NA to NA) |  |  |  |  |
| Social isolation | Anxiety | PRESSO - outlier corrected | NA (NA to NA) |  |  |  |  |
| Loneliness | Wellbeing spectrum | Inverse variance weighted | -0.26 (-0.35 to -0.18) | 6.71e-06 | 16.61 |  | 68.45 (8.24e-08) |
| Social isolation | Wellbeing spectrum | Inverse variance weighted | 0.02 (-0.32 to 0.37) | 0.89 | 18.25 |  |  |
| Loneliness | Wellbeing spectrum | Egger - orientation Lone | -0.32 (-0.53 to -0.10) | 9.07e-03 |  |  |  |
| Social isolation | Wellbeing spectrum | Egger - orientation SI | 0.10 (-0.55 to 0.74) | 0.77 |  |  |  |
| Loneliness | Wellbeing spectrum | PRESSO - raw | -0.26 (-0.35 to -0.18) | 6.71e-06 |  | Global = 152.60; p=<5e-04, Distortion = 13.15; p=1.40 |  |
| Social isolation | Wellbeing spectrum | PRESSO - raw | 0.02 (-0.32 to 0.37) | 0.89 |  | Global = 152.60; p=<5e-04, Distortion = -86.13; p=1.40 |  |
| Loneliness | Wellbeing spectrum | PRESSO - outlier corrected | -0.30 (-0.37 to -0.24) | 3.41e-08 |  |  |  |
| Social isolation | Wellbeing spectrum | PRESSO - outlier corrected | 0.17 (-0.11 to 0.45) | 0.24 |  |  |  |
| Loneliness | Positive affect | Inverse variance weighted | -0.31 (-0.42 to -0.19) | 3.46e-05 | 16.61 |  | 30.72 (0.03) |
| Social isolation | Positive affect | Inverse variance weighted | -0.30 (-0.77 to 0.16) | 0.21 | 18.25 |  |  |
| Loneliness | Positive affect | Egger - orientation Lone | -0.26 (-0.54 to 0.03) | 0.09 |  |  |  |
| Social isolation | Positive affect | Egger - orientation SI | -0.48 (-1.33 to 0.38) | 0.29 |  |  |  |
| Loneliness | Positive affect | PRESSO - raw | -0.31 (-0.42 to -0.19) | 3.46e-05 |  | Global = 46.13; p=0.04, Distortion = -3.12; p=0.89 |  |
| Social isolation | Positive affect | PRESSO - raw | -0.30 (-0.77 to 0.16) | 0.21 |  | Global = 46.13; p=0.04, Distortion = -365.66; p=0.89 |  |
| Loneliness | Positive affect | PRESSO - outlier corrected | -0.30 (-0.36 to -0.23) | 4.40e-08 |  |  |  |
| Social isolation | Positive affect | PRESSO - outlier corrected | -0.07 (-0.34 to 0.21) | 0.65 |  |  |  |
| Loneliness | Life satisfaction | Inverse variance weighted | -0.44 (-0.67 to -0.21) | 1.29e-03 | 16.61 |  | 29.09 (0.05) |
| Social isolation | Life satisfaction | Inverse variance weighted | 0.14 (-0.82 to 1.10) | 0.78 | 18.25 |  |  |
| Loneliness | Life satisfaction | Egger - orientation Lone | -0.49 (-1.07 to 0.10) | 0.12 |  |  |  |
| Social isolation | Life satisfaction | Egger - orientation SI | 1.28 (-0.40 to 2.96) | 0.15 |  |  |  |
| Loneliness | Life satisfaction | PRESSO - raw | -0.44 (-0.67 to -0.21) | 1.29e-03 |  | Global = 37.79; p=0.07, Distortion = NA; p=NA |  |
| Social isolation | Life satisfaction | PRESSO - raw | 0.14 (-0.82 to 1.10) | 0.78 |  | Global = 37.79; p=0.07, Distortion = NA; p=NA |  |
| Loneliness | Life satisfaction | PRESSO - outlier corrected | NA (NA to NA) |  |  |  |  |
| Social isolation | Life satisfaction | PRESSO - outlier corrected | NA (NA to NA) |  |  |  |  |

*OR=odds ratio, CI=confidence intervals, MR=Mendelian Randomisation, SI=social isolation, PRESSO=Pleiotropy RESidual Sum and Outlier, CAD=Coronary artery disease, HF=heart failure, T2D=type 2 diabetes, SBP=systolic blood pressure*

### Figure S1. (a) Manhattan plot and (b) Quantile-Quantile plot from social isolation GWAS

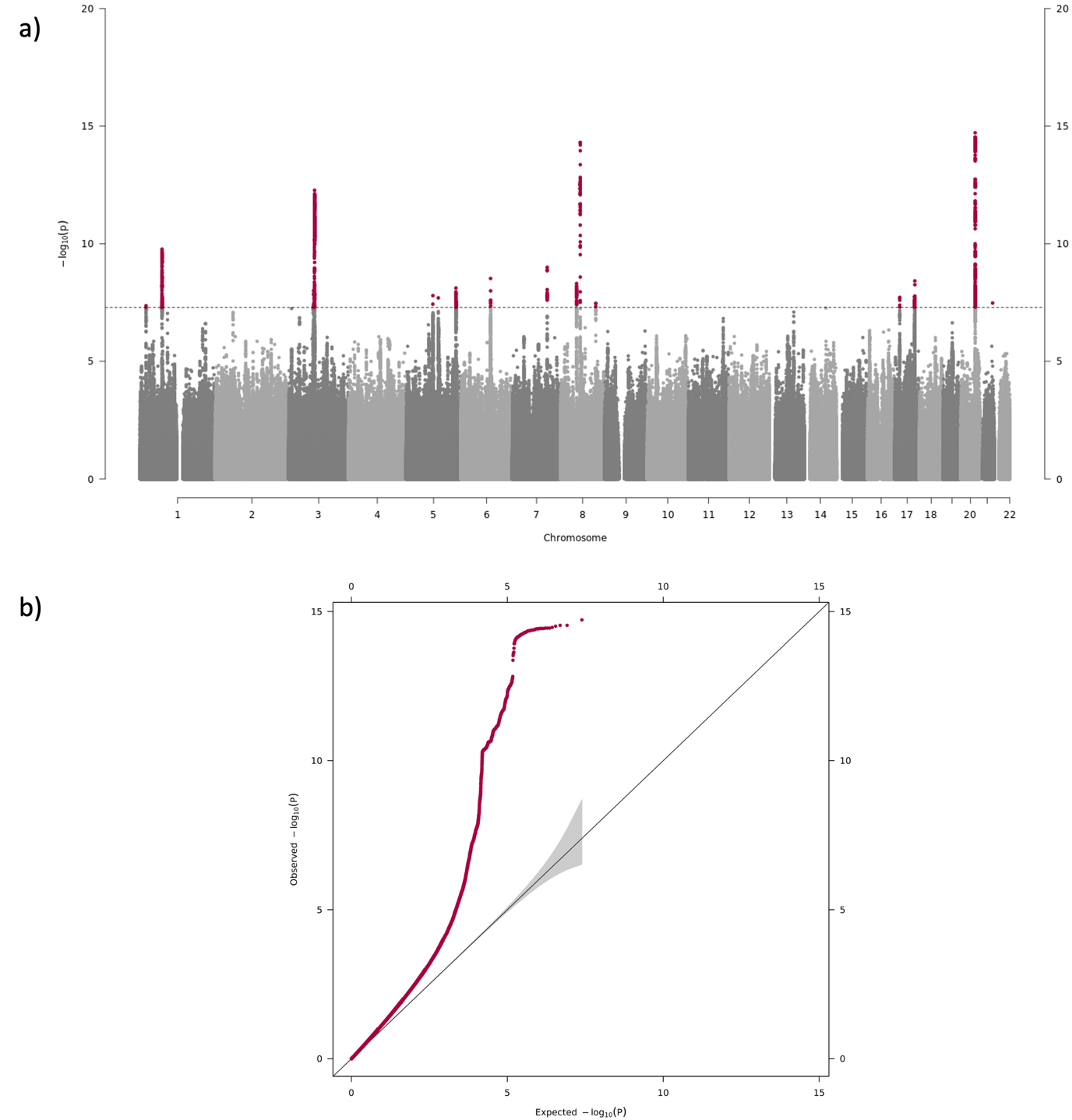

*The Manhattan plot (a) shows the genetic variants per chromosome and their respective p-values from the social isolation genome-wide association study (GWAS). The dotted line represents a p-value threshold of 5x10-08. The genetic variants above this line were clumped for independence. The quantile-quantile (QQ) plot presents the observed-values against the expected p-values for the genetic variants.*
